# The immune response to childhood vaccines is seasonal

**DOI:** 10.64898/2026.04.23.26351620

**Authors:** LA Barrero Guevara, G Féghali, SC Kramer, M Domenech de Cellès

## Abstract

Vaccination programs worldwide have effectively reduced the burden of childhood diseases, yet immune responses remain highly heterogeneous among individuals ^1,2^. While host characteristics such as age and sex are established determinants of vaccine immunogenicity, the timing of vaccination, specifically the calendar season of vaccination, remains largely underexplored ^3^. Although circadian rhythms are known to regulate daily immune function ^4^, evidence for long-term circannual patterns has been limited by the difficulty of collecting year-round vaccination data across diverse populations. Here, we show that the season of vaccination systematically shapes the immune response across a broad range of pediatric vaccines. By leveraging data from 96 randomized control trials worldwide, including over 48,000 children vaccinated against 14 pathogens, we demonstrate that immunogenicity after vaccination follows a pronounced latitudinal gradient, typically peaking during colder months in temperate regions and exhibiting distinct variability in the tropics. These findings suggest that the circadian human immune response might extend to a circannual scale, potentially synchronized by environmental cues. Incorporating the season of vaccination into the design of clinical trials and public health campaigns may optimize vaccine performance and enhance seroprotection.

## Introduction

Global vaccination programs have successfully reduced the burden of childhood infectious diseases ^1^, yet immune responses to vaccination vary widely between individuals ^2^. Understanding the determinants of this heterogeneity is critical for optimizing vaccine schedules and maximizing seroprotection. Age, sex, vaccine formulation, and dose intervals are known to affect immunogenicity ^3^, but the season of vaccination remains underexplored^5^.

While there is growing evidence that the immune response to vaccination fluctuates over a day through circadian rhythms ^4^, much less is known about how these temporal patterns extend to circannual scales. Consistent with circadian rhythms, evidence shows that morning vaccination often elicits stronger immunogenicity than afternoon vaccination—likely reflecting diurnal variation in leukocyte trafficking and effector function ^4^. For circannual rhythms, early studies observed stronger influenza responses when vaccination occurred during winter ^6^, and transcriptomic analyses have shown winter-associated “pro-inflammatory” immune profiles ^7^. Yet whether the calendar season of vaccination systematically shapes immune responses across vaccines and populations remains unclear, in part because obtaining detailed, year-round vaccination data while accounting for host-, vaccine-, and schedule-related determinants has been challenging.

Here, we leverage data from 96 randomized clinical trials (RCTs) involving over 48,000 children vaccinated against 14 pediatric pathogens worldwide to investigate how the calendar season of vaccination shapes immunogenicity. We assess how seasonal patterns shift with latitude and evaluate their consistency across antigens and between binding and functional antibody assays. We find that vaccination season systematically influences the immune response across multiple pediatric vaccines. This seasonal response follows a pronounced latitudinal gradient, consistently reproduced across antigenic targets and assay types. These findings suggest that the established framework of circadian immune regulation might broaden to a circannual scale and demonstrate that the season of vaccination is an important dimension of vaccine response.

## Results

### Participants

To understand how vaccination season and vaccine schedules impact the immune response after vaccination, we analyzed RCTs, whose designs provide rigorous participant selection and documentation of vaccination and immunogenicity measurements. Our data comprised 96 RCTs conducted between 1998 and 2018, including 48,318 participants vaccinated against 14 pathogens after applying inclusion criteria (See Methods and Figure 1). The studies assessed primary vaccinations, and we included only participants with no evidence of prior exposure (seronegative at baseline antibody measurements within 30 days before vaccination). Because immunogenicity after vaccination was measured using different antibody antigens and methodologies across the studies, we restricted our main analyses to the antibody antigen and methodology that included the largest number of participants. Together, these studies provide a uniquely comprehensive dataset for evaluating immune responses to primary vaccination across pathogens that cause common pediatric diseases.

**Figure 1.**
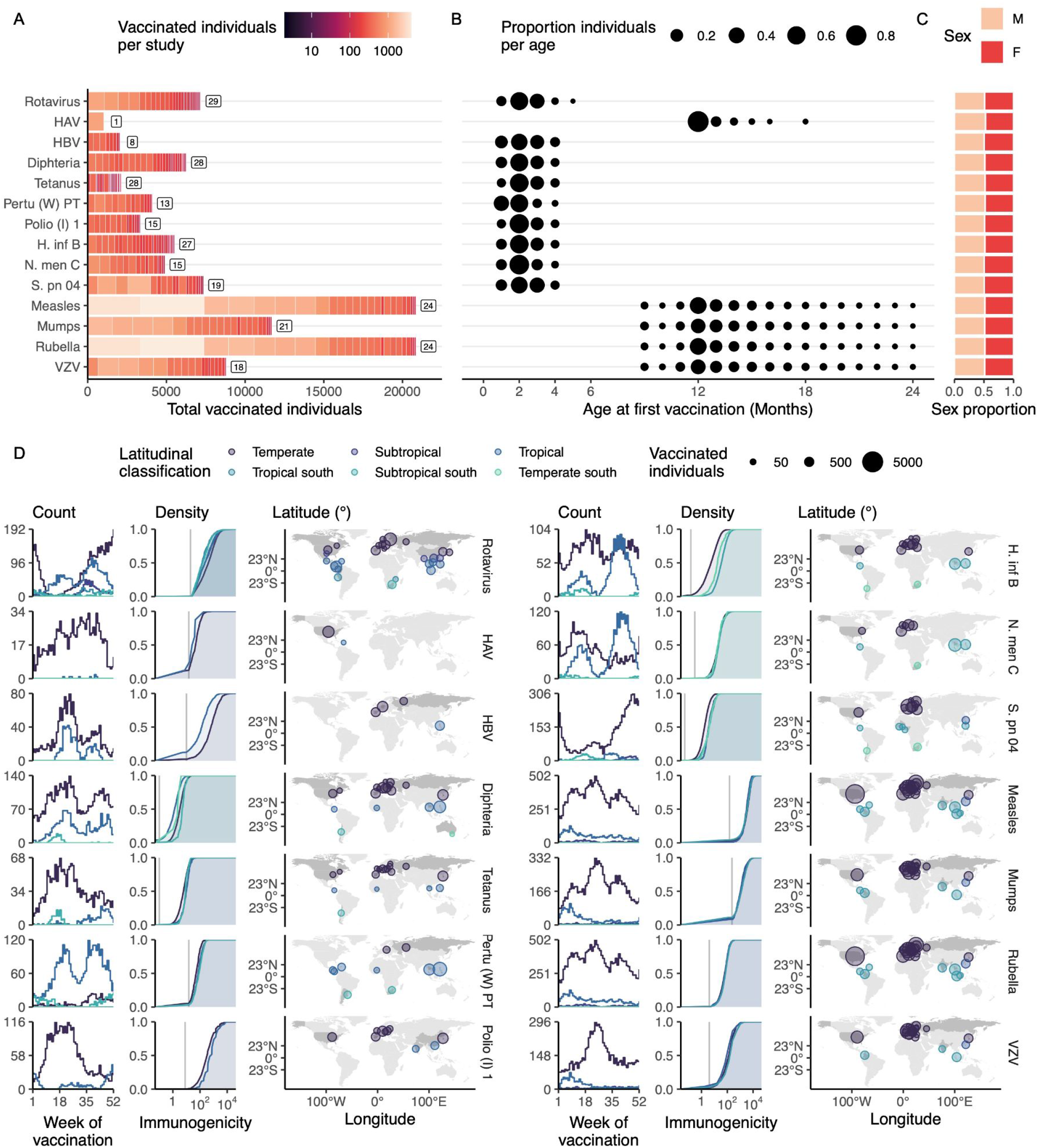
Global distribution, demographic characteristics, and immunogenicity profiles of pediatric vaccination trials included in the study. Between 1998 and 2018, 96 clinical trials of vaccines for 14 pediatric diseases were conducted worldwide. A) Total number of vaccinated individuals per pathogen; color intensity indicates contributions from individual studies, and numbers denote the number of RCTs. B) Age at first vaccination, with point size proportional to the fraction of participants vaccinated at each age (months). C) Sex distribution of participants by pathogen. D) For each pathogen, weekly distribution of vaccinations across the calendar year, density of post-vaccination immunogenicity measurements, and geographic distribution of study sites (point size proportional to the number of vaccinated individuals; colors indicate latitudinal classification). HAV = Hepatitis A virus, HBV = Hepatitis B virus, Pertu (W) = Pertussis (whole-cell vaccine), Polio (I) = Polio (inactivated vaccine), H. inf B = *Haemophilus influenzae B*, N. men = *Neisseria meningitidis*, S. pn = *Streptococcus pneumoniae*.

For each pathogen, we included 1–29 RCTs, comprising 1,022–20,847 participants (Figure 1A). The median age of vaccination was 2 months for rotavirus, HBV, diphtheria, tetanus, pertussis (whole-cell)-PT, polio-1, *H. influenzae B, N. meningitidis*-C, and *S. pneumoniae*-04, and 12 months for HAV, measles, mumps, rubella, and varicella (Figure 1B). As expected for RCTs, the children had an even female-to-male ratio (range 0.47:0.53–0.49:0.51, Figure 1C). Children were vaccinated throughout the year; however, in some cases, due to data limitations, vaccinations were concentrated in a few months (Figure 1D). For most pathogens, immunogenicity after vaccination exceeded the serological threshold (ranging from 88.26% for HAV to 99.97% for polio-1). The studies were conducted in 27 countries, spanning a worldwide range of latitudes from 37.80°S to 63.187°N and longitudes from 89.65°W to 144.96°E. Our data represent a diverse and extensive collection of RCT participants vaccinated throughout the calendar year against pediatric pathogens across different latitudes worldwide.

### The season of vaccination accounts for part of the variability in the immune response to vaccination

We evaluated whether including vaccination seasons improved predictions of post-vaccination immunogenicity by comparing Bayesian multilevel models with and without date of vaccination as a predictor. Baseline models included only established predictors of immunogenicity: sex, age at vaccination, vaccine characteristics, and time between doses. Seasonal models additionally included vaccination date as a sinusoidal term. To allow seasonal patterns to differ across space, we also considered seasonal-by-latitude models, in which the seasonal function varied by latitude. We tested four latitudinal classification schemes, ranging from coarse (two groups) to finer (six groups, Table 1). We found that including the vaccination season improves the models’ fit to the data. The seasonal models had lower widely applicable information criterion (WAIC) than the baseline model for 12 of the 14 pathogens in our main analyses (range of ΔWAIC: 2.36 for HAV and 74.34 for rubella). Despite its greater complexity, the model that included different seasonalities across 6 latitudinal regions was the best for 9 of these pathogens: rotavirus, varicella, measles, rubella, polio, *H. Influenzae B, S. pneumoniae* 04, pertussis (W) PT, and HAV. Interestingly, the difference in WAIC increased as the models incorporated finer information on latitude, allowing seasonality to vary across space (Figure 2A and 2C). For the diphtheria vaccination, the best model was the one incorporating only 2 of the seasonal regions. In contrast, for mumps and meningitidis C, the best model was the one with no change in seasonality across latitudes. Finally, for tetanus and hepatitis B, the base model was the best. Hence, there was no evidence of seasonal variations in the immune response for these two vaccine antigens. Despite the large heterogeneity in immunogenicity measurements, the best-fitting models reproduced the observed data across all pathogens (mean R^2^ = 21.8%, range 6.4%–47.7%; Figures 2B and 2D). Our data show that including vaccination season improves model performance for most pathogens in our study.

**Table 1.** Latitudinal classification schemes.

|  | **Classification** | **Definition** |
| --- | --- | --- |
| **Two groups** | North<br>South | (0°, 90°N]<br>[90°S, 0°] |
| **Three groups** | Temperate north<br>Tropical<br>Temperate south | (23.5°N, 90°N]<br>(23.5°S, 23.5°N]<br>[90°S, 23.5°S] |
| **Four groups** | Temperate north<br>Tropical north<br>Tropical south<br>Temperate south | (23.5°N, 90°N]<br>(0°, 23.5°N]<br>(23.5°S, 0°]<br>[90°S, 23.5°S] |
| **Six groups** | Temperate north<br>Subtropical north<br>Tropical north<br>Tropical south<br>Subtropical south<br>Temperate south | (35°N, 90°N]<br>(23.5°N, 35°N]<br>(0°, 23.5°N]<br>(23.5°S, 0°]<br>(35°S, 23.5°S]<br>[90°S, 35°S] |

**Figure 2.**
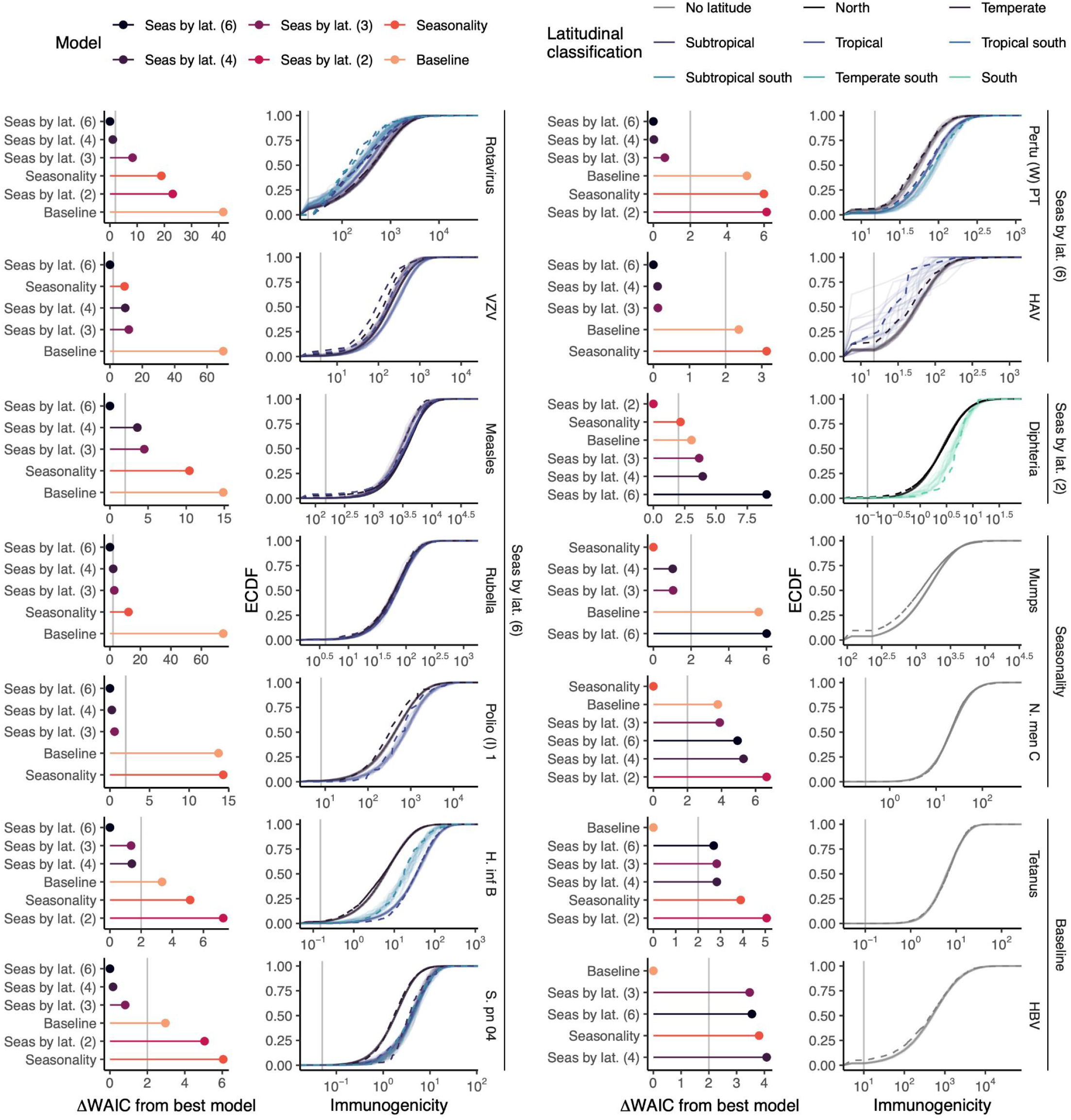
Incorporating vaccination date improves the performance of vaccine immunogenicity models across pathogens and latitudes. Comparison of baseline, seasonal, and seasonal-by-latitude models for each pathogen. Left panels show differences in WAIC relative to the best-fitting model (ΔWAIC), with points indicating model variants (baseline, seasonal only, and seasonal stratified by 2–6 latitudinal groups); lower values indicate better model performance. The vertical grey line denotes a difference of 2 points. Right panels show the corresponding empirical cumulative distribution functions (ECDF) of observed (solid) and model-predicted (dashed) post-vaccination immunogenicity, colored by latitudinal classification. Vertical grey lines denote serological thresholds. Across most pathogens, models including vaccination date—particularly those allowing seasonality to vary by latitude—outperform baseline models and reproduce the observed immunogenicity distributions. HAV = Hepatitis A virus, HBV = Hepatitis B virus, Pertu (W) = Pertussis (whole-cell vaccine), Polio (I) = Polio (inactivated vaccine), H. inf B = *Haemophilus influenzae B*, N. men = *Neisseria meningitidis*, S. pn = *Streptococcus pneumoniae*.

**Figure 3.**
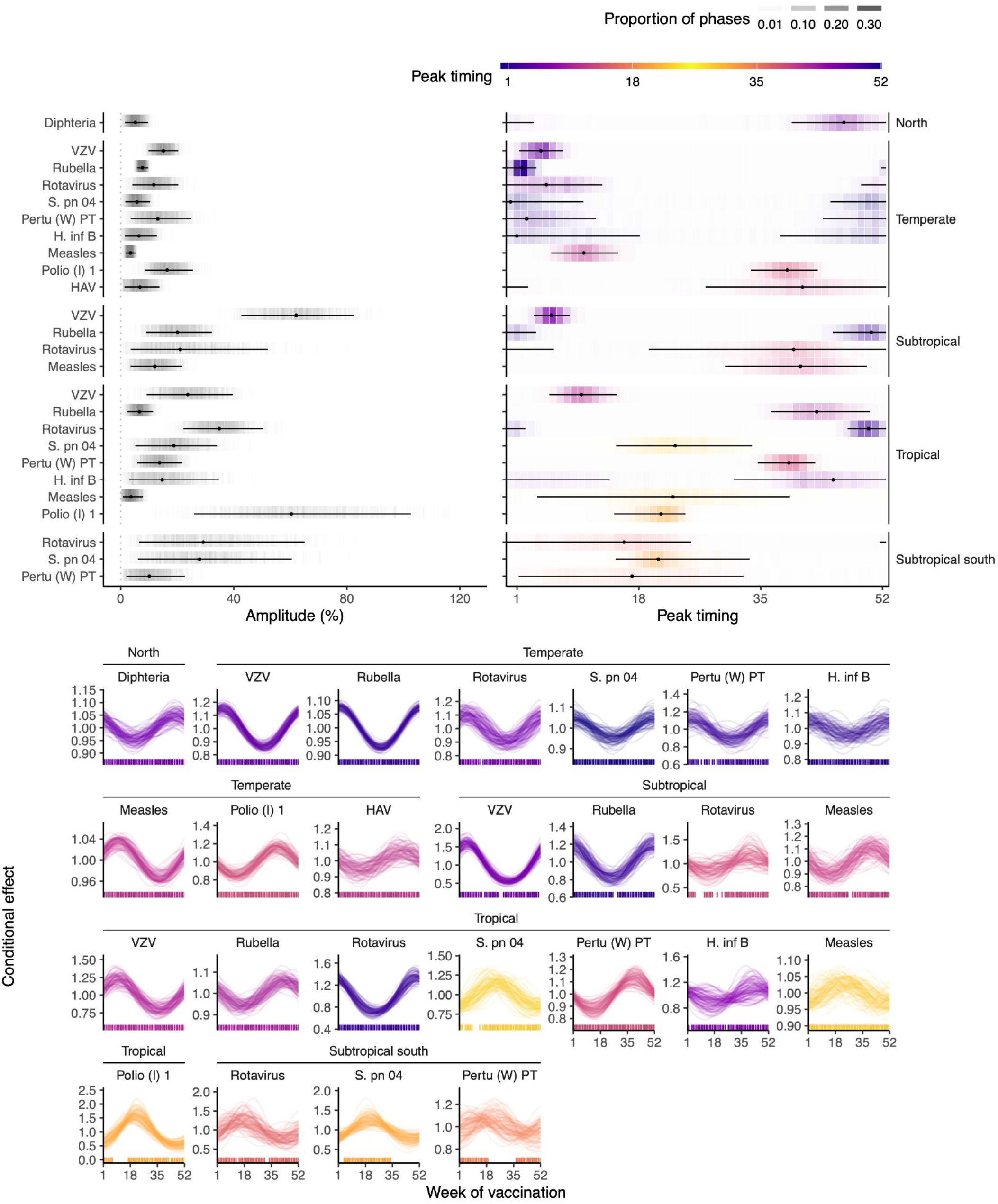
Latitudinal gradients in the seasonal timing and amplitude of the vaccine immunogenicity. Top panels show posterior estimates of seasonal amplitude (left) and peak timing (right, week of year) from the best-fitting seasonal-by-latitude models for each pathogen and latitudinal group (points, medians; lines, 95% credible intervals; color indicates posterior phase density). Bottom panels show posterior conditional effects of vaccination week on immunogenicity for selected pathogens and latitudes, with curves representing posterior draws and rug marks indicating observed vaccination weeks. Peak immunogenicity aligns with the colder months in temperate regions and exhibits distinct timing and greater variability in tropical and subtropical regions. HAV = Hepatitis A virus, HBV = Hepatitis B virus, Pertu (W) = Pertussis (whole-cell vaccine), Polio (I) = Polio (inactivated vaccine), H. inf B = *Haemophilus influenzae B*, N. men = *Neisseria meningitidis*, S. pn = *Streptococcus pneumoniae*.

### The amplitude and peak timing of the seasonal immune response follow a latitudinal gradient

To describe how vaccination season resulted in a seasonal immune response, we extracted the amplitude and phase estimated by the best-fitting models that included seasonality, representing the amplitude and timing of the immunogenicity peak —the time of year when vaccinations elicited a stronger immune response. To assess the robustness of these models in estimating characteristics of the seasonal trend in data with gaps in temporal coverage, we performed posterior predictive checks.

We found that the amplitude of the immunogenicity peak varied across pathogens and had a moderate impact (15.69% change from the mean immunogenicity), ranging from 2.72% for measles vaccination in tropical regions to 61.03% for varicella vaccinations in northern subtropical regions. While the amplitude tended to increase towards the tropical latitudinal regions, the confidence intervals of the estimates were wider due to reduced data availability. For rotavirus and polio 1, we found that peak amplitude was higher in tropical regions than in northern temperate regions (Difference [95% credible interval (CI)] = 23.04% [8.94%, 39.37%] and 45.15% [9.77%, 87.17%], respectively). In addition, while the amplitude for rubella and varicella was not different between the tropical and temperate regions, the amplitude of the peak was higher in subtropical regions than in the northern temperate regions (Difference [95% CI] = 12.44% [1.18%, 24.44%] and 46.54% [26.24%, 68.46%], respectively) and the tropical regions (Difference [95% CI] = 13.34% [0.45%, 25.74%] and 37.66% [12.59%, 63.11%], respectively). Overall, the impact of time of vaccination on immunogenicity increased toward tropical regions.

We also found a latitudinal pattern for the peak timing. In the northern temperate regions, vaccinating very early in the year, in the colder months, elicited the strongest immune response (mean week 1, ranging across the antigens from September to March). Similarly, the northern-hemisphere subtropical regions had a mean peak timing towards the end of the year (mean week 47, ranging from September to January). In contrast, the tropical regions showed greater variability, with peak timings throughout the year (mean week 36, ranging from March to December). Mirroring the results in the northern hemisphere, vaccinating in the middle of the year in the subtropical region of the southern hemisphere, during the corresponding colder months, elicited higher immunogenicity (mean week 18, between May and August). Interestingly, for pathogens whose best-fitting models did not include a latitudinal pattern, the peak timing was estimated at the end of the year, as in northern temperate regions, and was consistent with the majority of their studies being conducted in those regions. Overall, our results reveal a striking latitudinal pattern in the seasonal immune response, indicating that vaccination season can elicit different immunogenicity across latitudes and throughout the calendar year.

To assess the robustness of these models in estimating characteristics of the seasonal trend in data with gaps in temporal coverage, we performed posterior predictive checks. Our results show that the models could reliably recover the seasonal patterns of immunogenicity, despite sparse data, for the majority of latitude groups in each pathogen in the main analyses (Supplementary Figure 3). In addition, we fitted generalized additive models (GAMs) with cyclic splines for date of vaccination to evaluate whether more flexible seasonal structures could reveal patterns not captured by the sinusoidal model. The resulting splines closely matched the cosine fits, consistently exhibiting a single dominant peak for most seasonal immunogenicity patterns (Supplementary Figure 4).

### The seasonal immune responses and their latitudinal gradients remained consistent across antigens

Immunogenicity after vaccination is often assessed using multiple antigens or strain-specific targets, particularly for multivalent vaccines such as those for Poliovirus (types 1–3), *S. pneumoniae* (multiple serotypes), *N. meningitidis* (serogroups A and C), and the acellular pertussis vaccine (PT, FHA, and PRN). We therefore evaluated whether the impact of vaccination season extended to these additional antigenic endpoints beyond those considered in our main analyses. These datasets comprised fewer participants (range: 2,056–9,305) enrolled across 5–28 RCTs conducted throughout the year and across multiple regions worldwide (Supplementary Figure 5).

Consistent with our primary results, the seasonal models performed better than the base model for Poliovirus, *S. pneumoniae*, and *N. meningitidis* (ΔWAIC = 21.5, 26.79, and 9.42, respectively), but not for the acellular pertussis vaccine (Supplementary Figure 6). Notably, the seasonal immune responses for these additional antigens recapitulated the same latitudinal gradients in both peak amplitude and timing described above (Supplementary Figure 7). Because several of these studies were conducted in both northern and southern temperate regions, we observed a mirrored seasonal pattern between hemispheres, with immunogenicity peaking during the colder months of each hemisphere (mean peak timing: week 48 in the north and week 18 in the south).

Within each pathogen, estimates of peak amplitude and timing were highly consistent across antigens (Supplementary Figure 7). The only notable exception was *S. pneumoniae* 6B, which showed a stronger seasonal response in tropical regions than other *S. pneumoniae* serotypes. Additionally, while the whole-cell pertussis vaccine showed improved performance when the vaccination season was included, this was not the case for the acellular vaccine formulation, which may reflect the different immune responses elicited by the two vaccines. Together, these analyses demonstrate that the seasonal effects and their latitudinal pattern are robust across antigenic targets.

### The seasonal immune responses and their latitudinal gradients remained consistent when considering functional antibody measurements

Immunogenicity after vaccination can also be assessed using functional assays, which quantify not only the presence of antibody response (e.g., ELISA or IFA), but also its biological activity, such as bactericidal, opsonophagocytic, or neutralizing capacity (e.g., BAC, OPA, PRNT). We evaluated whether vaccination season influenced these functional immune responses. In our main analyses, all immunogenicity endpoints were measured by binding assays, except for polio (PRNT). Because most RCTs did not include such measurements, these analyses relied on a smaller subset of studies for diphtheria, mumps, *N. meningitidis*, and *S. pneumoniae*, comprising 197–5,051 participants across 4–5 RCTs (Supplementary Figure 8).

We found that the seasonal models outperformed the baseline models for *N. meningitidis* and *S. pneumoniae* (ΔWAIC = 8.15 and 3.22, respectively, and Supplementary Figure 9), but not for diphtheria or mumps. For vaccinations against these latter pathogens, the limited geographic and latitudinal coverage likely hindered the models’ ability to detect seasonal patterns (Supplementary Figure 8). For *N. meningitidis* and *S. pneumoniae*, however, the seasonal immune responses mirrored the previously observed latitudinal gradients in peak amplitude and timing, although with greater uncertainty due to the reduced sample sizes (Supplementary Figure 10). Overall, these results, together with those from polio described above, show that seasonal patterns in immunogenicity extend to functional antibody measurements. Nevertheless, the magnitude of seasonal differences might be less pronounced at the functional level than in general antibody responses.

### Season of birth did not improve the model fit to the immune response to vaccination

Beyond the season of vaccination, the season of birth could also influence immunogenicity. Although our models already adjust for age—a known determinant of vaccine response—birth timing could still have an independent effect. For example, perinatal seasonal exposures may shape early immune function, with consequences that might persist or accumulate into the first months of life and modulate responses to subsequent vaccination ^8,9^. To evaluate this possibility and ensure that the influence of season of vaccination was not concealing that of season of birth, we compared seasonal models with season of birth to those with season of vaccination. Despite the very strong correlation between these two dates, the season of birth did not improve model fit and was often outperformed by the vaccination season (Supplementary Figure 13). These results indicate that the seasonal patterns in immunogenicity observed in our study are primarily driven by the vaccination season rather than the season of birth.

### Characteristics of vaccinated individuals, vaccine schedules, and the vaccine composition influence immunogenicity

Beyond the season of vaccination, we found that intrinsic characteristics of vaccinated individuals, vaccine schedules, and vaccine properties strongly shaped the immune response, consistent with previous studies (Figure 4 and Supplementary Results). Across vaccines, males showed weaker immune responses than females, and older age resulted in stronger responses. Longer intervals between successive doses enhanced antibody responses for diphtheria, polio, and HBV, whereas for *N. meningitidis*, longer intervals slightly reduced the response. The time between the final dose and the immunogenicity measurement also influenced the observed immune response, reflecting the initial rise and subsequent decline of antibody levels after vaccination. Differences in vaccine type and concentration further contributed to variation in immune responses. We also found that immune responses were correlated when vaccines were administered as multivalent formulations. Finally, we characterized the underlying dispersion of antibody responses across individuals by estimating the shape parameter (α) of the Gamma distribution. Altogether, these findings demonstrate that the highly heterogeneous immune response after vaccination reflects a combination of individual, schedule-dependent, and vaccine-specific characteristics.

**Figure 4.**
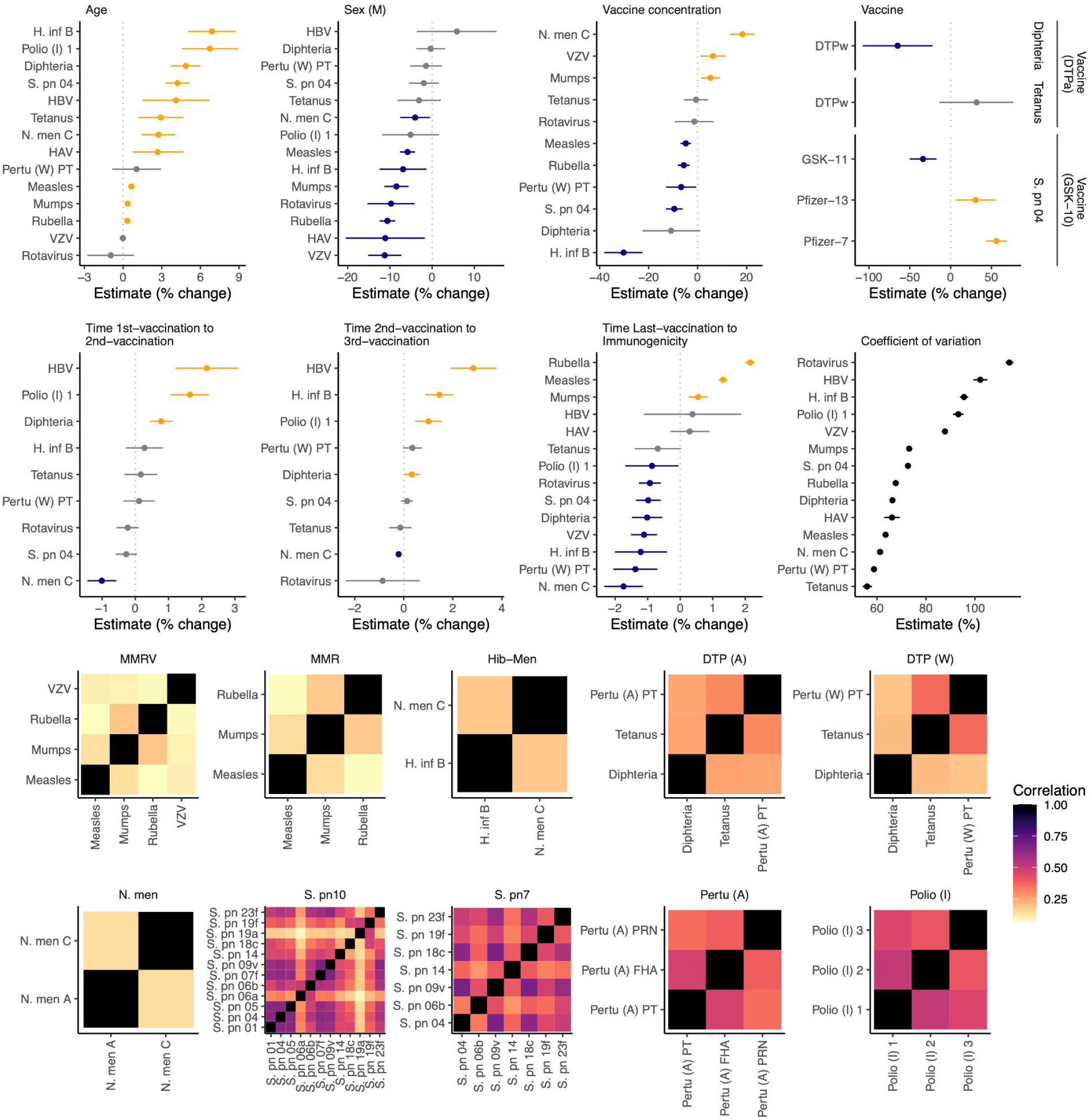
Host, schedule, and vaccine determinants of post-vaccination immunogenicity. The top panels show posterior estimates (points, medians; lines, 95% credible intervals) of fixed effects from the best-fitting models across pathogens: age at vaccination, sex (male vs female), vaccine concentration, vaccine formulation, intervals between doses, time from last vaccination to immunogenicity measurement, and the coefficient of variation of antibody responses (estimated from the shape parameter). Heatmaps show correlations between immune responses to multiple antigens within multivalent vaccines (e.g., MMR/MMRV, Hib–Men, DTP, pneumococcal serotypes, polio types, and acellular pertussis components), with darker colors indicating stronger correlations. HAV = Hepatitis A virus, HBV = Hepatitis B virus, Pertu (W) = Pertussis (whole-cell vaccine), Polio (I) = Polio (inactivated vaccine), H. inf B = *Haemophilus influenzae B*, N. men = *Neisseria meningitidis*, S. pn = *Streptococcus pneumoniae*.

## Discussion

Our study provides the most comprehensive evidence to date that the vaccination season meaningfully shapes immunogenicity across a broad range of pediatric vaccines, establishing seasonal variation as a fundamental property of the human immune response. By integrating data from over 48,000 children across 96 RCTs conducted worldwide, we show that including the vaccination season consistently improves model performance for 12 of the 14 pathogens analyzed. This seasonal pattern follows a striking latitudinal gradient: immunogenicity consistently peaks in colder months in temperate regions, while exhibiting distinct, higher-amplitude variability in tropical regions. Importantly, we further identified these spatiotemporal patterns across multiple antigenic endpoints and in functional antibody assays, underscoring the robustness of our findings. Beyond seasonality, we also identified substantial contributions of individual characteristics, vaccine schedules, and vaccine formulations to immunogenicity, consistent with previous evidence. However, our data reveal that these known determinants operate within a seasonally dynamic baseline. Altogether, our findings show that the heterogeneous immune response to vaccination arises from an interplay among host-, schedule-, and vaccine-related determinants, which is also shaped by the season of vaccination.

We found that the vaccination calendar season explained part of the variability in immune responses across vaccines. Prior work examining timing effects has largely focused on circadian rhythms ^4^. A recent systematic review showed that morning vaccination generally elicited stronger antibody responses than afternoon vaccination across several vaccines ^10^, including influenza, SARS-CoV-2, *S. pneumoniae*, HBV, and HAV. Our results suggest that this chronobiological framework can be extended to a circannual scale. Few studies have sought evidence of season-of-year effects on immunogenicity ^5^. When assessed, the immunogenicity peaks seem to occur in winter, as reported for rubella and polio in Israel ^11,12^ and for influenza in the USSR ^6^. In line with our results, more variable patterns have been documented in tropical settings: in The Gambia, *S. pneumoniae* responses peaked during the rainy season, whereas oral polio vaccine responses were higher in the dry season ^13^. We notably did not detect seasonal effects for tetanus or HBV, consistent with some prior studies ^13–16^. For HBV in particular, the strong influence of booster intervals may obscure seasonal variation in immunogenicity following primary vaccination—an interpretation supported by our models, which showed that booster intervals modulated the HBV response more than that of other pathogens. Notably, one study conducted in Austria found that the season of the final HBV dose affected the vaccination responses ^16^. Taken together, our results provide robust evidence that vaccination season influences immunogenicity across multiple pathogens.

The seasonal response to vaccine immunogenicity we observed could be explained by two distinct mechanisms: external ecological dynamics (e.g., the seasonality of pathogen circulation ^17,18^) or internal physiological rhythms (e.g., host chronobiology ^4^). A primary consideration is whether these peaks simply reflect specific immune activation or heterologous immunity driven by concurrent seasonal infections. However, the rigorous design of the included RCTs minimizes this possibility. We strictly included participants who were seronegative at baseline and excluded those with symptomatic infections or important comorbidities (e.g., HIV ^19^) that could impair or artificially boost immune response. While we cannot strictly rule out that asymptomatic, non-target infections might transiently modulate the immune baseline—like seasonal malaria has been proposed to influence vaccine responses in The Gambia ^13^—the systemic consistency of our findings across 14 distinct pathogens argues against such external drivers. Furthermore, the short interval between vaccination and outcome measurement (typically 1-2 months) limits the window for unobserved infections to significantly affect our results, particularly for pathogens with low circulation rates, such as measles ^5^. Thus, the observed seasonality likely reflects intrinsic modulation of the host’s immune status rather than the dynamics of seasonal pathogens.

Our results add to the growing body of evidence that the immune system possesses a “circannual oscillator” analogous to the circadian clock. Just as daily rhythms regulate leukocyte trafficking and effector functions—mechanisms recently shown to optimize responses to vaccines administered in the morning versus the afternoon ^4,20,21^—our data suggest these mechanisms scale to a seasonal rhythm. This hypothesis is supported by transcriptomic studies demonstrating that ∼23% of the human genome exhibits seasonal variability in expression ^7^. In temperate regions, this manifests as a wintertime “pro-inflammatory profile”, characterized by upregulated IL-6, IL-1β, B-cell receptor signaling ^7^, and elevated leukocyte counts ^22^. In tropical regions, the changes in the circulating immune cell composition instead peak during the rainy season ^13,23^. This “pro-inflammatory” profile might lower the activation threshold for immune response, priming a more robust response to vaccination during these periods.

The striking latitudinal gradient we identified suggests that local environmental cues, or *Zeitgebers*, may synchronize seasonal variation in vaccine responses. The improved model fit with finer latitudinal stratification indicates that this rhythm does not follow a universal calendar but instead aligns with local environmental conditions. Photoperiod has been proposed as a major regulator of the immune circadian clock, potentially acting through endocrine pathways such as melatonin ^24^. While this mechanism could explain the winter peaks observed in temperate regions, photoperiod alone cannot account for the high-amplitude and distinct peaks we observed in the tropics, where day length is relatively constant. Our findings, therefore, point to additional environmental variables. Experimental work has shown that ambient temperature can modulate adaptive immunity, including impaired response to influenza infection in mice ^25^, and transcriptomic studies have revealed marked differences between tropical and temperate populations that cannot be attributed to photoperiod alone ^7^. Other proposed cues, such as sleep and dietary patterns, are unlikely to vary in a manner that explains the broad geographic patterns we observed. Together, our results suggest that the seasonal immune response to vaccination may be cued not only by photoperiod but also by local climatic or environmental conditions.

Crucially, the spatiotemporal patterns of the seasonal immune response were reproducible across distinct immunogenicity measures, reinforcing the clinical relevance of our findings. For vaccines where binding antibody concentrations are established as correlates of protection—such as HAV, varicella, and *H. influenzae B* ^26–29^—we confirmed a clear seasonal response. Similarly, for *S. pneumoniae* and *N. meningitidis*, where protection is best defined by functional activity ^26,30,31^, we found that functional antibody responses followed the same latitudinal gradients. While the lack of defined correlates of protection for some pathogens, such as pertussis ^32^, limits our assessment of immune protection, the consistency we observed between binding and functional assays suggests that seasonal modulation affects not only the strength of the immune response but also its quality and effector function. Although the seasonal signal/strength was slightly attenuated in functional assays, its persistence implies that vaccination season could have tangible consequences for clinical protection.

Beyond the season of vaccination, our analyses corroborate established evidence that host, vaccine schedules, and vaccine characteristics can shape immune response to vaccination. In line with prior comprehensive reviews ^33,34^, we found that the immune response in girls was consistently higher or non-inferior to that of boys. These differences could reflect hormonal, environmental, and genetic mechanisms, including X-chromosome gene dosage effects. We also identified a distinct age-dependent pattern, with older infants consistently exhibiting a stronger immune response, a trend that was most pronounced during early infancy (2-4 months) compared to the second year of life ^35,36^. As our inclusion criteria excluded participants with baseline seropositivity—effectively ruling out interference from maternal antibodies—this age trend almost certainly reflects the functional maturation of the infant immune system ^36,37^. Furthermore, dosing schedules also impacted the immune response. Extended intervals between doses generally enhanced immunogenicity, as observed for HBV, polio, and diphtheria ^38–40^. Conversely, for *N. meningitidis* vaccination, shorter intervals appeared to slightly boost responses ^41^. Finally, we found that vaccine characteristics can also influence the immune response to vaccination, including vaccine concentration and multivalent vaccines.

Our study has limitations inherent to the use of aggregated RCT data. First, while we account for major vaccine types, we did not explicitly control for more detailed differences in vaccine formulations, such as specific adjuvant systems, manufacturing lots, or strain compositions ^42^. However, our inclusion of random effects within RCT arms effectively captures the clustering of individuals under these specific conditions. Second, we did not evaluate the immunomodulatory impact of concomitant vaccinations ^43^. Yet, given the diversity of schedules across our dataset, it is unlikely that co-administration patterns would introduce a systematic seasonal bias sufficient to drive the consistent latitudinal gradients we observed. Finally, we lacked data on the time of day of vaccination. Although vaccination timing during the day was unlikely to shift consistently throughout the year, future clinical trials should record the exact timing of vaccination, given the growing evidence of its impact on immune response ^5^. A critical next step will be to disentangle the distinct yet potentially interacting roles of circadian and circannual rhythms, and to elucidate the environmental cues and biological mechanisms that govern immune response across these temporal scales.

Our findings have critical implications for both clinical trial design and public health policy. Currently, vaccine efficacy trials are randomized by individual characteristics but rarely consider the vaccination season ^5^. Because recruitment often occurs within defined temporal windows, unadjusted comparisons between trials conducted at different times of the year may confound seasonal variation with vaccine performance. We therefore propose that the vaccination season be recorded and modeled as a standard covariate in future RCTs. From a public health perspective, our findings could help optimize the timing of mass catch-up vaccination campaigns ^44^. For example, scheduling campaigns for measles or polio during periods of peak immunogenicity could maximize seroconversion rates, particularly in resource-limited or hard-to-reach settings.

In summary, we present the first worldwide, multi-pathogen atlas of the seasonal immune response to pediatric vaccination. By integrating data from RCTs with over 48,000 children, we demonstrate that vaccine response is not constant throughout the year but is governed by distinct seasonal rhythms that vary across latitudes. Crucially, this seasonality proved robust across diverse antigenic targets and functional assays. Our findings extend the framework of immune response from a circadian clock to a circannual calendar, suggesting that accounting for vaccination season in vaccine schedule design could enhance our ability to induce optimal immune responses.

## Methods

### Data

#### Clinical trials data

We requested data from 265 randomized clinical trials (RCTs) of vaccines for childhood infections (Supplementary methods, Supplementary Figure 1, and Supplementary Table 1) via the ClinicalStudyDataRequest.com (CSDR) platform. The studies included vaccinations against pathogens causing common pediatric bacterial diseases, namely Corynebacterium diphtheriae (diphtheria), Clostridium tetani (tetanus), Bordetella pertussis (pertussis), Haemophilus influenzae B (HiB, haemophilus influenzae diseases), Neisseria meningitidis (meningococcal disease), and Streptococcus pnuemoniae (pneumococcal disease), as well viral diseases, namely Hepatitis A virus (HAV, hepatitis A), Hepatitis B virus (HBV, hepatitis B), Poliovirus (polio), Rotavirus (rotavirus infection), Measles morbillivirus (measles), Mumps virus (mumps), Rubella virus (rubella), and Varicella-Zoster virus (VZV, varicella). The data included information on each participant (age at recruitment, sex, birth date, anonymized center, country, RCT group, and eligibility), vaccination (name, vaccination date, administration), and immunogenicity metrics (target, method, sample date, cut-offs, quantitative results; Supplementary Table 2). We validated these data against the RCT’s protocols and further extracted information on specific inclusion and exclusion criteria, as well as vaccine concentrations.

#### Inclusion and exclusion of studies and participants

Studies were excluded if they did not meet population criteria (14), had missing data (64), or were already included within another study (66). The studies were assigned to at least one of 14 pathogen groups. For inclusion in the specific pathogen groups, studies needed to assess primary vaccination and provide immunogenicity data against the specific pathogen. Then, individual participants were excluded based on data completeness (age, sex, vaccination date, vaccination concentration, immunogenicity results, immunogenicity method, and baseline immunogenicity results) and additional criteria (eligibility, >30 days between immunogenicity baseline and primary vaccination, age >26 months for HAV, measles, mumps, rubella and VZV and >6 months for all other pathogens, <6 months between last vaccination and immunogenicity measurement, and immunogenicity values at baseline over the cutoff). We then divided the data into per-antigen measurements for the subsequent analyses (Supplementary Table 3).

#### Spatial data

Most studies reported only participants’ country of residence. Because countries can cover large areas, to better define participants’ locations, we extracted center locations from the GSK database (https://www.gsk.com/en-gb/) and assigned each to a latitudinal group. As the center locations could not be traced back to each participant due to anonymization procedures, participants were included only if more than 80% of the locations within their country fell within the same latitudinal group. When the study did not provide any center locations, we defined them based on the capital city of the first-level administrative division.

### Model of immunogenicity response to vaccination

We modeled post-vaccination immunogenicity, quantified as antibody titers, using Bayesian generalized multilevel (mixed-effect) models for each antigen. In the *baseline models*, we did not account for seasonal variation in immunogenicity but included sex, age at vaccination, vaccine concentration, interval between vaccine doses, and time from the last vaccine dose to serology sampling as fixed effects. To account for the hierarchical data structure, we specified random effects/intercepts at both the highest level (study) and lowest level (study-country-center-RCT group).

To assess the seasonality in immunogenicity by vaccination season (defined as the calendar week of vaccination), we formulated *seasonal models* by adding a sinusoidal function (i.e., sine and cosine of date of vaccination scaled to an annual cycle):

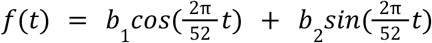

where amplitude (*A*) was estimated on the natural scale as the proportional difference between the peak and the mean of the fitted annual curve, and peak timing (*φ*) as the timing of the peak:

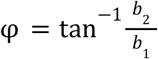

We further extended this to *seasonal-by-latitude models*, allowing the sinusoidal term to vary across latitudinal groups. We tested four latitudinal classification schemes (Table 1). Because available data are spatially limited and clustered (e.g., towards the Northern Hemisphere; Figure 1), considering coarse (two groups) to finer (six groups) classifications allowed us to explore how latitude modifies seasonality without overinterpreting beyond what the spatial coverage supported.

For all models, we assumed the antibody titers follow a Gamma distribution with a log link. The serological assays have a lower detection threshold (or sensitivity). We treated such values as censored and constrained within the [0, threshold] interval. We used the default, non-informative or weakly informative priors from the brms package in R: for fixed effects, an improper flat prior over the real line; for random-effect standard deviations, a half Student-t prior with 3 degrees of freedom (restricted to non-negative values), and for the shape parameter, a Gamma(0.01, 0.01) prior. The models were fitted using four Markov chains, each with 2,000 warm-up iterations followed by 10,000 sampling iterations. We used initial parameter values set to 0.

### Model comparison

To evaluate whether accounting for seasonal variation in immunogenicity by vaccination season improved model fit, we compared the baseline models from the seasonal and seasonal-by-latitude models. For each model, we computed the widely applicable information criterion (WAIC) from the posterior log-likelihood, which is comparable to the frequentist Akaike Information Criterion (AIC). We then identified the best model as the one with the lowest WAIC, defining a minimum difference of 2 points as meaningful.

### Model power assessment

Although most pathogen-antigen subsets combined data from RCTs in which individuals were vaccinated throughout the calendar year, in some subsets the vaccinations occurred only during a few weeks (Supplementary Figure 2). To evaluate whether gaps in temporal coverage could compromise our models’ ability to recover seasonal patterns of immunogenicity, we performed posterior predictive checks. For each pathogen-antigen subset, we simulated 100 datasets from its best-fitted model, preserving the original/observed weekly counts of vaccinated individuals. In each simulation, all covariates except the cosine terms were set to 0. We then refitted the models using only the cosine terms on those simulated datasets, extracted amplitude and phase estimates, and compared them with the estimates from the original fit to the observed data. Models whose simulation-based bias exceeded 20% for amplitude and 7 weeks for the phase were excluded from further interpretation (Supplementary Figure 3).

### Sensitivity analyses

To evaluate the robustness of our findings, we performed several complementary sensitivity analyses. First, to assess sensitivity to the assumed seasonal functional form, we also fitted generalized additive models (GAMs) with cyclic cubic splines for the date of vaccination. These models allowed for more flexible seasonal shapes without imposing a sinusoidal structure. We compared the resulting seasonal patterns to those obtained from the cosine models and found that GAM-based fits yielded highly similar shapes, typically characterized by a single dominant seasonal peak (Supplementary Figure 4). Second, we examined whether the estimates depended on the assay used to measure immunogenicity, repeating the analyses across different measurement techniques. Third, given the bimodal distribution of rotavirus immunogenicity, as some children did not develop an immune response after vaccination, we compared estimates from models that included all immunogenicity results with those from models restricted to immunopositive individuals. Fourth, for vaccines that generate responses to multiple antigens—for example, polio (anti-polio 1, 2, and 3) or MMR (anti-measles, anti-mumps, and anti-rubella)—we used multivariate models to assess whether jointly modeling the correlated antigen responses altered the estimated seasonal effects. Finally, we tested whether the season of birth, rather than the season of vaccination, better explained seasonal variation in immunogenicity by refitting the seasonal and seasonal-by-latitude models with birth season as the time variable and comparing model fit using WAIC.

## Data Availability

The data underlying this study cannot be shared publicly by the authors, but may be requested through the independent data-sharing platform ClinicalStudyDataRequest.com. All data included in these analyses were provided by GlaxoSmithKline through this platform. GlaxoSmithKline did not participate in the data analyses or in the writing of the manuscript.

## Acknowledgements

We thank both CSDR and GlaxoSmithKline Research & Development Ltd for providing access to the clinical trial data via the www.clinicalstudydatarequest.com website. We thank the Infectious Disease Epidemiology laboratory for a group discussion of the manuscript during a group meeting. This study was funded by the Max Planck Society.

This publication is based on research using data from GlaxoSmithKline, which has been made available to us through secured access. The CSDR team or GSK team has not contributed to or approved, and is not in any way responsible for, the contents of this publication.

## Code availability

The code used to perform the analyses and generate the figures in this study has been deposited in a GitHub repository (https://github.com/laurabarreroguevara/code_exp_imm.git) and will be made publicly available upon publication.

