## Supplemental materials for "The immune response to childhood vaccines is seasonal"

#### Supplementary methods

##### Data selection

We obtained data from 265 randomized clinical trials (RCTs) of vaccines for childhood infections via the ClinicalStudyDataRequest.com (CSDR) platform (Supplementary Table 1). We excluded 144 initially because they did not meet the study population criteria, had missing data, or were duplicates (14, 64, and 66 studies, respectively; Supplementary Figure 1). We then classified each RCT according to at least one of 14 pathogen groups, provided the vaccines against the specific pathogen were applied to children for the first time, and the immunogenicity was subsequently measured. In addition, four studies were excluded because they lacked pathogen-specific baseline antibody measurements, which we required to assess or confirm prior exposure to the pathogen. The remaining 117 studies, including 85,165 participants, were evaluated for inclusion at the participant level. Of these, only 110 studies were retained, as they contained complete records for age, sex, vaccine applied, vaccination date, vaccine concentration, immunogenicity measurement, immunogenicity date, immunogenicity result, and immunogenicity method. Furthermore, baseline antibody measurements had to be obtained within 30 days of the primary vaccination, yielding baseline seronegative immunogenicity results. Participants also needed to be at least 1 month old at the time of first vaccination, with age restrictions set at under 26 months for measles, mumps, rubella, and HAV vaccinations, or under 6 months for all other pathogen vaccinations. Additionally, the time between vaccination and serology testing had to be less than 6 months. Due to anonymization, most studies reported only participants' country of residence. Because countries can cover large areas, to better define participants' locations, we extracted center locations from the GSK database (https://www.gsk.com/en-gb/) and assigned each to a latitudinal group. Because center locations could not be traced back to each participant due to anonymization procedures, participants were included only if more than 80% of their locations within their country fell into the same latitudinal group. When the study did not provide any center locations, we defined them based on the capital cities of the first-level administrative divisions. Finally, our data comprised 96 studies, involving 48,318 participants.

#### Supplementary results

##### Characteristics of vaccinated individuals, vaccine schedules, and the vaccines influence immunogenicity

Intrinsic characteristics of the vaccinated individuals, vaccine schedules, and vaccines strongly shaped the immune response (Figure 4). Across vaccines, and consistent with previous studies[1,2], males showed weaker immune responses than females. This sex difference was consistently detected for all vaccines administered at 1 year of age—measles, mumps, rubella, varicella, and HAV—and for most of the earlier-infancy vaccines, including rotavirus, *H. influenzae B*, and tetanus (Range of estimates [95% credible interval (CI)] = –11.24% [–15.18%, –7.29%], –4.02% [–7.56%, –0.50%]). In addition, as found in other studies[3,4], age at vaccination also had a clear positive effect on immunogenicity, with older infants displaying stronger responses. Interestingly, this effect appeared milder for vaccines given at 1 year of age (Range of estimates [95% CI] = 0.34% [0.19%, 0.48%], 2.68% [0.75%, 4.69%]) compared with those administered in early infancy (Range of estimates [95% CI] = 2.75% [1.45%, 4.05%], 6.90% [5.04%, 8.76%]), a pattern consistent with rapid developmental maturation of the infant immune system.

Other characteristics of the vaccine schedules also affected the immune response after vaccination (Figure 4). As described in other studies[5–7], we found that longer intervals between successive doses enhanced the antibody response for diphtheria, polio, and HBV, both between the first-to-second (Estimates [95% CI] = 0.78% [0.44%, 1.12%], 1.65% [1.08%, 2.22%], and 2.15% [1.21%, 3.10%], respectively) and with a milder effect the second-to-third doses (Estimates [95% CI] = 0.33% [0.01%, 0.65%], 1.00% [0.46%, 1.54%], and 1.45% [0.89%, 2.00%], respectively). In contrast, for *N. meningitidis*, longer intervals between doses resulted in a slightly weaker immune response (Estimates [95% CI] = –1.01% [–1.44%, –0.57%], –0.21% [–0.28%, –0.13%] for the first-to-second and second-to-third intervals, respectively).

The interval between the last vaccine dose and the immunogenicity measurement also affected the measured immune response (Figure 4). Of note, these intervals capture short time periods (1–2 months after vaccination), as the RCTs intended to measure immunogenicity consistently across individuals. For vaccines for which immunogenicity was measured after a single dose—measles, mumps, and rubella—the immune response increased over time shortly after vaccination (Estimates [95% CI] = 1.31% [1.17%, 1.45%], 0.55% [0.26%, 0.84%], and 2.15% [2.00%, 2.30%], respectively), consistent with the progressive establishment of adaptive immunity after first exposure. For vaccines for which immunogenicity was measured after primary set (two or three doses), however, longer intervals showed a slight decrease in immune response (Range of estimates [95% CI] = –1.74% [–2.33%, –1.15%], –0.87% [–1.69%, –0.06%]), reflecting the much more rapid rise and subsequent decay of the immune response after secondary exposure. The varicella vaccination showed a different pattern; despite being a primary vaccine, its immunogenicity appeared to decline as early as 6 weeks post-vaccination (Estimate [95% CI] = –1.11% [–1.51%, –0.71%]). This may reflect the different immune pathways elicited by the varicella vaccine.

Differences in vaccine formulation also contributed to variation in immune response (Figure 4). For DTP vaccines, diphtheria responses were weaker with the whole-cell vaccine than with the acellular vaccine (Range of estimates [95% CI] = –64.87% [–107.73%, –22.01%]), while tetanus responses were similar across the two vaccines. For *S. pneumoniae* serotype 4, the GSK-10 formulation produced a weaker response than the Pfizer formulations (Estimates [95% CI] = 56.06% [43.40%, 68.81%] and 30.69% [6.20%, 55.08%], for Pfizer-7 and -13, respectively), and among GSK vaccines, the GSK-10 formulation elicited a stronger immune response than GSK-11 (Estimate [95% CI] = –-33.73% [–50.02%, –17.29%]), consistent with its higher antigen concentration (Concentration = [6 μg/mL] and [2 μg/mL], respectively). Similarly, we found that the higher the vaccine concentration, the stronger the immune response against *N. meningitidis* C, varicella, and mumps (Estimates [95% CI] = 18.31% [13.19%, 23.47%], 5.29% [1.40%, 9.15%], 6.27% [1.05%, 11.55%]). In contrast, we found that the vaccine concentration was negatively associated with the immunogenicity against measles, rubella, pertussis PT, *S. pneumoniae* serotype 4, and *H. influenzae* B (Range of estimates [95% CI] = –30.22% [–38.12%, –22.52%], –4.83% [–7.02%, –2.65%]). Although these heterogeneous results in vaccine concentrations have been reported before[8], this variable might be a proxy for other vaccine differences, such as adjuvant systems, vaccine lots, and formulations, which we did not explicitly account for.

Of note, we also characterized the underlying dispersion of antibody responses across individuals by estimating the shape parameter ($\alpha$) of the Gamma distribution. This parameter is directly related to the coefficient of variation ($CV = 1/\sqrt{\alpha}$), providing a measure of relative variability. Across the different antigens, our estimates for $\alpha$ ranged from 1 to 3.5, which corresponds to a CV from 55.95% [54.14%, 57.97%] to 112.48% [115.73%, 114.04%] (Figure 4). This high degree of dispersion underscores the substantial inter-individual variability inherent in immunogenicity measurements, even within the controlled scheme of randomized clinical trials.

Finally, because several vaccines target multiple pathogens, serotypes, serogroups, or antigens, we examined correlations in immune responses across these components using multivariate models (Figure 4). We found that immune responses were correlated when vaccines targeted antigenically related components of the same pathogen (Range of correlations [95% CI] = 0.092 [0.055, 0.129], 0.676 [0.639, 0.709]). Correlations were less strong when vaccines targeted phylogenetically distant pathogens—for example, in multi-pathogen formulations such as MMRV, MMR, Hib–Men, DTPa, and DTPw (Range of correlations [95% CI] = 0.076 [0.052, 0.100], 0.367 [0.238, 0.486]). Together, these results show that coordinated immune responses are more likely within pathogen-specific antigenic groups than across pathogens.

Altogether, these findings demonstrate that the highly heterogeneous immune response after vaccination reflects a composite of individual, schedule-dependent, and vaccine-specific characteristics.

##### The seasonal immune responses and their latitudinal gradients remained consistent when considering seronegative individuals in the rotavirus studies

Given the bimodal distribution of rotavirus immunogenicity, we performed a sensitivity analysis comparing our main results, which were restricted to seronegative individuals, with models that included all participants regardless of baseline serostatus. We found that the estimated seasonal patterns, including peak amplitude and timing, remained consistent between the two approaches (Supplementary figures 11 and 12). This indicates that the observed seasonality is not an artifact of excluding baseline seropositivity but rather reflects a robust signal across the entire rotavirus vaccination cohort.

### Supplementary figures

**Sfig 1. Flowchart of study selection and participant inclusion.** Systematic selection process for the randomized clinical trials (RCTs) and participants included in the analysis.

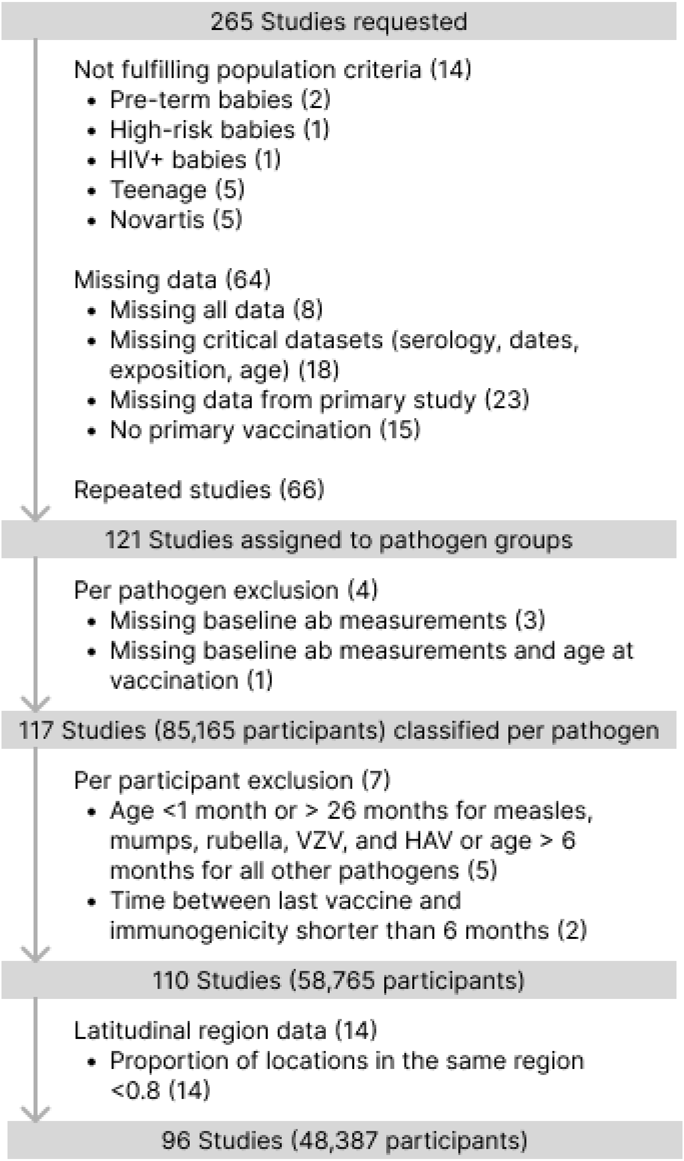

**Sfig 2. Spatiotemporal distribution of vaccination events.** Weekly histograms show the vaccination calendar week for study participants for each pathogen, colored by latitudinal classification.

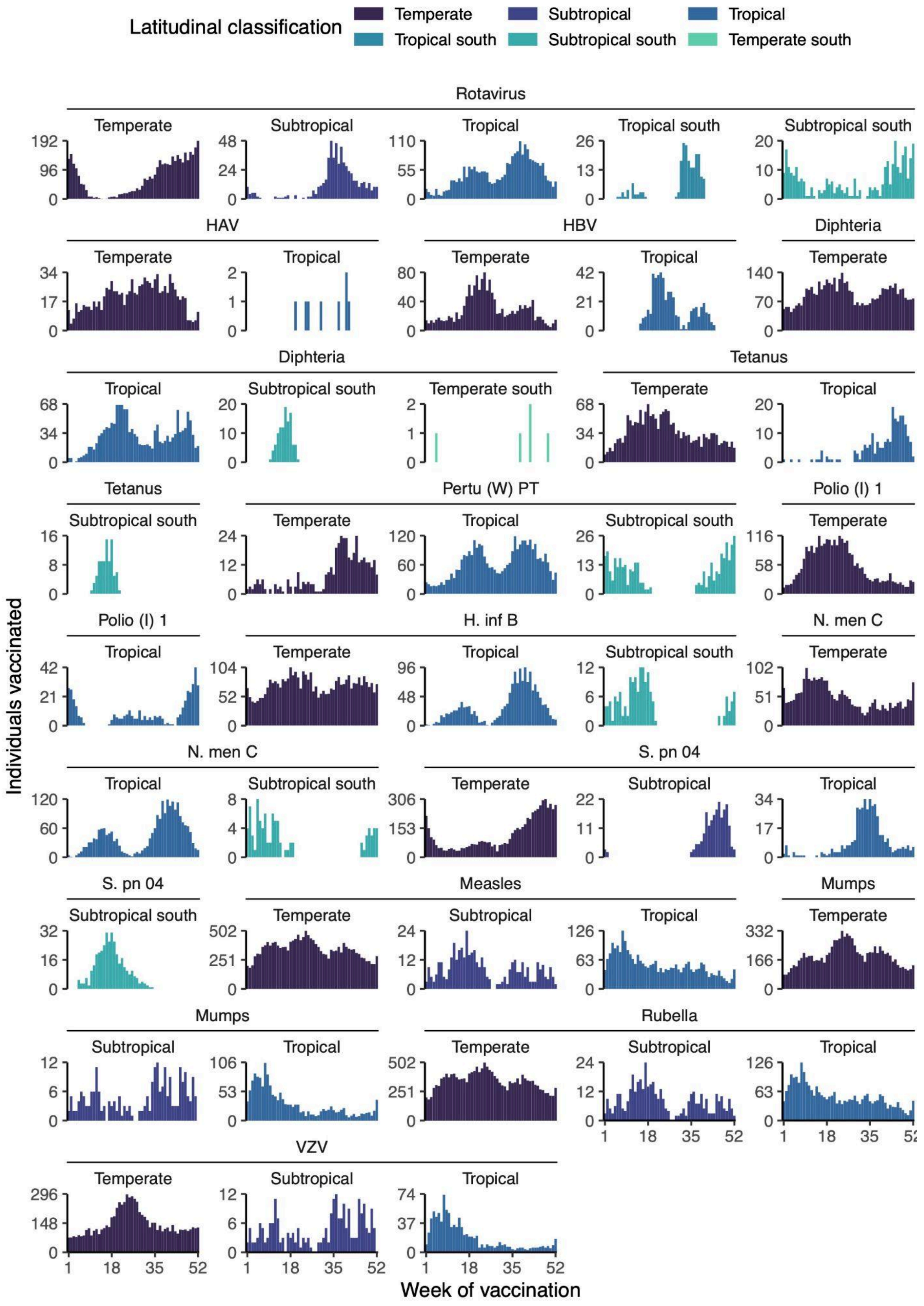

**Sfig 3. Model validation and bias assessment across pathogen-antigen subsets.** The scatter plots illustrate the mean absolute bias for peak amplitude and peak timing obtained from posterior predictive checks. To evaluate the robustness of seasonal estimates considering the temporal data gaps, datasets were simulated from the best-fitted models and refitted to assess parameter recovery. Horizontal and vertical lines represent the predefined exclusion thresholds of 20% for amplitude bias and 7 weeks for phase bias.

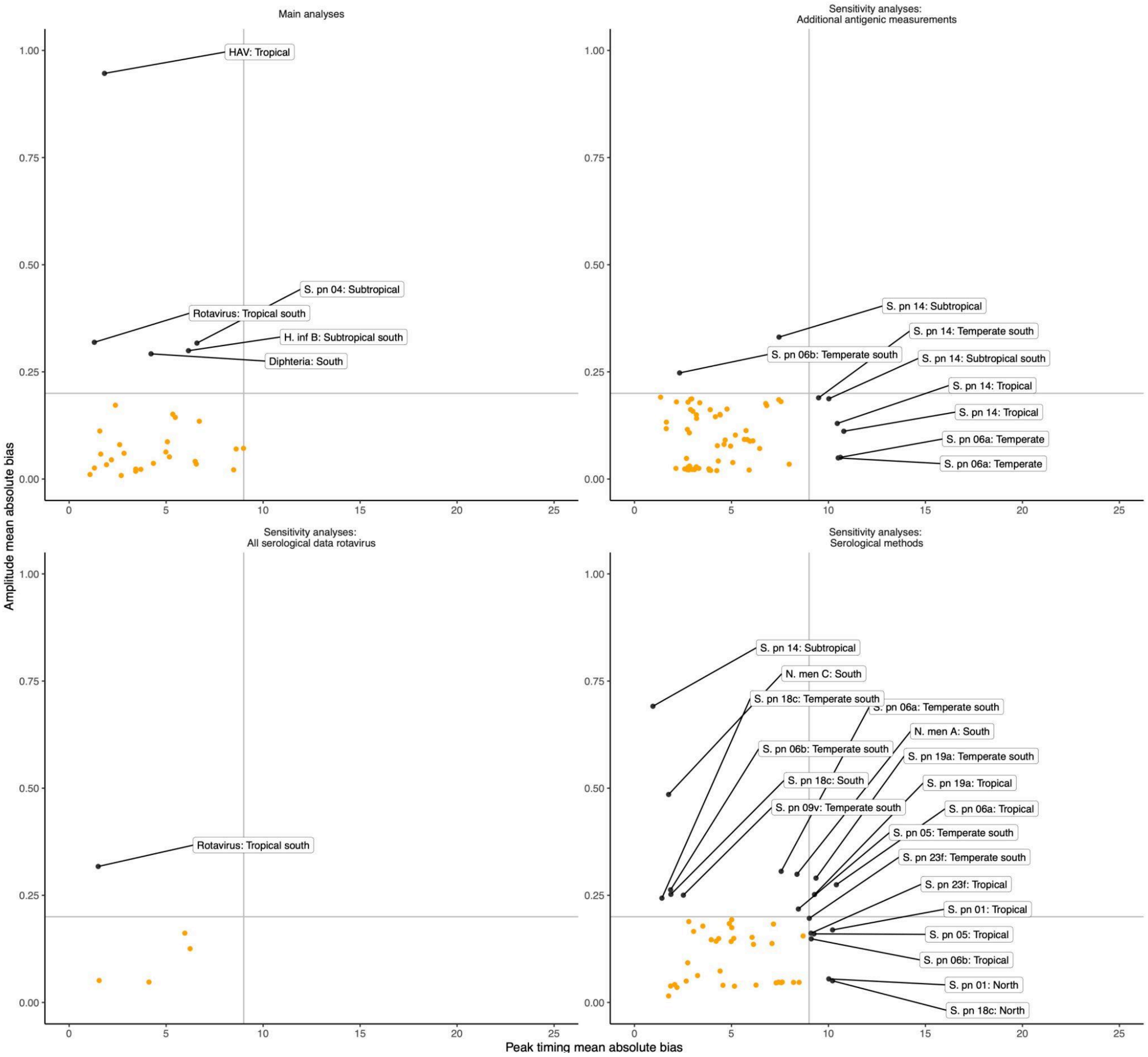

**Sfig 4. Comparison of seasonal model forms.** Seasonal immunogenicity patterns estimated using generalized additive models with cyclic splines (red) compared with cosine model fits (black), showing highly similar shapes with a single dominant seasonal peak. The panels show posterior conditional effects of vaccination week on immunogenicity for selected pathogens and latitudes, with curves representing posterior draws and rug marks indicating observed vaccination weeks.

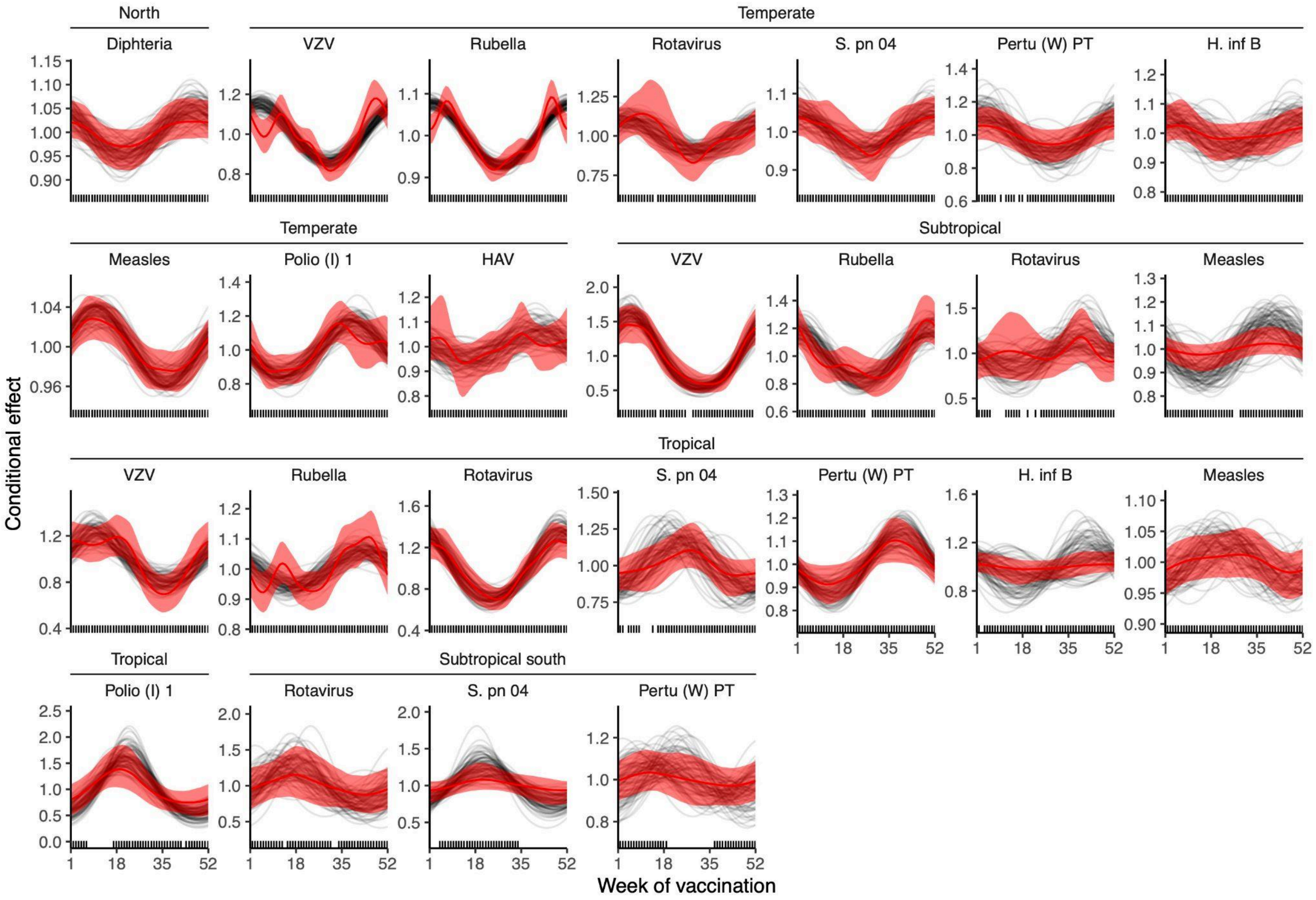

**Sfig 5. Global distribution, demographic characteristics, and immunogenicity profiles of pediatric vaccination trials included in the study for sensitivity analyses with additional antigens.** A) Total number of vaccinated individuals per pathogen; color intensity indicates contributions from individual studies, and numbers denote the number of RCTs. B) Age at first vaccination, with point size proportional to the fraction of participants vaccinated at each age (months). C) Sex distribution of participants by pathogen. D) For each pathogen, weekly distribution of vaccinations across the calendar year, density of post-vaccination immunogenicity measurements, and geographic distribution of study sites (point size proportional to the number of vaccinated individuals; colors indicate latitudinal classification).

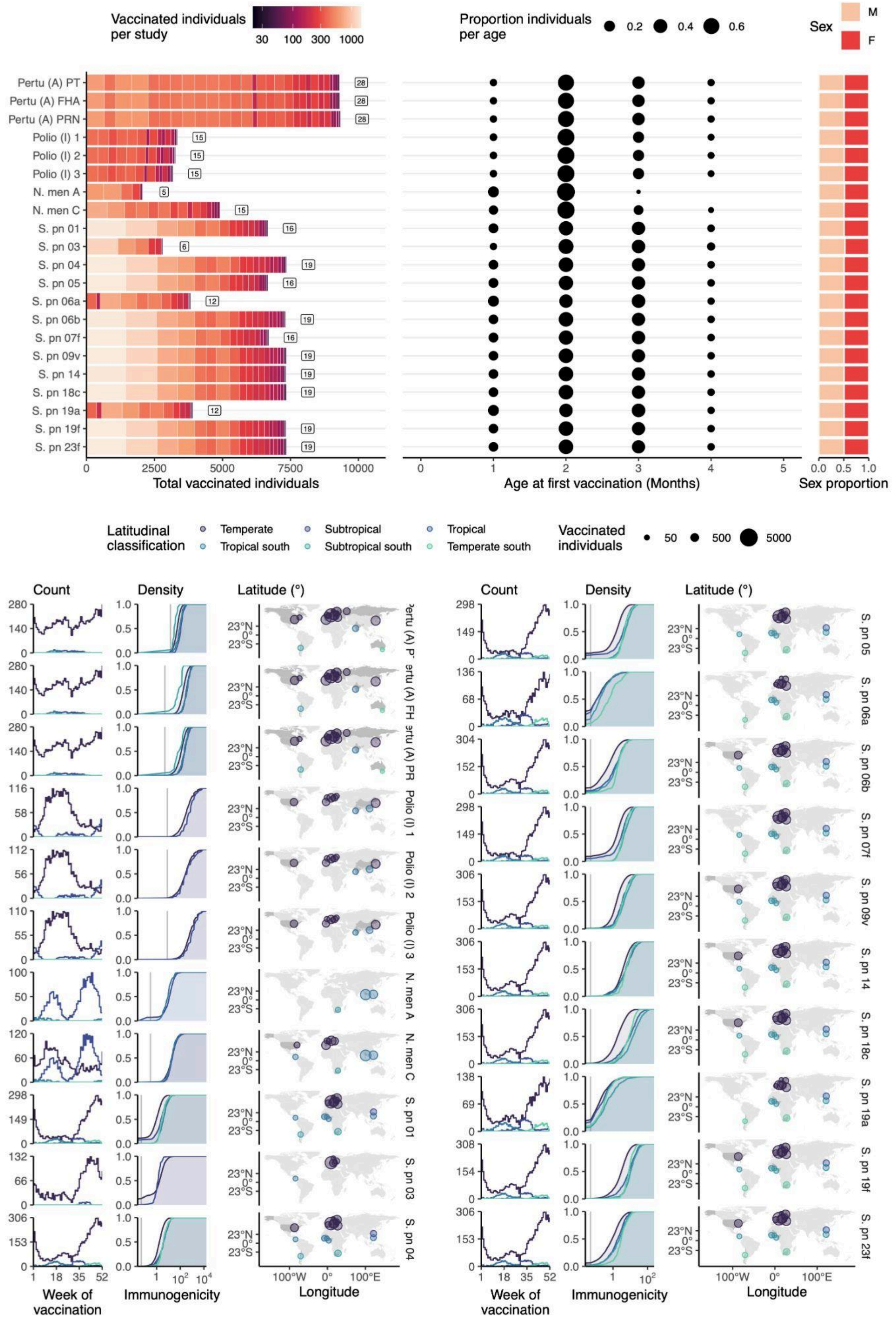

**Sfig 6. Incorporating vaccination date improves the predictive performance of vaccine immunogenicity models across pathogens and latitudes in sensitivity analyses with additional antigens.** Comparison of baseline, seasonal, and seasonal-by-latitude models for each antigen. Top panels show differences in WAIC relative to the best-fitting model (ΔWAIC), with points indicating model variants (baseline, seasonality only, and seasonality stratified by 2–6 latitudinal groups); lower values indicate better predictive performance. The vertical grey line denotes a difference of 2 points. Bottom panels show the corresponding empirical cumulative distribution functions of observed (solid) and model-predicted (dashed) post-vaccination immunogenicity, colored by antigen. Vertical lines denote serological thresholds.

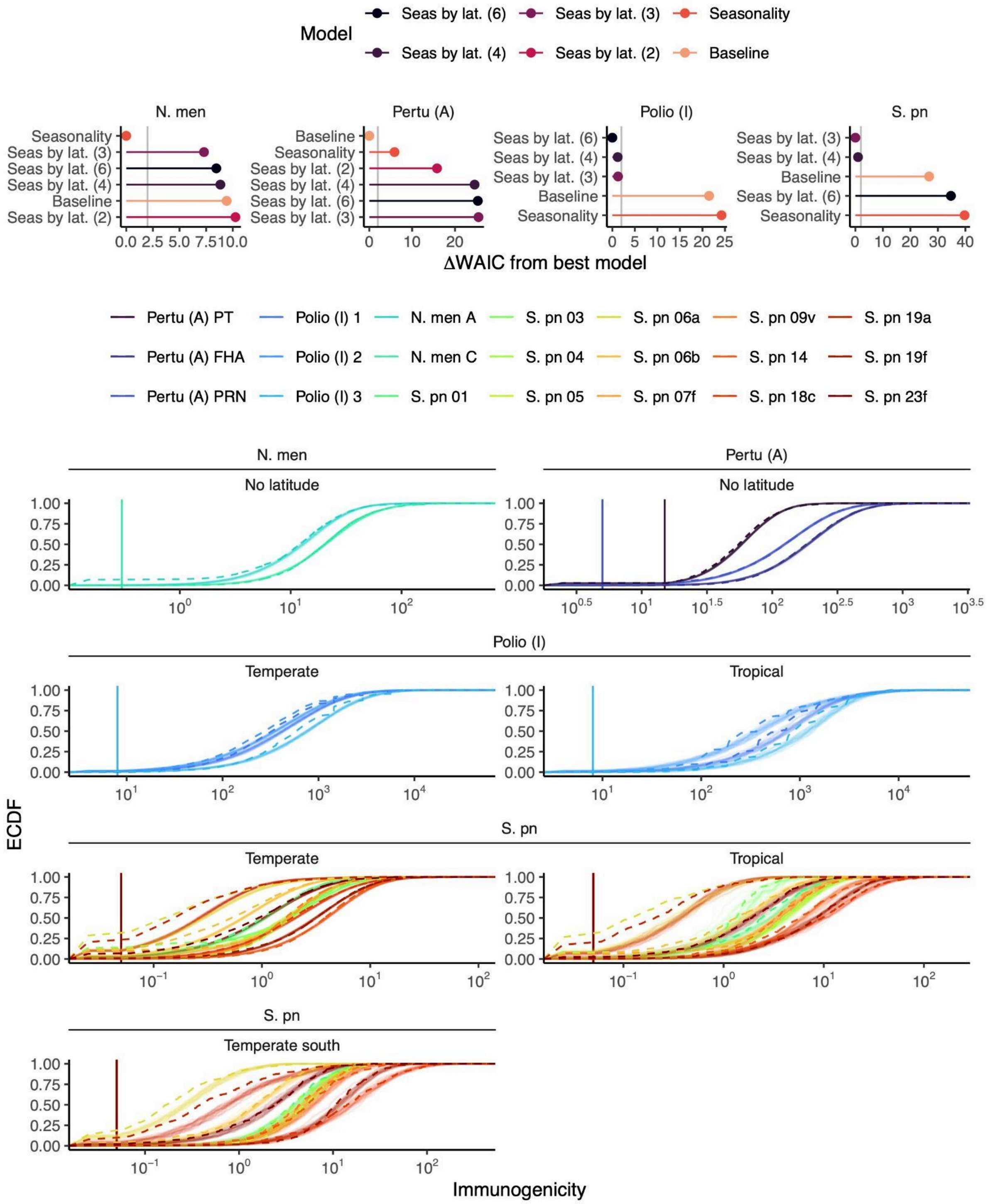

**Sfig 7. Latitudinal gradients in the seasonal timing and amplitude of the vaccine immunogenicity in sensitivity analyses with additional antigens.** Top panels show posterior estimates of seasonal amplitude (left) and peak timing (right, week of year) from the best-fitting seasonal-by-latitude models for each antigen and latitudinal group (points, medians; lines, 95% credible intervals; color indicates posterior phase density). Bottom panels show posterior conditional effects of vaccination week on immunogenicity for selected pathogens and latitudes, with curves representing posterior draws and rug marks indicating observed vaccination weeks.

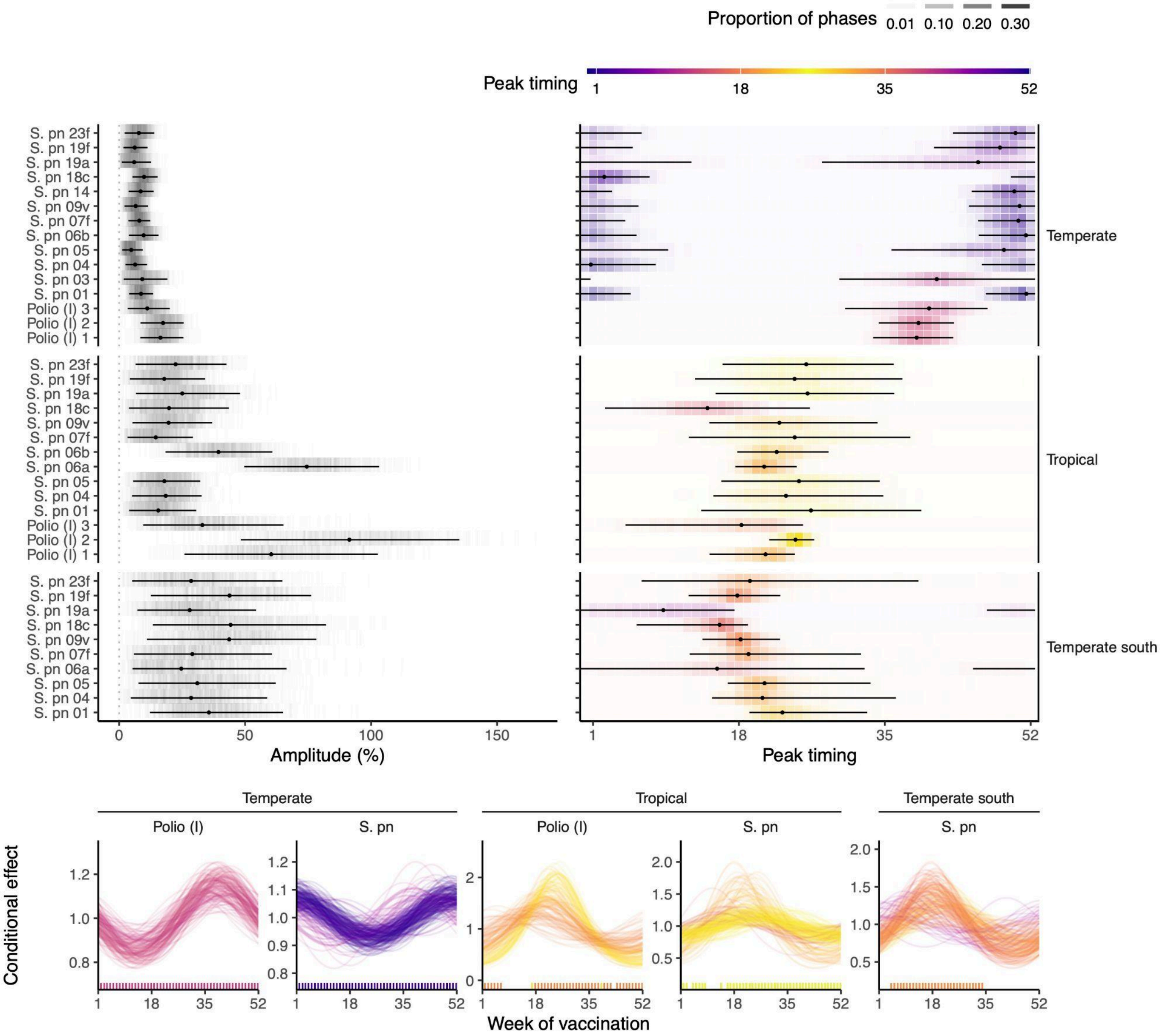

**Sfig 8. Global distribution, demographic characteristics, and immunogenicity profiles of pediatric vaccination trials included in the study for sensitivity analyses with functional antibody measurements.** A) Total number of vaccinated individuals per pathogen; color intensity indicates contributions from individual studies, and numbers denote the number of RCTs. B) Age at first vaccination, with point size proportional to the fraction of participants vaccinated at each age (months). C) Sex distribution of participants by pathogen. D) For each pathogen, weekly distribution of vaccinations across the calendar year, density of post-vaccination immunogenicity measurements, and geographic distribution of study sites (point size proportional to the number of vaccinated individuals; colors indicate latitudinal classification).

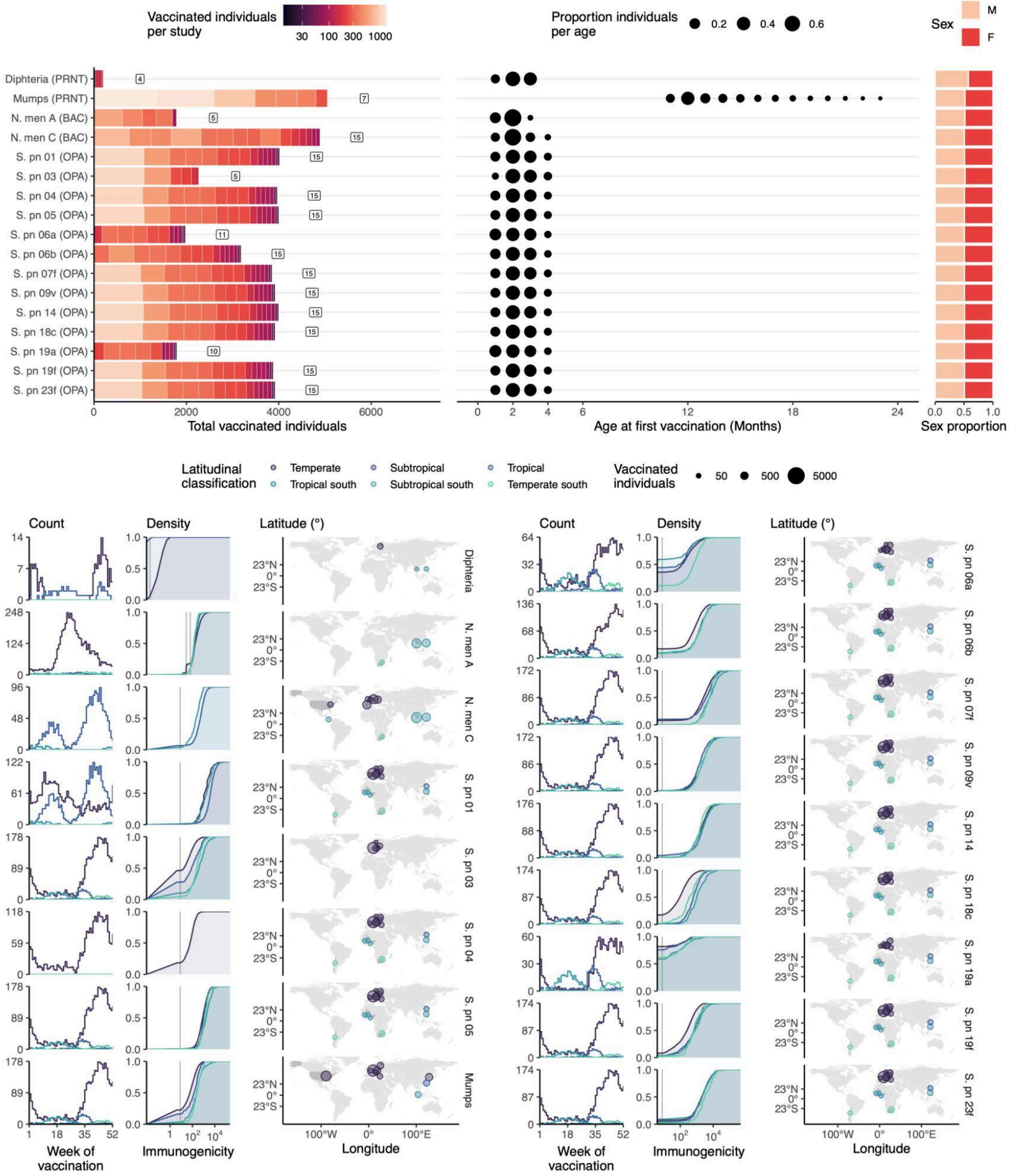

**Sfig 9. Incorporating vaccination date improves the predictive performance of vaccine immunogenicity models across pathogens and latitudes in sensitivity analyses with functional antibody measurements.** Comparison of baseline, seasonal, and seasonal-by-latitude models for each antigen. Top panels show differences in WAIC relative to the best-fitting model (ΔWAIC), with points indicating model variants (baseline, seasonality only, and seasonality stratified by 2–6 latitudinal groups); lower values indicate better predictive performance. The vertical grey line denotes a difference of 2 points. Bottom panels show the corresponding empirical cumulative distribution functions of observed (solid) and model-predicted (dashed) post-vaccination immunogenicity, colored by antigen. Vertical lines denote serological thresholds.

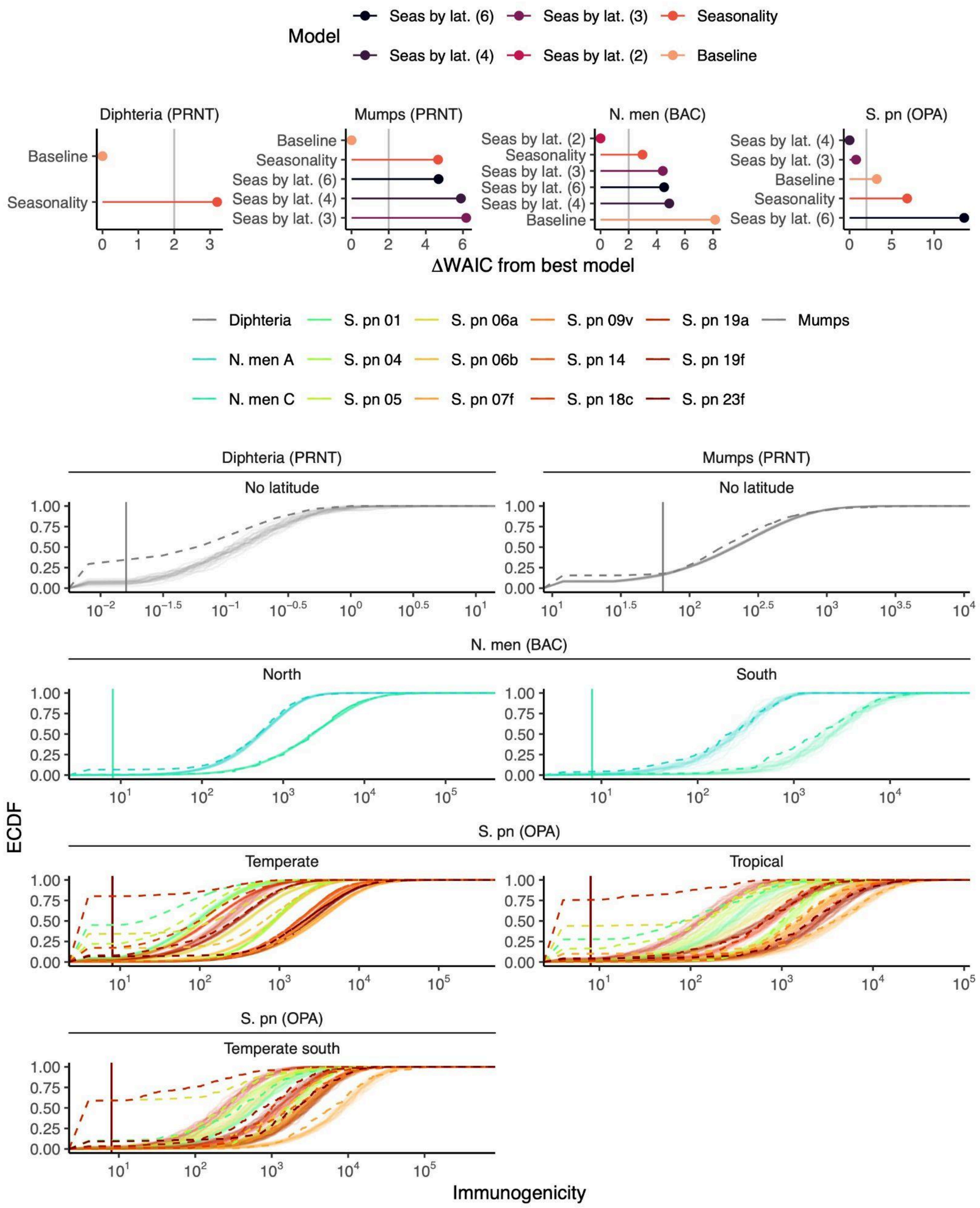

**Sfig 10. Latitudinal gradients in the seasonal timing and amplitude of the vaccine immunogenicity in sensitivity analyses with functional antibody measurements.** Top panels show posterior estimates of seasonal amplitude (left) and peak timing (right, week of year) from the best-fitting seasonal-by-latitude models for each antigen and latitudinal group (points, medians; lines, 95% credible intervals; color indicates posterior phase density). Bottom panels show posterior conditional effects of vaccination week on immunogenicity for selected pathogens and latitudes, with curves representing posterior draws and rug marks indicating observed vaccination weeks.

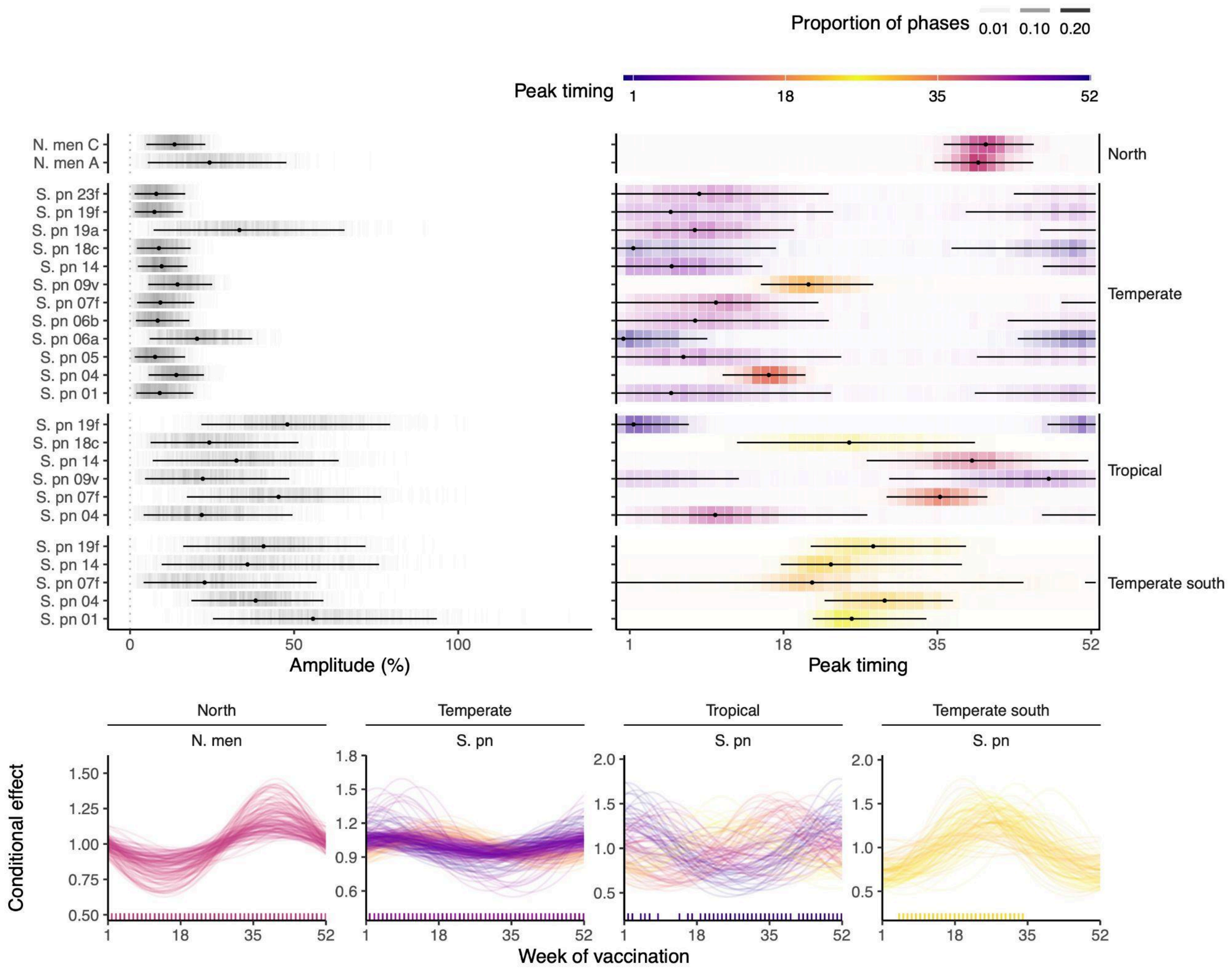

**Sfig 11. Incorporating vaccination date improves the predictive performance of vaccine immunogenicity models across pathogens and latitudes in sensitivity analyses for rotavirus studies that included seronegative individuals.** Comparison of baseline, seasonal, and seasonal-by-latitude models. The left panel shows differences in WAIC relative to the best-fitting model (ΔWAIC), with points indicating model variants (baseline, seasonality only, and seasonality stratified by 2–6 latitudinal groups); lower values indicate better predictive performance. The vertical grey line denotes a difference of 2 points. The right panel shows the corresponding empirical cumulative distribution functions of observed (solid) and model-predicted (dashed) post-vaccination immunogenicity, colored by antigen. The vertical grey line denotes serological thresholds.

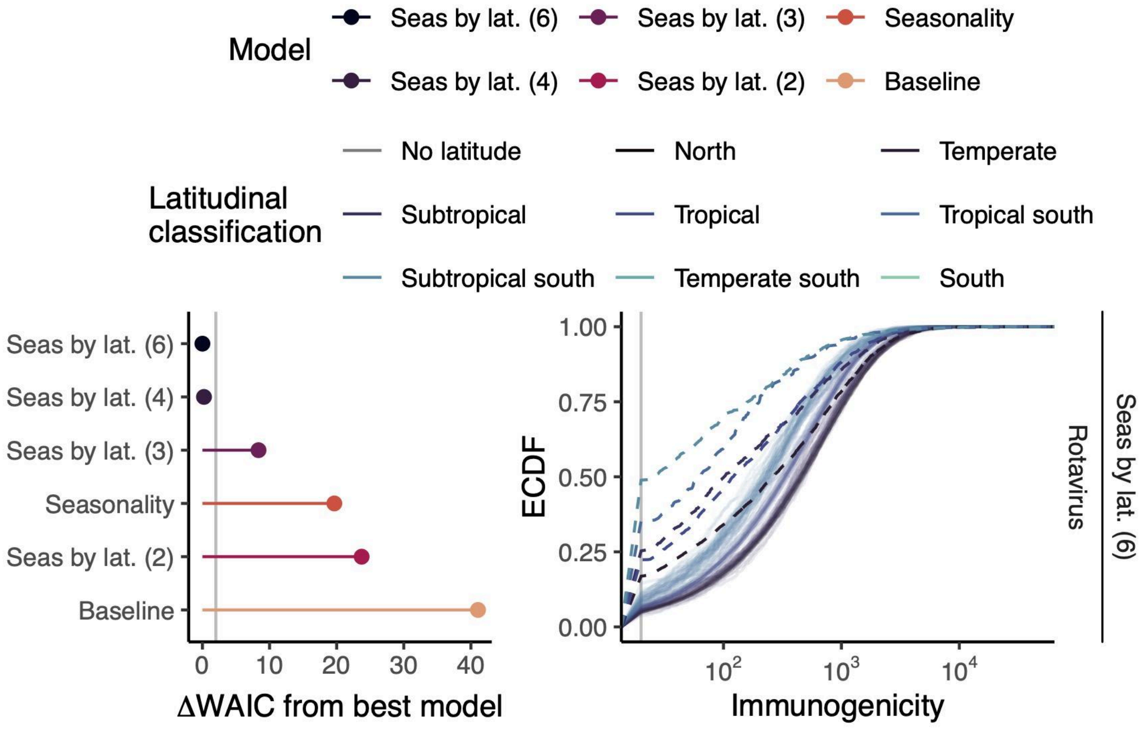

**Sfig 12. Latitudinal gradients in the seasonal timing and amplitude of the vaccine immunogenicity in sensitivity analyses for rotavirus studies that included seronegative individuals.** Top panels show posterior estimates of seasonal amplitude (left) and peak timing (right, week of year) from the best-fitting seasonal-by-latitude models for each model (points, medians; lines, 95% credible intervals; color indicates posterior phase density). Bottom panels show posterior conditional effects of vaccination week on immunogenicity for selected pathogens and latitudes, with curves representing posterior draws and rug marks indicating observed vaccination weeks.

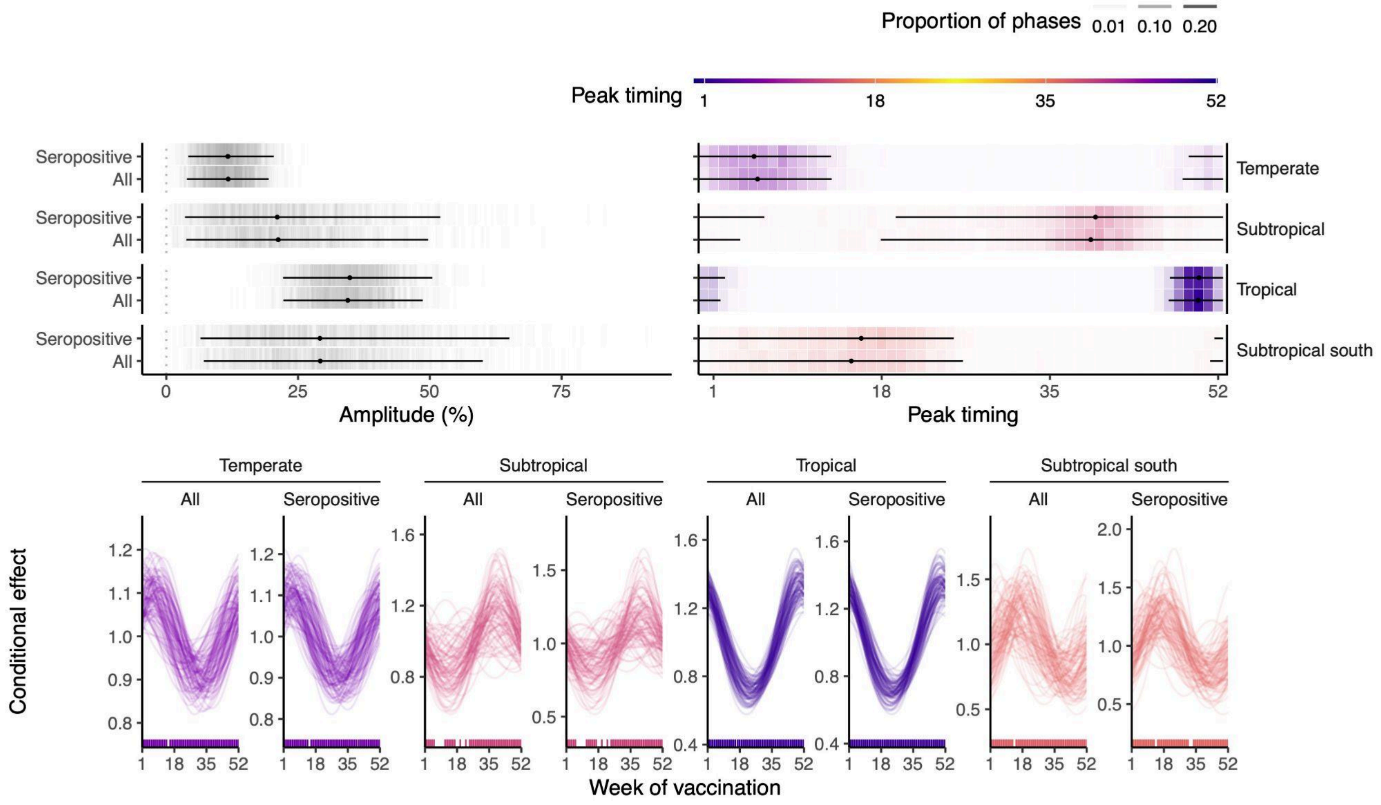

**Sfig 13. Incorporating the date of birth did not outperform models that included the date of vaccination across pathogens.** Comparison of seasonal and seasonal-by-latitude models with date of birth with the best model with date of vaccination. The panels show differences in WAIC relative to the best-fitting model, with points indicating model variants (seasonality only and seasonality stratified by 2–6 latitudinal groups); lower values indicate better predictive performance. The vertical grey line denotes a difference of 2 points.

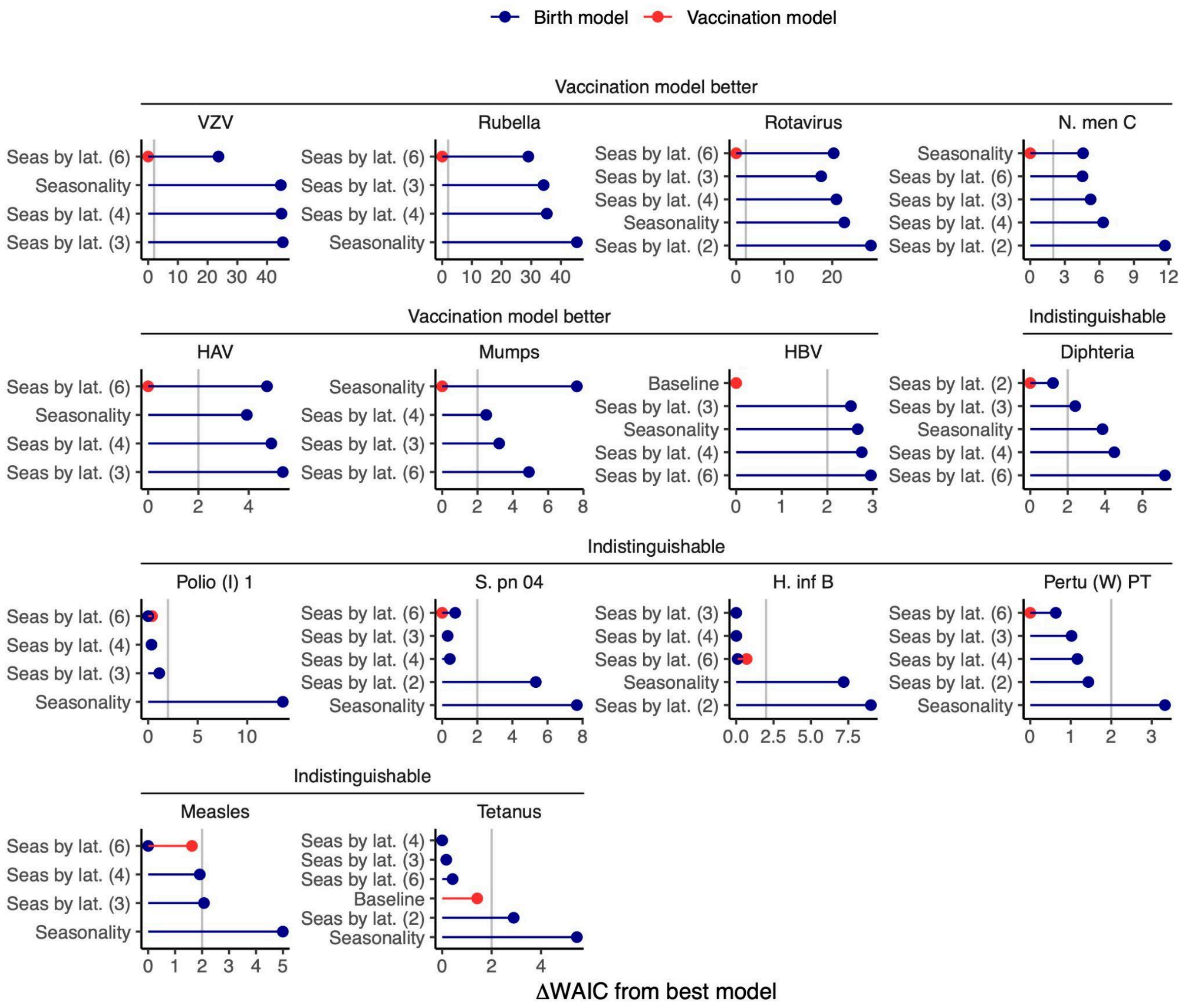

### Supplementary tables

**Stable 1 Randomized controlled trials included in the study evaluating vaccine immunogenicity.** The table details the specific inclusion status of each study; "Partial exclusion" indicates that while the study was included in the broader analysis, data for specific antigens were excluded.

| Study ID | Study title | Link | Exclusion | Reason | Inclusion per pathogen |
|---|---|---|---|---|---|
| 217744-076 | An open, multicenter, phase IV clinical trial to assess the immunogenicity and reactogenicity of three doses of GSK Biologicals' combined DTPa-HBV-IPV/Hib vaccine in healthy infants at 2, 4 and 6 months of age, when co-administered with Wyeth-Lederle's meningococcal group C conjugate vaccine. | NA | Included | Included | Diphtheria, HBV, HiB, IPV (1, 2, 3), Nmen (C), Pertussis-a (FHA, PRN, PT), Tetanus |
| 711202-001 | Evaluate immunogenicity, reactogenicity, safety of GSK Biologicals' MenC-TT vaccine (2 formulations) given with Infanrix hexa® + GSK Biologicals' Hib MenC-TT vaccine (2 formulations) given with Infanrix penta® to infants in mths 3,4,5 of life | http://clinicaltrials.gov/show/NCT00135486 | Included | Included | Diphtheria, HBV, HiB, IPV (1, 2, 3), Nmen (C), Pertussis-a (FHA, PRN, PT), Tetanus |
| 811936-001 | Phase 2, open, randomized, controlled study to demonstrate the non-inferiority of the meningococcal serogroup C immune response and the superiority of the Hib immune response of GSK Biologicals' Haemophilus influenzae type b-meningococcal C-TT conjugate vaccine administered with Infanrix™ penta versus Meningitec™ administered with Infanrix™ | NA | Included | Included | Diphtheria, HBV, HiB, IPV (1, 2, 3), Nmen (C), Pertussis-a (FHA, PRN, PT), Tetanus |
| 217744-078 | An open, multicentre, phase IV clinical trial to assess the immunogenicity and reactogenicity of GSK Biologicals' combined DTPa-HBV-IPV/Hib vaccine, when co-administered at 3-4-5 Mth of age with Wyeth-Lederle's seven-valent pneumococcal conjugate vaccine at a different injection site during the same visit | NA | Included | Included | Diphtheria, HBV, HiB, IPV (1, 2, 3), Pertussis-a (FHA, PRN, PT), Sp (04, 06b, 09v, 14, 18c, 19f, 23f), Tetanus |
| 217744-070 | An open clinical study to assess the immunogenicity and safety of GSK Bio's DTPa-HBV-IPV vaccine mixed in one syringe with Hib vaccine, as a primary vaccination course to pre-term infants(<37 weeks) at 2, 4 and 6 months of age in comparison with infants born after normal gestation period | http://clinicaltrials.gov/show/NCT00366366 | Partial exclusion of RCT group | Pre-term babies RCT group | Diphtheria, HBV, HiB, IPV (1, 2, 3), Pertussis-a (FHA, PRN, PT), Tetanus |
| 208108-091 | A phase II, double-blind, randomized study to compare the immunogenicity, safety and reactogenicity of GlaxoSmithKline (GSK) Biologicals' Tritanrix™-HepB/Hib2.5 to GSK Biologicals' Tritanrix™-HepB/Hiberix™ when administered as a three-dose primary vaccination course to healthy infants at 6, 10 and 14 weeks of age. A dose of unconjugated Hib vaccine (plain PRP booster) will be administered at the age of 10 months to 50% of the subjects | http://clinicaltrials.gov/show/NCT01061541 | Partial exclusion for Polio | No Ab (Polio) No primary (Polio) | Diphtheria, HBV, HiB, Pertussis-w (PT), Tetanus |
| 101222 | Study to demonstrate the non-inferiority of GSK Biologicals' DTPw-HBV/Hib Kft. vaccine compared to GSK Biologicals' Tritanrix™-HepB/Hiberix™ vaccine and to separate administration of DTPw-HBV Kft. and Hiberix™ vaccines with respect to the immunogenicity of all antigens, when administered to healthy infants. | NA | Partial exclusion for Tetanus and HiB | No Ab baseline (Tetanus, HiB) | Diphtheria, HBV, Pertussis-w (PT) |
| 103974 | Demonstrate non-inferiority of Men-C immune response of Hib-MenC with Infanrix™-IPV versus a licensed Men-C vaccine with Pediacel™ when given at 2, 3, 4 months and the immunogenicity of Hib-MenC when given as a booster dose at 12-15 months | http://clinicaltrials.gov/show/NCT00258700 | Included | Included | Diphtheria, HiB, IPV (1, 2, 3), Measles, Nmen (C), Mumps, Pertussis-a (FHA, PRN, PT), Rubella, Tetanus |
| 101858 | Evaluate Immuno and Safety of GSKBiologicals' HibMenCYTT vs Licensed Hib Conjugate Vaccine, Each Coadministered With Pediarix® and Prevnar®, in Healthy Infants. An Exploratory Control Group Will Receive Licensed Menomune® at 3 to 5 years | http://clinicaltrials.gov/show/NCT00129129 | Partial exclusion for HBV | No primary (HBV) | Diphtheria, HiB, IPV (1, 2, 3), Nmen (C), Pertussis-a (FHA, PRN, PT), Sp (04, 06b, 09v, 14, 18c, 19f, 23f), Tetanus |
| 113994 | Safety, reactogenicity & immunogenicity of GSK Biologicals' pneumococcal vaccine 2189242A when co-administered with DTPa-HBV-IPV/Hib vaccine in healthy infants | http://clinicaltrials.gov/show/NCT01204658 | Partial exclusion for HBV | No primary (HBV) | Diphtheria, HiB, IPV (1, 2, 3), Pertussis-a (FHA, PRN, PT), Sp (01, 03, 04, 05, 06a, 06b, 07f, 09v, 14, 18c, 19a, 19f, 23f), Tetanus |
| 347414-023 | A phase II, single-blind, randomized, controlled study to evaluate the immunogenicity and safety of four different formulations of an investigational vaccination regimen when given intramuscularly as primary vaccination in infants at 3, 4 ½ and 6 months of age. | NA | Included | Included | Diphtheria, HiB, IPV (1, 2, 3), Pertussis-a (FHA, PRN, PT), Sp (01, 03, 04, 05, 06b, 07f, 09v, 14, 18c, 19f, 23f), Tetanus |

| Study ID | Study title | Link | Exclusion | Reason | Inclusion per pathogen |
|---|---|---|---|---|---|
| 217744-076 | An open, multicenter, phase IV clinical trial to assess the immunogenicity and reactogenicity of three doses of GSK Biologicals' combined DTPa-HBV-IPV/Hib vaccine in healthy infants at 2, 4 and 6 months of age, when co-administered with Wyeth-Lederle's meningococcal group C conjugate vaccine. | NA | Included | Included | Diphtheria, HBV, HiB, IPV (1, 2, 3), Nmen (C), Pertussis-a (FHA, PRN, PT), Tetanus |
| 711202-001 | Evaluate immunogenicity, reactogenicity, safety of GSK Biologicals' MenC-TT vaccine (2 formulations) given with Infanrix hexa® + GSK Biologicals' Hib MenC-TT vaccine (2 formulations) given with Infanrix penta® to infants in mths 3,4,5 of life | http://clinicaltrials.gov/show/NCT00135486 | Included | Included | Diphtheria, HBV, HiB, IPV (1, 2, 3), Nmen (C), Pertussis-a (FHA, PRN, PT), Tetanus |
| 217744-085 | A phase III, open labeled, randomized, multicenter, clinical study of the safety and immunogenicity of a primary series of GlaxoSmithKline Biologicals' (GSK Biologicals') DTaP-HepBIPV candidate vaccine coadministered with HibTITER® and Prevnar® to healthy infants at 2, 4, and 6 months of age as compared to the separate administration of Infanrix® + Engerix-B® + IPOL® + HibTITER + Prevnar and to GSK Biologicals' DTaP-HepB-IPV candidate vaccine coadministered with HibTITER | NA | Partial exclusion for HBV | No primary (HBV) | Diphtheria, HiB, IPV (1, 2, 3), Pertussis-a (FHA, PRN, PT), Sp (04, 06b, 09v, 14, 18c, 19f, 23f), Tetanus |
| 114260 | Immunogenicity and safety of GlaxoSmithKline Biologicals' DTPa-IPV/Hib (Infanrix™-IPV+Hib) vaccine in healthy Korean infants | http://clinicaltrials.gov/show/NCT01309646 | Partial exclusion for Rotavirus and Sp | No Ab (Rotavirus, Sp) | Diphtheria, HiB, IPV (1, 2, 3), Pertussis-a (FHA, PRN, PT), Tetanus |
| 105553 | Assess lot-to-lot consistency of 3 lots (double blind design) of GlaxoSmithKline Biologicals' 10-valent pneumococcal vaccine and evaluate non-inferiority to Prevenar™ (single blind design) when administered as 3-dose primary immunization course before 6 months of age | http://clinicaltrials.gov/show/NCT00307554 | Partial exclusion for HBV and Polio | No primary routine (HBV),<br>No Ab baseline (Polio) | Diphtheria, HiB, Measles, Mumps, Pertussis-a (FHA, PRN, PT), Rubella, Sp (01, 04, 05, 06a, 06b, 07f, 09v, 14, 18c, 19a, 19f, 23f), Tetanus, VZV |
| 759346-001 | A phase II, open, randomized, controlled study to evaluate the immunogenicity of investigational vaccinationregimens administered as a three dose primary vaccination course at 6, 10 and 14 weeks of age. | NA | Partial exclusion for HBV | No primary (HBV) | Diphtheria, HiB, Nmen (A, C), Pertussis-w (PT), Tetanus |
| 104733 | Demonstrate lot-to-lot consistency of final production method of GSK Biologicals' Hib-MenAC vaccine mixed extemporaneously with Tritanrix™-HepB & demonstrate its non-inferiority vs Tritanrix™-HepB/Hiberix™ in healthy infants at 2, 4 and 6 months | http://clinicaltrials.gov/show/NCT00197275 | Partial exclusion for HBV and Polio | No Ab baseline (HBV, Polio) | Diphtheria, HiB, Nmen (A, C), Pertussis-w (PT), Tetanus |
| 347414-017 | A multinational, randomised, controlled, single-blind, phase-II study to evaluate the safety and immunogenicity of two investigational vaccination regimens versus a licensed Haemophilus influenzae type b (Hib) conjugate vaccine (Hiberix) given concomitantly with DTPw-HBV vaccine in a separate injection of DTPw-HBV to infants at 2, 4, and 6 months of age | NA | Partial exclusion for HBV | No primary (HBV) | Diphtheria, HiB, Nmen (C), Pertussis-w (PT), Sp (01, 03, 04, 05, 06b, 07f, 09v, 14, 18c, 19f, 23f), Tetanus |
| 107017 | Multicentre study to assess the effect of prophylactic antipyretic treatment on the rate of febrile reactions following concomitant administration of GSK Biologicals' 10-valent pneumococcal conjugate, Infanrix hexa and Rotarix vaccines | http://clinicaltrials.gov/show/NCT00370318 | Partial exclusion for HBV and Polio | No primary(HBV),<br>No Ab baseline (Polio) | Diphtheria, HiB, Pertussis-a (FHA, PRN, PT), Rotavirus, Sp (01, 04, 05, 06a, 06b, 07f, 09v, 14, 18c, 19a, 19f, 23f), Tetanus |
| 106208 | To assess reactogenicity and immunogenicity of GSK Biologicals' 10-valent pneumococcal conjugate vaccine, when co-administered with GSK Biologicals' DTPa-HBV-IPV/Hib vaccine (Infanrix™ hexa) at 2, 4 and 6 months of age. | http://clinicaltrials.gov/show/NCT00338351 | Partial exclusion for HBV and Polio | No Ab baseline (HBV, polio) | Diphtheria, HiB, Pertussis-a (FHA, PRN, PT), Sp (01, 04, 05, 06a, 06b, 07f, 09v, 14, 18c, 19a, 19f, 23f), Tetanus |
| 112921 | Impact of immediate or delayed prophylactic antipyretic treatment on the immunogenicity, reactogenicity and safety of GlaxoSmithKline Biologicals' pneumococcal vaccine 1024850A and the co-administered DTPa-combined vaccines | http://clinicaltrials.gov/show/NCT01235949 | Partial exclusion for HBV and Polio | No primary (HBV),<br>No Ab baseline (Polio) | Diphtheria, HiB, Pertussis-a (FHA, PRN, PT), Sp (01, 04, 05, 06a, 06b, 07f, 09v, 14, 18c, 19a, 19f, 23f), Tetanus |
| 105554 | Phase IIIa randomized, controlled study to assess the immunogenicity of GlaxoSmithKline (GSK) Biologicals' 10-valent pneumococcal conjugate vaccine, when administered as a 3-dose primary immunization course before 6 months of age | http://clinicaltrials.gov/show/NCT00307541 | Partial exclusion for HBV and Polio | No Ab baseline (HBV, Polio) | Diphtheria, HiB, Pertussis-a (FHA, PRN, PT), Sp (01, 04, 05, 06b, 07f, 09v, 14, 18c, 19f, 23f), Tetanus |
| 107007 | To assess the safety, reactogenicity and immunogenicity of GSK Biologicals' pneumococcal conjugate vaccine compared to Prevenar™, co-administered with DTPw-HBV/Hib & OPV or IPV vaccines as a 3-dose primary immunization course during the first 6 months of age | http://clinicaltrials.gov/show/NCT00344318 | Partial exclusion for HBV and Polio | No primary (HBV),<br>No Ab baseline (Polio) | Diphtheria, HiB, Pertussis-w (PT), Sp (01, 04, 05, 06a, 06b, 07f, 09v, 14, 18c, 19a, 19f, 23f), Tetanus |
| 104871 | A multicentric study to compare the immunogenicity, safety & reactogenicity of GSK Biologicals' DTPa-IPV vaccine vs. co-administration of GSK's DTPa vaccine & Sanofi-Pasteurs' IPV vaccine at different injection sites, to healthy children | http://clinicaltrials.gov/show/NCT00290342 | Included | Included | Diphtheria, IPV (1, 2, 3), Pertussis-a (FHA, PRN, PT), Tetanus |
| 106786 | A study to evaluate the immunogenicity and safety of a new formulation of GSK Biologicals' DTPa-HBV-IPV/Hib vaccine as compared to the currently licensed Infanrix hexa in healthy infants (2,3,4 M) | http://clinicaltrials.gov/show/NCT00376779 | Partial exclusion for HBV, HiB, and Polio | No Ab baseline (HBV, HiB, polio) | Diphtheria, Pertussis-a (FHA, PRN, PT), Tetanus |

| Study ID | Study title | Link | Exclusion | Reason | Inclusion per pathogen |
|---|---|---|---|---|---|
| 217744-076 | An open, multicenter, phase IV clinical trial to assess the immunogenicity and reactogenicity of three doses of GSK Biologicals' combined DTPa-HBV-IPV/Hib vaccine in healthy infants at 2, 4 and 6 months of age, when co-administered with Wyeth-Lederle's meningococcal group C conjugate vaccine. | NA | Included | Included | Diphtheria, HBV, HiB, IPV (1, 2, 3), Nmen (C), Pertussis-a (FHA, PRN, PT), Tetanus |
| 711202-001 | Evaluate immunogenicity, reactogenicity, safety of GSK Biologicals' MenC-TT vaccine (2 formulations) given with Infanrix hexa® + GSK Biologicals' Hib MenC-TT vaccine (2 formulations) given with Infanrix penta® to infants in mths 3,4,5 of life | http://clinicaltrials.gov/show/NCT00135486 | Included | Included | Diphtheria, HBV, HiB, IPV (1, 2, 3), Nmen (C), Pertussis-a (FHA, PRN, PT), Tetanus |
| 201330 | Immunogenicity and safety study of GSK Biologicals' combined diphtheria-Tetanusus-acellular pertussis-hepatitis B-inactivated polio-virus and Haemophilus influenzae type b vaccine (Infanrix hexa™) (217744) in healthy infants born to mothers vaccinated with Boostrix™ during pregnancy or immediately post-delivery | http://clinicaltrials.gov/show/NCT02422264 | Partial exclusion for HBV, HiB, Polio, and Sp | No Ab baseline (HBV, HiB, Polio, Sp) | Diphtheria, Pertussis-a (FHA, PRN, PT), Tetanus |
| 101197 | An open, multicentric, phase IV clinical trial to assess the immunogenicity and reactogenicity of GlaxoSmithKline Biologicals' combined diphtheria-Tetanusus-acellular pertussis vaccine (Infanrix™) administered to healthy infants at 2, 4 and 6 months of age. | NA | Included | Included | Diphtheria, Pertussis-a (FHA, PRN, PT), Tetanus |
| 104021 | A phase III, partially blind, randomized study to evaluate the immunogenicity, safety and reactogenicity of GlaxoSmithKline (GSK) Biologicals' Tritanrix™-HepB and GSK Biologicals Kft's DTPw-HBV vaccines as compared to concomitant administration of Commonwealth Serum Laboratory's (CSL's) DTPw (Triple Antigen™) and GSK Biologicals' HBV (Engerix™-B), when co-administered with GSK Biologicals' oral live attenuated human rotavirus (HRV) vaccine, to healthy infants at 3, 4½ and 6 months of age, after a birth dose of hepatitis B vaccine. | http://clinicaltrials.gov/show/NCT00158756 | Partial exclusion for Polio | No Ab baseline (Polio) | Diphtheria, Pertussis-w (PT), Rotavirus, Tetanus |
| 114056 | Immunogenicity, safety and reactogenicity of GSK Biologicals' pneumococcal vaccine 1024850A when administered to children between 8 weeks and 2 years of age | http://clinicaltrials.gov/show/NCT01175083 | Partial exclusion for HBV, HiB, and Polio, Partial exclusion for RCT group | No Ab (HBV, HiB, Polio) Sickle cell babies RCT group | Diphtheria, Pertussis-w (PT), Sp (01, 04, 05, 06a, 06b, 07f, 09v, 14, 18c, 19a, 19f, 23f), Tetanus |
| 115648 | Consistency study of GSK Biologicals' measles-mumps-rubella (MMR) vaccine (209762) (Priorix) comparing immunogenicity and safety to Merck & Co., Inc.'s MMR vaccine (M-M-R II), in healthy children 12 to 15 months of age | http://clinicaltrials.gov/show/NCT01702428 | Partial exclusion for Mumps, Sp and VZV | No Ab method (VZV) No Ab method (Mumps), No primary (Sp) | HAV, Measles, Rubella |
| 105910 | Compare immunogenicity & reactogenicity of 2 formulations of GSK Biologicals' DTPa-HBV-IPV/Hib vaccine (new vs current) given in healthy infants. The DTPa-HBV-IPV vaccine (new formulation) will also be assessed in a 3rd group of subjects | http://clinicaltrials.gov/show/NCT00320463 | Partial exclusion for Diphtheria, Tetanus, HiB, and Polio | No Ab baseline (Diphtheria, Tetanus, HiB, polio) | HBV, Pertussis-a (FHA, PRN, PT) |
| 759346-007 | Demonstrate non-inferiority of GSK Biologicals' Tritanrix™-HepB/Hib-MenAC vs Tritanrix™-HepB/Hiberix™ with respect to anti-HBs immune response, when given to healthy infants at 6,10 & 14 weeks age, after a birth dose of hepatitis B vaccine | http://clinicaltrials.gov/show/NCT00317122 | Partial exclusion for Diphtheria and Tetanus | No Ab baseline (Diphtheria, Tetanus) | HiB, Nmen (A, C), Pertussis-w (PT) |
| 100480 | Study to show lot-to-lot consistency of Hib-MenAC mixed with Tritanrix™-HBV, its non-inferiority to Tritanrix™-HBV/Hiberix™ with or without Meningitec™, and MenA response in 2, 4, 6 month infants with hepatitis B birth dose | http://clinicaltrials.gov/show/NCT00317161 | Partial exclusion for Diphtheria, Tetanus, and HBV | No Ab baseline (Diphtheria, Tetanus, HBV) | HiB, Nmen (A, C), Pertussis-w (PT) |
| 100478 | Study to show non-inferiority of Tritanrix™-HepB/Hib-MenAC (+/- hepatitis B vaccine at birth) versus Tritanrix™-HepB/Hiberix™ without hepatitis B vacc. at birth for antibody response to all vaccine antigens given in healthy infants | http://clinicaltrials.gov/show/NCT00290303 | Partial exclusion for Diphtheria, Tetanus, HBV, and Polio | No Ab baseline (Diphtheria, Tetanus, HBV, Polio) | HiB, Nmen (A, C), Pertussis-w (PT) |
| 110215 | Immunogenicity & safety study in preterm & full-term infants of GSK Biologicals' Hib-MenC vaccine, Menitorix™ co-administered with Infanrix™ penta & Prevenar™ at 2, 4, 6 months & as a booster with Infanrix™ IPV & Prevenar™ at 16-18 months | http://clinicaltrials.gov/show/NCT00586612 | Partial exclusion for Diphtheria, Tetanus, Pertussis, HBV, Polio, and Sp Partial exclusion for RCT group | No Ab (Diphtheria, Tetanus, Pertussis, Polio, Sp) No primary (HBV) Pre-term babies RCT group | HiB, Nmen (C) |
| 112157 | Immunogenicity and safety study of GlaxoSmithKline Biologicals' GSK2202083A vaccine in healthy infants at 2, 3 and 4 months of age | http://clinicaltrials.gov/show/NCT00970307 | Partial exclusion for Diphtheria, Tetanus, HBV, | No Ab (rota), No Ab baseline (Diphtheria, | HiB, Nmen (C), Pertussis-a (FHA, PRN, PT) |

| Study ID | Study title | Link | Exclusion | Reason | Inclusion per pathogen |
|---|---|---|---|---|---|
| 217744-076 | An open, multicenter, phase IV clinical trial to assess the immunogenicity and reactogenicity of three doses of GSK Biologicals' combined DTPa-HBV-IPV/Hib vaccine in healthy infants at 2, 4 and 6 months of age, when co-administered with Wyeth-Lederle's meningococcal group C conjugate vaccine. | NA | Included | Included | Diphtheria, HBV, HiB, IPV (1, 2, 3), Nmen (C), Pertussis-a (FHA, PRN, PT), Tetanus |
| 711202-001 | Evaluate immunogenicity, reactogenicity, safety of GSK Biologicals' MenC-TT vaccine (2 formulations) given with Infanrix hexa® + GSK Biologicals' Hib MenC-TT vaccine (2 formulations) given with Infanrix penta® to infants in mths 3,4,5 of life | http://clinicaltrials.gov/show/NCT00135486 | Included | Included | Diphtheria, HBV, HiB, IPV (1, 2, 3), Nmen (C), Pertussis-a (FHA, PRN, PT), Tetanus |
| | | | Polio, Rotavirus, and Sp | Tetanus, HBV, Polio, Sp) | |
| 792014-003 | A phase II, open (partially double-blind), randomised, controlled, multicentre, primary vaccination study to evaluate the immunogenicity, reactogenicity and safety of three different formulations of GSK Biologicals' combined Haemophilus influenzae type b-meningococcal serogroups C and Y- conjugate vaccine and one formulation of GSK Biologicals' Haemophilus influenzae type b-meningococcal serogroup C conjugate vaccine each given concomitantly with InfanrixTM penta, versus MeningitecTM, given concomitantly with InfanrixTM hexa in infants according to a 2-3-4 month schedule | http://clinicaltrials.gov/show/NCT00129116 | Partial exclusion for Diphtheria, HBV, and Polio | No Ab baseline (Diphtheria, HBV, Polio) | HiB, Nmen (C), Pertussis-a (FHA, PRN, PT), Tetanus |
| 112679 | Immunogenicity and safety of GSK Biologicals' IPV (Poliorix™) in infants | http://clinicaltrials.gov/show/NCT01021293 | Partial exclusion for Diphtheria, Tetanus, Pertussis, HiB, and Polio | No Ab (Diphtheria, Tetanus, Pertussis, HiB),<br>No Ab baseline (Polio) | IPV1, IPV2, IPV3 |
| 111157 | Immunogenicity and safety study of GlaxoSmithKline Biologicals' Infanrix hexa™ vaccine in healthy infants in India | http://clinicaltrials.gov/show/NCT01353703 | Partial exclusion for Diphtheria, Tetanus, and HiB | No Ab baseline (Diphtheria, Tetanus, HiB) | IPV (1, 2, 3), Pertussis-a (FHA, PRN, PT) |
| 759348-001 | A randomised, controlled phase II study to evaluate the safety and immunogenicity of 7 different formulations of GSK Biologicals' investigational vaccination regimen, when administered as a 3-dose primary immunisation schedule before 6 months of age, followed by a fourth dose during the second year of life | NA | Partial exclusion for Diphtheria, Tetanus, HBV, HiB, and Nmen | No Ab baseline (Diphtheria, Tetanus, HBV, HiB, Nmen) | IPV (1, 2, 3), Pertussis-a (FHA, PRN, PT), Sp (01, 03, 04, 05, 06a, 06b, 07f, 09v, 14, 18c, 19a, 19f, 23f) |
| 347414-029 | Multinational, randomised, controlled, open, phase 2 clinical study to evaluate the safety and immunogenicity of GSK Biologicals' investigational vaccination regimen administered as a booster dose to healthy children, previously vaccinated in infancy with the investigational vaccination regimen in a primary study 347414/017 | NA | Partial exclusion for Diphtheria, Tetanus, Pertussis, HBV, HiB, and Sp | No primary (Diphtheria, Tetanus, Pertussis, HBV, HiB, Sp) | Measles, Mumps, Rubella |
| 115555 | Immunogenicity and safety study of GlaxoSmithKline Biological's live attenuated measles mumps rubella varicella vaccine (PriorixTetra™) when co-administered with conjugated Meningococcal C vaccine (Meningitec®, Nuron Biotechs' Vaccine) in healthy children | http://clinicaltrials.gov/show/NCT01506193 | Partial exclusion for Nmen and VZV | No Ab method (VZV),<br>No Ab baseline (Nmen) | Measles, Mumps, Rubella |
| 209762-147 | Phase IV open study to assess the safety, reactogenicity and immunogenicity of GlaxoSmithKline (GSK)Biologicals live attenuated Measles-Mumps-Rubella (MMR) vaccine when given to healthy children at the age of 12 to 18months in Singapore. | http://clinicaltrials.gov/show/NCT00388440 | Partial exclusion for VZV | No Ab (VZV) | Measles, Mumps, Rubella |
| 209762-151 | Phase II study to evaluate immunogenicity, reactogenicity and safety of GSK Bios' investigational vaccination regimen compared to the currently licensed GSK Bios' Priorix™ and Merck and Co.'s M-M-R®II vaccines when administered as a primary vaccination to healthy children aged 12-24 months | NA | Included | Included | Measles, Mumps, Rubella |
| 208136-013 | Study to assess immunogenicity and safety of one dose of GSK Bios' live attenuated MMRV vaccine, co-admind with a booster dose of the GSK Bios' combined diphtheria-tetanus-acellular pertussis-hepatitis B-inactivated polio-Haemophilus influenzae type b conjugate vaccine | NA | Partial exclusion for Diphtheria, Tetanus, Pertussis, HBV, HiB, and Polio | No primary routine (Diphtheria, Tetanus, Pertussis, HBV, HiB, Polio) | Measles, Mumps, Rubella, VZV |
| 110876 | Immunogenicity & safety study of GSK Biologicals' combined measles-mumps-rubella-varicella vaccine 208136 | http://clinicaltrials.gov/show/NCT00751348 | Included | Included | Measles, Mumps, Rubella, VZV |
| 109995 | A phase IIIb, open, randomised, multicentre, primary study in healthy children, to establish the non-inferiority of GlaxoSmithKline (GSK) Biologicals' MeMuRu-OKA vaccine (administered at 9 and 15 months of age) versus Priorix™ (9 months of age) and Priorix™ co-administered with Varilrix™ at 15 months of age (comparator) and also to evaluate the | http://clinicaltrials.gov/show/NCT00969436 | Included | Included | Measles, Mumps, Rubella, VZV |

| Study ID | Study title | Link | Exclusion | Reason | Inclusion per pathogen |
|---|---|---|---|---|---|
| 217744-076 | An open, multicenter, phase IV clinical trial to assess the immunogenicity and reactogenicity of three doses of GSK Biologicals' combined DTPa-HBV-IPV/Hib vaccine in healthy infants at 2, 4 and 6 months of age, when co-administered with Wyeth-Lederle's meningococcal group C conjugate vaccine. | NA | Included | Included | Diphtheria, HBV, HiB, IPV (1, 2, 3), Nmen (C), Pertussis-a (FHA, PRN, PT), Tetanus |
| 711202-001 | Evaluate immunogenicity, reactogenicity, safety of GSK Biologicals' MenC-TT vaccine (2 formulations) given with Infanrix hexa® + GSK Biologicals' Hib MenC-TT vaccine (2 formulations) given with Infanrix penta® to infants in mths 3,4,5 of life | http://clinicaltrials.gov/show/NCT00135486 | Included | Included | Diphtheria, HBV, HiB, IPV (1, 2, 3), Nmen (C), Pertussis-a (FHA, PRN, PT), Tetanus |
| | non-inferiority of Priorix™ (9 months of age) and MeMuRu-OKA vaccine (15 months of age) versus the comparator, all administered subcutaneously as two-dose primary vaccination course | | | | |
| 103388 | Blinded, randomized study to evaluate the immunogenicity and safety of GlaxoSmithKline Biologicals' measles-mumps-rubella-varicella candidate vaccine given to healthy children during the second year of life | http://clinicaltrials.gov/show/NCT00127010 | Included | Included | Measles, Mumps, Rubella, VZV |
| 104020 | Blinded, randomised study to assess the immunogenicity and safety of GlaxoSmithKline (GSK) Biologicals' live attenuated measles-mumps-rubella-varicella candidate vaccine when given to healthy children in their second year of life | http://clinicaltrials.gov/show/NCT00126997 | Included | Included | Measles, Mumps, Rubella, VZV |
| 104389 | Blinded, randomised, controlled study to evaluate the immunogenicity and safety of GlaxoSmithKline Biologicals' combined measles-mumps-rubella-varicella candidate vaccine given to healthy children in their second year of life | http://clinicaltrials.gov/show/NCT00127023 | Included | Included | Measles, Mumps, Rubella, VZV |
| 108760 | Immunogenicity & safety study of GSK Biologicals' 208136 vaccine formulated with new measles and rubella working seeds | http://clinicaltrials.gov/show/NCT00892775 | Included | Included | Measles, Mumps, Rubella, VZV |
| 208136-038 | Study to evaluate immunogenicity and safety of three production lots of GSK Biologicals' combined MeMuRu-OKA candidate vaccine given on a two-dose schedule to healthy children in their second year of life, as compared to separate administration of GSK Biologicals' Priorix™ and Varilrix™ vaccines | NA | Included | Included | Measles, Mumps, Rubella, VZV |
| 100388 | Study in Healthy Children (<2 Years) to Evaluate the Safety and Efficacy of GSK Biologicals' Live Attenuated Varicella Vaccine (VarilrixTM) and of GSK Biologicals' Combined Measles-Mumps-Rubella-Varicella Vaccine | http://clinicaltrials.gov/show/NCT00226499 | Included | Included | Measles, Mumps, Rubella, VZV |
| 209762-136 | A phase III, blinded, randomized, multicenter U.S. study evaluating the clinical consistency of three production lots of SmithKline Beecham Biologicals' MMR vaccine (PRIORIX) and comparability of PRIORIX with Merck's M-M-R IIvaccine, administered to healthy children 12 to 18 months of age | NA | Included | Included | Measles, Mumps, Rubella, VZV |
| 209762-148 | Phase II, double blind, randomized sequential study to compare immunogenicity and reactogenicity of the current formulations of GSK Bios' measles-mumps-rubella vaccine and GSK Bios' varicella vaccine containing human serum albumin with the modified formulations which do not contain HSA | NA | Included | Included | Measles, Mumps, Rubella, VZV |
| 208136-006 | Study evaluating in healthy children safety and immunogenicity of GSK Bios' combined Measles-Mumps-Rubella-Varicella vaccine, compared to GSK Bios' Varicella and Measles-Mumps-Rubella vaccines admind as separate injections | NA | Included | Included | Measles, Mumps, Rubella, VZV |
| 208136-018 | Study to evaluate immunogenicity and safety of GSK Bios' MeMuRu-OKA vaccine compared to concomitant administrations of GSK Bios' Priorix™ and Varilrix™ vaccines | NA | Included | Included | Measles, Mumps, Rubella, VZV |
| 208136-007 | Study to assess consistency of 3 production lots of GSK Bios' combined measles-mumps-rubella-varicella vaccine in terms of immunogenicity and safety, compared to administration of GSK Bios' measles-mumps-rubella vaccine and varicella vaccine in healthy children in their second year of life | NA | Included | Included | Measles, Mumps, Rubella, VZV |
| 208136-016 | Study to assess immunogenicity and safety of two lots of GSK Bios' live attenuated measles-mumps-rubella-varicella vaccine, at two different titres, given as a single injection to healthy children with GSK Bios' measles-mumps-rubella vaccine as control group | NA | Included | Included | Measles, Mumps, Rubella, VZV |
| 115649 | Immunogenicity and safety study of GSK Biologicals' Priorix vaccine (209762) at an end of shelf-life potency compared to Merck & Co., Inc.'s measles-mumps-rubella (MMR) vaccine when both are given on a 2-dose schedule to healthy children in their 2nd year of life | http://clinicaltrials.gov/show/NCT01681992 | Partial exclusion for HAV, Mumps, Sp, and VZV | No Ab HAV, Sp, VZV),<br>No Ab method (Mumps),<br>No primary (Sp) | Measles, rubel |
| 115650 | Safety and immunogenicity study of GSK Biologicals' measles-mumps-rubella (MMR) vaccine (209762) comparing immunogenicity and safety to Merck & Co., Inc.'s MMR vaccine, in healthy children 12 to 15 months of age | http://clinicaltrials.gov/show/NCT02184572 | Partial exclusion for HAV, Mumps, Sp, and VZV | No Ab (HAV, Sp VZV), | Measles, rubel |

| Study ID | Study title | Link | Exclusion | Reason | Inclusion per pathogen |
|---|---|---|---|---|---|
| 217744-076 | An open, multicenter, phase IV clinical trial to assess the immunogenicity and reactogenicity of three doses of GSK Biologicals' combined DTPa-HBV-IPV/Hib vaccine in healthy infants at 2, 4 and 6 months of age, when co-administered with Wyeth-Lederle's meningococcal group C conjugate vaccine. | NA | Included | Included | Diphtheria, HBV, HiB, IPV (1, 2, 3), Nmen (C), Pertussis-a (FHA, PRN, PT), Tetanus |
| 711202-001 | Evaluate immunogenicity, reactogenicity, safety of GSK Biologicals' MenC-TT vaccine (2 formulations) given with Infanrix hexa® + GSK Biologicals' Hib MenC-TT vaccine (2 formulations) given with Infanrix penta® to infants in mths 3,4,5 of life | http://clinicaltrials.gov/show/NCT00135486 | Included | Included | Diphtheria, HBV, HiB, IPV (1, 2, 3), Nmen (C), Pertussis-a (FHA, PRN, PT), Tetanus |
| | | | | No Ab method (Mumps), No primary (Sp) | |
| 217744-097 | Study to assess immunogenicity and reactogenicity of three doses of GSK Bio's combined Hib-MenC vaccine co-admind with GSK Bio's DTPa-HBV-IPV vaccine and of two doses of Baxters meningococcal C conjugate vaccine co-admind with GSK Bio's DTPa-HBV-IPV/Hib vaccine | http://clinicaltrials.gov/show/NCT00352963 | Partial exclusion for Diphtheria, Tetanus, HBV, HiB, and Polio | No Ab baseline (Diphtheria, Tetanus, HBV, HiB, Polio) | Nmen (C), Pertussis-a (FHA, PRN, PT) |
| 103488 | A randomized, controlled, phase II study to evaluate the safety and immunogenicity of different formulations of GlaxoSmithKline Biologicals' 11-valent pneumococcal conjugate vaccine, when administered intramuscularly as a 3-dose primary immunization (2-3-4 month schedule) before 6 months of age | http://clinicaltrials.gov/show/NCT00169481 | Partial exclusion for Diphtheria, Tetanus, HBV, HiB, and Polio | No Ab baseline (Diphtheria, Tetanus, HBV, HiB, Polio) | Pertussis-a (FHA, PRN, PT), Sp (01, 03, 04, 05, 06a, 06b, 07f, 09v, 14, 18c, 19a, 19f, 23f) |
| 759348-007 | A randomized, controlled, phase II study to evaluate the safety and immunogenicity of five formulations of GlaxoSmithKline Biologicals' investigational vaccination regimen, when administered intramuscularly as a 3-dose primary immunization schedule (2-3-4 month schedule) before 6 months of age. | NA | Partial exclusion for Diphtheria, Tetanus, HBV, HiB, and Polio | No Ab baseline (Diphtheria, Tetanus, HBV, HiB, Polio) | Pertussis-a (FHA, PRN, PT), Sp (01, 03, 04, 05, 06b, 07f, 09v, 14, 18c, 19f, 23f) |
| 104489 | Study to assess immunogenicity and safety of GlaxoSmithKline Biologicals' Kft's DTPw-HBV/Hib vs DTPwCSL-HBV/Hib Kft and vs concomitant administration of CSL's Triple Antigen and GlaxoSmithKline Biologicals' Hiberix, to infants at 2, 4, 6 months of age, after a birth dose of hepatitis B | http://clinicaltrials.gov/show/NCT00316680 | Partial exclusion for Diphtheria, Tetanus, HBV, and HiB | No Ab baseline (Diphtheria, Tetanus, HBV, HiB) | Pertussis-w (PT) |
| 101223 | Study to assess the lot-to-lot consistency of the production method of GSK Biologicals' DTPw-HBV/ Hib Kft. vaccine and to compare to GSK Biologicals' Tritanrix™-HepB/Hiberix™ vaccine, when administered as a primary vaccination course. | NA | Partial exclusion for Diphtheria, Tetanus, HBV, HiB, and Polio | No Ab (Polio), No Ab baseline (Diphtheria, Tetanus, HBV, HiB) | Pertussis-w (PT) |
| 113808 | Efficacy, immunogenicity and safety of two doses of GlaxoSmithKline (GSK) Biologicals' Oral Live Attenuated Liquid Human Rotavirus (HRV) Vaccine (444563), in healthy infants | http://clinicaltrials.gov/show/NCT01171963 | Partial exclusion for Diphtheria, Tetanus, and Pertussis | No Ab method (Diphtheria, Tetanus, Pertussis) | Rotavirus |
| 444563-021 | Phase II, double-blind, randomized, placebo-controlled clinical study to assess immunogenicity and reactogenicity of doses of a modified vaccine formulation versus GSK Biologicals' live attenuated human rotavirus vaccine when orally administered to healthy infants at 2, 4 and 6 months of age | NA | Partial exclusion for Diphtheria, Tetanus, Pertussis, HBV, HiB | No Ab (Diphtheria, Tetanus, Pertussis, HBV, HiB) | Rotavirus |
| 115461 | Immunogenicity and safety study of two formulations of GlaxoSmithKline (GSK) Biologicals' human rotavirus (HRV) vaccine (444563), in healthy infants starting at age 6-12 weeks | http://clinicaltrials.gov/show/NCT02914184 | Partial exclusion for Diphtheria, Tetanus, Pertussis, HBV, HiB, Nmen, Polio, and Sp | No Ab (Diphtheria, Tetanus, Pertussis, HBV, HiB, Nmen, Polio, Sp) | Rotavirus |
| 102247 | A multi-country & multi-center study to assess the efficacy, safety & immunogenicity of 2 doses of GSK Biologicals' oral live attenuated human rotavirus (HRV) vaccine in healthy infants in co-administration with specific childhood vaccines | http://clinicaltrials.gov/show/NCT00140686 | Partial exclusion for Diphtheria, Tetanus, Pertussis, HBV, HiB, Nmen, Polio, and Sp | No Ab baseline (Diphtheria, Tetanus, Pertussis, HBV, HiB, Nmen, Polio, Sp) | Rotavirus |

| Study ID | Study title | Link | Exclusion | Reason | Inclusion per pathogen |
|---|---|---|---|---|---|
| 217744-076 | An open, multicenter, phase IV clinical trial to assess the immunogenicity and reactogenicity of three doses of GSK Biologicals' combined DTPa-HBV-IPV/Hib vaccine in healthy infants at 2, 4 and 6 months of age, when co-administered with Wyeth-Lederle's meningococcal group C conjugate vaccine. | NA | Included | Included | Diphtheria, HBV, HiB, IPV (1, 2, 3), Nmen (C), Pertussis-a (FHA, PRN, PT), Tetanus |
| 711202-001 | Evaluate immunogenicity, reactogenicity, safety of GSK Biologicals' MenC-TT vaccine (2 formulations) given with Infanrix hexa® + GSK Biologicals' Hib MenC-TT vaccine (2 formulations) given with Infanrix penta® to infants in mths 3,4,5 of life | http://clinicaltrials.gov/show/NCT00135486 | Included | Included | Diphtheria, HBV, HiB, IPV (1, 2, 3), Nmen (C), Pertussis-a (FHA, PRN, PT), Tetanus |
| 106260 | A Phase IIIb, Randomized, Double-Blind, Placebo-Controlled Study to Explore the Existence of Horizontal Transmission of the RIX4414 Vaccine Strain Between Twins Within a Family. | http://clinicaltrials.gov/show/NCT00396630 | Partial exclusion for Diphtheria, Tetanus, Pertussis, HBV, HiB, and Polio | No Ab (Diphtheria, Tetanus, Pertussis, HBV, HiB, Polio) | Rotavirus |
| 107077 | A study to assess the immunogenicity, reactogenicity and safety of 2 different formulations of GSK Biologicals' live attenuated HRV vaccine, given as a two-dose primary vaccination, in healthy infants previously uninfected with HRV | http://clinicaltrials.gov/show/NCT00363545 | Partial exclusion for Diphtheria, Tetanus, Pertussis, HBV, HiB, and Polio | No Ab (Diphtheria, Tetanus, Pertussis, HBV, HiB, Polio) | Rotavirus |
| 107876 | Study to evaluate clinical consistency of the liquid formulation of GSK Biologicals' HRV vaccine and to evaluate liquid formulation compared to lyophilised formulation of the HRV vaccine administered as a two-dose primary vaccination. | http://clinicaltrials.gov/show/NCT00382772 | Partial exclusion for Diphtheria, Tetanus, Pertussis, HBV, HiB, and Polio | No Ab (Diphtheria, Tetanus, Pertussis, HBV, HiB, Polio) | Rotavirus |
| 444563-020 | Phase II, double-blind randomised, placebo controlled clinical dose-range study to assess immunogenicity and reactogenicity of an investigational vaccination regimen, and to assess immunogenicity of OPV orally co-administered to healthy infants at 2, 4 and 6 months of age | NA | Partial exclusion for Diphtheria, Tetanus, Pertussis, HBV, HiB, and Polio | No Ab baseline (Diphtheria, Tetanus, Pertussis, HBV, HiB, Polio) | Rotavirus |
| 444563-005 | Phase II, double-blind, randomized, placebo-controlled study of 2 doses of GSK Bios' live attenuated human rotavirus vaccine at different virus concentrations (10 5.2 and 10 6.4 ffu) in healthy infants following a 0, 2 month schedule and previously uninfected with human rotavirus | http://clinicaltrials.gov/show/NCT00729001 | Partial exclusion for Diphtheria, Tetanus, Pertussis, HBV, Nmen, Polio, and Sp | No Ab (HBV, Nmen), No Ab baseline (Diphtheria, Tetanus, Pertussis, Polio, Sp) | Rotavirus |
| 103992 | Evaluate immunogenicity, reactogenicity & safety of 2 doses of GSK Biologicals' oral live attenuated HRV vaccine (RIX4414 at 106.5 CCID50) when given concomitantly with OPV versus given alone (HRV vaccine dose given 15 days after the OPV dose) in healthy infants in Bangladesh | http://clinicaltrials.gov/show/NCT00139334 | Partial exclusion for Diphtheria, Tetanus, Pertussis, HBV, and Polio | No Ab (Diphtheria, Tetanus, Pertussis, HBV), No primary (Polio) | Rotavirus |
| 105722 | A placebo-controlled study to evaluate the immunogenicity, reactogenicity and safety of two doses of GSK Biologicals' oral live attenuated human rotavirus (HRV) liquid vaccine, when given to healthy infants, in Vietnam | http://clinicaltrials.gov/show/NCT00345956 | Partial exclusion for Diphtheria, Tetanus, Pertussis, HBV, and Polio | No Ab (Diphtheria, Tetanus, Pertussis, HBV, Polio) | Rotavirus |
| 444563-014 | Study of safety, reactogenicity and immunogenicity of two doses of GSK Bio's oral live attenuated human rotavirus vaccine co-administered with either oral polio vaccine (OPV) or inactivated polio vaccine (IPV) in healthy infants in South Africa | http://clinicaltrials.gov/show/NCT00346892 | Partial exclusion for Diphtheria, Tetanus, Pertussis, HBV, and Polio | No Ab (Diphtheria, Tetanus, Pertussis, HiB), No primary (Polio) | Rotavirus |
| 444563-007 | Study to assess efficacy, immunogenicity, reactogenicity and safety of two doses of GSK Bio's oral live attenuated human rotavirus vaccine at different viral concentrations in healthy infants previously uninfected with human rotavirus and approximately 3 months of age | http://clinicaltrials.gov/show/NCT00429481 | Partial exclusion for Diphtheria, Tetanus, Pertussis, HBV, and Polio | No Ab baseline (Diphtheria, Tetanus, Pertussis, HBV, HiB, Polio) | Rotavirus |

| Study ID | Study title | Link | Exclusion | Reason | Inclusion per pathogen |
|---|---|---|---|---|---|
| 217744-076 | An open, multicenter, phase IV clinical trial to assess the immunogenicity and reactogenicity of three doses of GSK Biologicals' combined DTPa-HBV-IPV/Hib vaccine in healthy infants at 2, 4 and 6 months of age, when co-administered with Wyeth-Lederle's meningococcal group C conjugate vaccine. | NA | Included | Included | Diphtheria, HBV, HiB, IPV (1, 2, 3), Nmen (C), Pertussis-a (FHA, PRN, PT), Tetanus |
| 711202-001 | Evaluate immunogenicity, reactogenicity, safety of GSK Biologicals' MenC-TT vaccine (2 formulations) given with Infanrix hexa® + GSK Biologicals' Hib MenC-TT vaccine (2 formulations) given with Infanrix penta® to infants in mths 3,4,5 of life | http://clinicaltrials.gov/show/NCT00135486 | Included | Included | Diphtheria, HBV, HiB, IPV (1, 2, 3), Nmen (C), Pertussis-a (FHA, PRN, PT), Tetanus |
| 444563-013 | A phase II, randomized, double-blind, placebo-controlled study of safety, reactogenicity and immunogenicity of 2 or 3 doses of GSK Biologicals' oral live attenuated human rotavirus vaccine at 10E6.5 CCID50 viral concentration in healthy infants (approximately 5-10 weeks old) in the Republic of South Africa | http://clinicaltrials.gov/show/NCT00383903 | Partial exclusion for Polio | No primary routine (Polio) | Rotavirus |
| 444563-003 | A phase II, double-blind, randomized, placebo-controlled, dose-escalating, stepwise study to assess safety, reactogenicity and immunogenicity of GlaxoSmithKline Biologicals' live attenuated human rotavirus (HRV) vaccine in healthy infants previously uninfected with human rotavirus. | NA | Partial exclusion for RCT group | Placebo RCT group | Rotavirus |
| 109216 | Immunogenicity and safety of two doses of GlaxoSmithKline (GSK) Biologicals' oral live attenuated human rotavirus (HRV) liquid vaccine (GSK 357941A) in healthy infants. | http://clinicaltrials.gov/show/NCT00432380 | Included | Included | Rotavirus |
| 112269 | Immunogenicity, reactogenicity and safety study to evaluate two doses of the lyophilised formulation of the human rotavirus (HRV) vaccine when administered to healthy Korean infants previously uninfected with HRV | http://clinicaltrials.gov/show/NCT00969228 | Included | Included | Rotavirus |
| 113518 | Reactogenicity and safety of two doses of GlaxoSmithKline (GSK) Biologicals' oral live attenuated liquid human rotavirus (HRV) vaccine 444563, in healthy infants | http://clinicaltrials.gov/show/NCT01107587 | Included | Included | Rotavirus |
| 101555 | A phase II, double-blind, randomized, placebo-controlled study to compare the immunogenicity, reactogenicity and safety of 2 different formulations of GSK Biologicals' live attenuated human rotavirus (HRV) vaccine given as a two-dose primary vaccination in healthy infants previously uninfected with HRV | NA | Included | Included | Rotavirus |
| 444563-033 | Study to assess the clinical consistency of three production lots of GSK Biologicals' HRV vaccine in terms of immunogenicity and safety when given to healthy infants at 2 and 4 months of age | http://clinicaltrials.gov/show/NCT00757770 | Included | Included | Rotavirus |
| 102248 | Multi-Center Study to Assess the Efficacy, Safety and Immunogenicity of 2 or 3 Doses of GSK Biologicals' Oral Live Attenuated Human Rotavirus (HRV) Vaccine Given Concomitantly With Routine EPI Vaccinations in Healthy Infants | http://clinicaltrials.gov/show/NCT00241644 | Included | Included | Rotavirus |
| 103477 | Study to assess the immunogenicity, reactogenicity and safety of GlaxoSmithKline Biologicals' oral live attenuated human rotavirus (HRV) vaccine following a 0, 2 month schedule, in healthy infants previously uninfected with human rotavirus | http://clinicaltrials.gov/show/NCT00169455 | Included | Included | Rotavirus |
| 103478 | Assess the immunogenicity, safety & reactogenicity of 2 doses of GSK Biologicals' oral live attenuated human rotavirus (HRV) vaccine in healthy infants (6-12 weeks of age at first dose) previously uninfected with human rotavirus | http://clinicaltrials.gov/show/NCT00134732 | Included | Included | Rotavirus |
| 104480 | Compare the immunogenicity, reactogenicity & safety of 2 different formulations of GSK Biologicals' live attenuated human rotavirus (HRV) vaccine given as a two-dose primary vaccination in healthy infants previously uninfected with HRV | http://clinicaltrials.gov/show/NCT00137930 | Included | Included | Rotavirus |
| 444563-004 | A study to assess the efficacy, immunogenicity and safety of two doses of oral live attenuated human rotavirus (HRV) vaccine (Rotarix) in healthy infants. | http://clinicaltrials.gov/show/NCT00425737 | Included | Included | Rotavirus |
| 114351 | Immunogenicity, reactogenicity and safety study of GlaxoSmithKline (GSK) Biologicals' oral live attenuated human rotavirus (HRV) vaccine in healthy Taiwanese infants who received hepatitis B immunoglobulin after birth. | http://clinicaltrials.gov/show/NCT01198769 | Included | Included | Rotavirus |
| 111634 | Primary and booster vaccination course in human immunodeficiency virus (HIV) infected infants, HIV exposed uninfected infants and unexposed uninfected infants receiving the pneumococcal vaccine GSK 1024850A. | http://clinicaltrials.gov/show/NCT00829010 | Partial exclusion for Diphtheria, Tetanus, Pertussis, HBV, HiB, Measles, and Polio, | No vaccine concentration (Measles),<br>No Ab baseline (Diphtheria, Tetanus, Pertussis, HBV, HiB, Polio), | Rotavirus, Sp (01, 04, 05, 06a, 06b, 07f, 09v, 14, 18c, 19a, 19f, 23f) |

| Study ID | Study title | Link | Exclusion | Reason | Inclusion per pathogen |
|---|---|---|---|---|---|
| 217744-076 | An open, multicenter, phase IV clinical trial to assess the immunogenicity and reactogenicity of three doses of GSK Biologicals' combined DTPa-HBV-IPV/Hib vaccine in healthy infants at 2, 4 and 6 months of age, when co-administered with Wyeth-Lederle's meningococcal group C conjugate vaccine. | NA | Included | Included | Diphtheria, HBV, HiB, IPV (1, 2, 3), Nmen (C), Pertussis-a (FHA, PRN, PT), Tetanus |
| 711202-001 | Evaluate immunogenicity, reactogenicity, safety of GSK Biologicals' MenC-TT vaccine (2 formulations) given with Infanrix hexa® + GSK Biologicals' Hib MenC-TT vaccine (2 formulations) given with Infanrix penta® to infants in mths 3,4,5 of life | http://clinicaltrials.gov/show/NCT00135486 | Included | Included | Diphtheria, HBV, HiB, IPV (1, 2, 3), Nmen (C), Pertussis-a (FHA, PRN, PT), Tetanus |
| | | | Partial exclusion for RCT group | HIV+ babies RCT group | |
| 110521 | Primary vaccination course in children receiving the pneumococcal vaccine GSK 1024850A co-administered with Zilbrix™ Hib and Polio Sabin™ | http://clinicaltrials.gov/show/NCT00678301 | Partial exclusion for Diphtheria, Tetanus, Pertussis, HBV, HiB, and Polio | No Ab (Polio), No primary (HBV), No Ab baseline (Diphtheria, Tetanus, Pertussis, HiB) | Sp (01, 04, 05, 06a, 06b, 07f, 09v, 14, 18c, 19a, 19f, 23f) |
| 109861 | Primary vaccination course in children receiving the pneumococcal vaccine GSK 1024850A, Infanrix hexa and Rotarix | http://clinicaltrials.gov/show/NCT00533507 | Partial exclusion for Diphtheria, Tetanus, Pertussis, HBV, HiB, Polio, and Rotavirus | No Ab baseline (Diphtheria, Tetanus, Pertussis, HiB, Polio, Rotavirus), No primary (HBV) | Sp (01, 04, 05, 06a, 06b, 07f, 09v, 14, 18c, 19a, 19f, 23f) |
| 105908 | Comparative study evaluating the immunogenicity and safety of MeMuRu-OKA vaccine and measles-mumps-rubella vaccine (Priorix™) co-administered with varicella vaccine (Varilrix™) in children primed with measles-mumps-rubella vaccine | http://clinicaltrials.gov/show/NCT00353288 | Partial exclusion for Measles, Mumps, and Rubella | No primary (Measles, Mumps, Rubella) | VZV |
| 109705 | Study of two formulations of GSK Biologicals' varicella vaccine given as a 2-dose course in the second year of life | http://clinicaltrials.gov/show/NCT00568334 | Included | Included | VZV |
| 104727 | Assess immunogenicity, safety & reactogenicity of a 4th dose of GSK Biologicals' Tritanrix™-HepB/Hib-MenAC at 15-24 m & of a dose of Mencevax™ ACWY at 24-30 m in subjects primed with 3 doses of Tritanrix™-HepB/Hib-MenAC | http://clinicaltrials.gov/show/NCT00136604 | Excluded for all | Added to 100480 | NA |
| 102015 | Evaluate Immuno and Safety of GSKBiologicals' HibMenCYTT vs Licensed Hib Conjugate Vaccine, Each Coadministered With Pediarix® and Prevnar®, in Healthy Infants. An Exploratory Control Group Will Receive Licensed Menomune® at 3 to 5 years | http://clinicaltrials.gov/show/NCT00129129 | Excluded for all | Added to 101858 | NA |
| 107824 | A study to evaluate the long-term antibody persistence at 1, 3 & 5 years after the administration of a fourth dose of Hib-MenCY-TT Vaccine compared to ActHIB in subjects boosted in a previous study. | http://clinicaltrials.gov/show/NCT00359983 | Excluded for all | Added to 101858 | NA |
| 104056 | Study to demonstrate the non-inferiority of the meningococcal serogroup C immune response of GlaxoSmithKline Biologicals' Hib-MenC vaccine co-administered with Infanrix™-IPV versus a licensed meningococcal serogroup C vaccine co-administered with Pediacel™ vaccine | NA | Excluded for all | Added to 103974 | NA |
| 109664 | Assessment of long-term antibody persistence after a booster dose of GSK Biologicals' Hib & meningococcal C vaccine (Menitorix™) 811936 given at 12-15 months of age to subjects primed with 3 doses of Menitorix™ at 2, 3, 4 months of age | http://clinicaltrials.gov/show/NCT00454987 | Excluded for all | Added to 103974 | NA |
| 111736 | Vaccination course in children primed and boosted with pneumococcal vaccine GSK 1024850A and in age-matched unprimed children | http://clinicaltrials.gov/show/NCT00792909 | Excluded for all | Added to 105539 | NA |
| 106623 | Multicentre immune memory study in healthy children following a 3 dose primary vaccination with Prevenar or GSK Biologicals' pneumococcal conjugate vaccine via the administration of a single booster dose of Pneumovax 23 | http://clinicaltrials.gov/show/NCT00333450 | Excluded for all | Added to 105554 | NA |
| 112807 | Vaccination with the pneumococcal vaccine GSK 1024850A or Prevenar™ at approximately 4 years of age in children primed with 3 doses of GSK 1024850A vaccine or Prevenar™ and boosted with 23-valent pneumococcal plain polysaccharide vaccine | http://clinicaltrials.gov/show/NCT00907777 | Excluded for all | Added to 105554 | NA |

| Study ID | Study title | Link | Exclusion | Reason | Inclusion per pathogen |
|---|---|---|---|---|---|
| 217744-076 | An open, multicenter, phase IV clinical trial to assess the immunogenicity and reactogenicity of three doses of GSK Biologicals' combined DTPa-HBV-IPV/Hib vaccine in healthy infants at 2, 4 and 6 months of age, when co-administered with Wyeth-Lederle's meningococcal group C conjugate vaccine. | NA | Included | Included | Diphtheria, HBV, HiB, IPV (1, 2, 3), Nmen (C), Pertussis-a (FHA, PRN, PT), Tetanus |
| 711202-001 | Evaluate immunogenicity, reactogenicity, safety of GSK Biologicals' MenC-TT vaccine (2 formulations) given with Infanrix hexa® + GSK Biologicals' Hib MenC-TT vaccine (2 formulations) given with Infanrix penta® to infants in mths 3,4,5 of life | http://clinicaltrials.gov/show/NCT00135486 | Included | Included | Diphtheria, HBV, HiB, IPV (1, 2, 3), Nmen (C), Pertussis-a (FHA, PRN, PT), Tetanus |
| 109507 | Booster Vaccination With Pneumococcal Vaccine GSK1024850A, a DTPa-Combined and MenC or Hib-MenC Vaccines | http://clinicaltrials.gov/show/NCT00463437 | Excluded for all | Added to 107005 | NA |
| 112830 | Persistence of antibodies after full vaccination course with GSK Biologicals' Menitorix or MenC conjugate vaccine, co-administered with DTPa or DTPa/Hib containing vaccine and pneumococcal conjugate vaccine, in children up to 6 years of age | http://clinicaltrials.gov/show/NCT00891176 | Excluded for all | Added to 107005 | NA |
| 110217 | Immunogenicity & safety study in preterm & full-term infants of GSK Biologicals' Hib-MenC vaccine, Menitorix™ co-administered with Infanrix™ penta & Prevenar™ at 2, 4, 6 months & as a booster with Infanrix™ IPV & Prevenar™ at 16-18 months | http://clinicaltrials.gov/show/NCT00586612 | Excluded for all | Added to 110215 | NA |
| 114306 | Immunogenicity and safety of a booster dose of GlaxoSmithKline Biologicals' IPV (PoliorixTM) in healthy Chinese toddlers | http://clinicaltrials.gov/show/NCT01323647 | Excluded for all | Added to 112679 | NA |
| 208108-092 | A phase II, double-blind, randomized study to compare the immunogenicity, safety and reactogenicity of GlaxoSmithKline (GSK) Biologicals' Tritanrix™-HepB/Hib2.5 to GSK Biologicals' Tritanrix™-HepB/Hiberix™ when administered as a three-dose primary vaccination course to healthy infants at 6, 10 and 14 weeks of age. A dose of unconjugated Hib vaccine (plain PRP booster) will be administered at the age of 10 months to 50% of the subjects | http://clinicaltrials.gov/show/NCT01061541 | Excluded for all | Added to 208108-091 | NA |
| 106745 | Multicentre study to assess persistence of antibodies against hepatitis B & immune response to a hepatitis B challenge dose in healthy children 4 to 6 yrs old previously vaccinated with 4 doses of GSK Biologicals' DTPa-HBV-IPV/Hib vaccine | http://clinicaltrials.gov/show/NCT00335881 | Excluded for all | Added to 217744-078 | NA |
| 217744-081 | An open, multicentre, phase IV booster vaccination study to assess the immunogenicity and reactogenicity of a 4th dose of GSK Biologicals' combined DTPa-HBV-IPV/Hib vaccine, co-administered with Wyeth's seven-valent Pneumococcal conjugate vaccine at a different injection site during the same visit in healthy children | NA | Excluded for all | Added to 217744-078 (partially - no dates) | NA |
| 102547 | Evaluate immunogenicity,safety & reactogenicity of a booster dose of Hib-MenC conjugate vaccine when given to healthy subjects aged 13-14 months who were primed with 3 doses of Hib-MenC vs a booster dose of Infanrix hexa given to subjects primed with 3 doses of Infanrix hexa and Meningitec | http://clinicaltrials.gov/show/NCT00323050 | Excluded for all | Added to 217744-097 | NA |
| 106672 | Phase III, open, multicenter Study to Assess the Long-Term Persistence of a Booster Dose of GSK Biologicals' Hib-MenC compared to a Booster Dose of Infanrix™ Hexa When Given to 14 month-old Subjects Primed in study DTPa-HBV-IPV-097 & Boosted in study Hib-MenC-TT-010 BST: DTPa-HBV-IPV-097 | http://clinicaltrials.gov/show/NCT00322335 | Excluded for all | Added to 217744-097 | NA |
| 106673 | Phase III, open, multicenter Study to Assess the Long-Term Persistence of a Booster Dose of GSK Biologicals' Hib-MenC compared to a Booster Dose of Infanrix™ Hexa When Given to 14 month-old Subjects Primed in study DTPa-HBV-IPV-097 & Boosted in study Hib-MenC-TT-010 BST: DTPa-HBV-IPV-097 | http://clinicaltrials.gov/show/NCT00322335 | Excluded for all | Added to 217744-097 | NA |
| 106675 | Phase III, open, multicenter Study to Assess the Long-Term Persistence of a Booster Dose of GSK Biologicals' Hib-MenC compared to a Booster Dose of Infanrix™ Hexa When Given to 14 month-old Subjects Primed in study DTPa-HBV-IPV-097 & Boosted in study Hib-MenC-TT-010 BST: DTPa-HBV-IPV-097 | http://clinicaltrials.gov/show/NCT00322335 | Excluded for all | Added to 217744-097 | NA |
| 106679 | Phase III, open, multicenter Study to Assess the Long-Term Persistence of a Booster Dose of GSK Biologicals' Hib-MenC compared to a Booster Dose of Infanrix™ Hexa When Given to 14 month-old Subjects Primed in study DTPa-HBV-IPV-097 & Boosted in study Hib-MenC-TT-010 BST: DTPa-HBV-IPV-097 | http://clinicaltrials.gov/show/NCT00322335 | Excluded for all | Added to 217744-097 | NA |
| 106680 | Phase III, open, multicenter Study to Assess the Long-Term Persistence of a Booster Dose of GSK Biologicals' Hib-MenC compared to a Booster Dose of Infanrix™ Hexa When Given to 14 month-old Subjects Primed in study DTPa-HBV-IPV-097 & Boosted in study Hib-MenC-TT-010 BST: DTPa-HBV-IPV-097 | http://clinicaltrials.gov/show/NCT00322335 | Excluded for all | Added to 217744-097 | NA |
| 347414-026 | A Phase III, randomized single-blind clinical trial to assess the immune memory induced by an investigational vaccination regimen in comparison with Prevnar and the immune response to a fourth dose of the pneumococcal vaccine and the investigational vaccination regimen, when admininistered to healthy children (12-15 Months) | NA | Excluded for all | Added to 347414-020 (partially - no dates) | NA |

| Study ID | Study title | Link | Exclusion | Reason | Inclusion per pathogen |
|---|---|---|---|---|---|
| 217744-076 | An open, multicenter, phase IV clinical trial to assess the immunogenicity and reactogenicity of three doses of GSK Biologicals' combined DTPa-HBV-IPV/Hib vaccine in healthy infants at 2, 4 and 6 months of age, when co-administered with Wyeth-Lederle's meningococcal group C conjugate vaccine. | NA | Included | Included | Diphtheria, HBV, HiB, IPV (1, 2, 3), Nmen (C), Pertussis-a (FHA, PRN, PT), Tetanus |
| 711202-001 | Evaluate immunogenicity, reactogenicity, safety of GSK Biologicals' MenC-TT vaccine (2 formulations) given with Infanrix hexa® + GSK Biologicals' Hib MenC-TT vaccine (2 formulations) given with Infanrix penta® to infants in mths 3,4,5 of life | http://clinicaltrials.gov/show/NCT00135486 | Included | Included | Diphtheria, HBV, HiB, IPV (1, 2, 3), Nmen (C), Pertussis-a (FHA, PRN, PT), Tetanus |
| 347414-036 | A phase-3, open, controlled study to assess the safety and immunogenicity of four different formulations of GSK Biologicals' investigational vaccination regimen when administered as a booster to healthy infants, 12 to 16 months old, previously vaccinated in infancy in a primary study 347414/023 | NA | Excluded for all | Added to 347414-023 (partially - no dates) | NA |
| 759346-002 | Assess immune persistence & memory by giving plain PRP,PSA & PSC (10 mths age), & immunogenicity & safety of a Tritanrix™-HBV/Hib-MenAC/ Tritanrix™-HBV/Hib2.5 booster (15-18 mths age) in previously primed subjects | http://clinicaltrials.gov/show/NCT00317174 | Excluded for all | Added to 759346-001 | NA |
| 792014-002 | A phase II, open, randomized, controlled, multicentre, primary vaccination study to evaluate the immunogenicity, reactogenicity and safety of an investigational vaccination regimen versus ActHIB® and Menjugate® given concomitantly with Infanrix® penta and Prevenar® in infants | NA | Excluded for all | Added to 792014-001 | NA |
| 100381 | Study to assess safety, reactogenicity and immunogenicity of a booster dose of an investigational vaccination regimen and GSK Biologicals Hib-MenC vaccine (co-admind with Infanrix penta) compared to a booster dose of Menjugate (co-admind with Infanrix hexa) | NA | Excluded for all | Added to 792014-003 | NA |
| 107706 | To assess safety, reactogenicity & immunogenicity of a booster dose of pneumococcal conjugate vaccine, co-admin with GSK Biologicals' MMRV vaccine in children (2nd yr of life) primed with the pneumococcal conjugate vaccine in study 105553. | http://clinicaltrials.gov/show/NCT00370227 | Excluded for all | Booster 105553 | NA |
| 107046 | To assess the safety, reactogenicity & immunogenicity of a 4th dose of GSK Biologicals' pneumococcal vaccine or Prevenar™ in children (12-18 months) previously vaccinated in the primary study 105553 with either pneumococcal vaccine or Prevenar™ | http://clinicaltrials.gov/show/NCT00370396 | Excluded for all | Booster 105553 | NA |
| 109509 | Booster vaccination course with the pneumococcal vaccine GSK 1024850A, DTPw-HBV/Hib and OPV or IPV in children who completed the primary vaccination course in study 107007 | http://clinicaltrials.gov/show/NCT00547248 | Excluded for all | Booster 107007 | NA |
| 113166 | Safety, reactogenicity and immunogenicity study of GSK Biologicals' pneumococcal vaccine GSK1024850A, given either as a booster dose or as a 2-dose catch-up immunization in healthy Malian children | http://clinicaltrials.gov/show/NCT00985465 | Excluded for all | Booster 110521 | NA |
| 113199 | Safety, reactogenicity and immunogenicity study of GSK Biologicals' pneumococcal vaccine GSK1024850A, given either as a booster dose or as a 2-dose catch-up immunization in healthy Nigerian children | http://clinicaltrials.gov/show/NCT01153893 | Excluded for all | Booster 110521 | NA |
| 105555 | A phase II, multicentre booster study to evaluate booster vaccination with GSK Biologicals' 10-valent pneumococcal conjugate vaccine or to evaluate the immune memory following the administration of a single dose of 23-valent plain polysaccharide vaccine in healthy children, previously vaccinated in infancy in the primary study 11PN-PD-DIT-002 (103488) | http://clinicaltrials.gov/show/NCT00307567 | Excluded for all | Contained in 103488 | NA |
| 209762-150 | Double blind, randomized sequential study to compare immunogenicity and reactogenicity of current formulations of GSK Bios' measles-mumps-rubella vaccine and GSK Bios' varicella vaccine to investigational vaccination regimens | NA | Excluded for all | Contained in study 209762-148 | NA |
| 104105 | Study in Healthy Children (<2 Years) to Evaluate the Safety and Efficacy of GSK Biologicals' Live Attenuated Varicella Vaccine (VarilrixTM) and of GSK Biologicals' Combined Measles-Mumps-Rubella-Varicella Vaccine | http://clinicaltrials.gov/show/NCT00226499 | Excluded for all | Follow up 100388 | NA |
| 104106 | Study in Healthy Children (<2 Years) to Evaluate the Safety and Efficacy of GSK Biologicals' Live Attenuated Varicella Vaccine (VarilrixTM) and of GSK Biologicals' Combined Measles-Mumps-Rubella-Varicella Vaccine | http://clinicaltrials.gov/show/NCT00226499 | Excluded for all | Follow up 100388 | NA |
| 103494 | Study in Healthy Children (<2 Years) to Evaluate the Safety and Efficacy of GSK Biologicals' Live Attenuated Varicella Vaccine (VarilrixTM) and of GSK Biologicals' Combined Measles-Mumps-Rubella-Varicella Vaccine | http://clinicaltrials.gov/show/NCT00226499 | Excluded for all | Follow up 100388 | NA |
| 105239 | Booster Vaccination Study to Assess Immunogenicity & Safety of a Dose of GSK Biologicals' Mencevax™ ACWY & 1/5th of a Dose of Mencevax™ ACWY in Subjects Primed in the DTPW-HBV=HIB-MENAC-TT-011 Study | http://clinicaltrials.gov/show/NCT00291343 | Excluded for all | Follow up 100478 | NA |

| Study ID | Study title | Link | Exclusion | Reason | Inclusion per pathogen |
|---|---|---|---|---|---|
| 217744-076 | An open, multicenter, phase IV clinical trial to assess the immunogenicity and reactogenicity of three doses of GSK Biologicals' combined DTPa-HBV-IPV/Hib vaccine in healthy infants at 2, 4 and 6 months of age, when co-administered with Wyeth-Lederle's meningococcal group C conjugate vaccine. | NA | Included | Included | Diphtheria, HBV, HiB, IPV (1, 2, 3), Nmen (C), Pertussis-a (FHA, PRN, PT), Tetanus |
| 711202-001 | Evaluate immunogenicity, reactogenicity, safety of GSK Biologicals' MenC-TT vaccine (2 formulations) given with Infanrix hexa® + GSK Biologicals' Hib MenC-TT vaccine (2 formulations) given with Infanrix penta® to infants in mths 3,4,5 of life | http://clinicaltrials.gov/show/NCT00135486 | Included | Included | Diphtheria, HBV, HiB, IPV (1, 2, 3), Nmen (C), Pertussis-a (FHA, PRN, PT), Tetanus |
| 105245 | Booster Vaccination Study to Assess Immunogenicity & Safety of a Dose of GSK Biologicals' Mencevax™ ACWY & 1/5th of a Dose of Mencevax™ ACWY in Subjects Primed in the DTPW-HBV=HIB-MENAC-TT-011 Study | http://clinicaltrials.gov/show/NCT00291343 | Excluded for all | Follow up 100478 | NA |
| 104065 | Immune memory of GSK's DTPw-HBV/Hib vaccine by giving Plain PRP polysaccharide at 10 mths. Immuno & reacto of a booster dose of DTPw-HBV/Hib or DTPw-HBV or DTPw-HBV+Hib at 15-18 mths in infants previously primed with DTPw-HBV/Hib | http://clinicaltrials.gov/show/NCT00169442 | Excluded for all | Follow up 101222 | NA |
| 106602 | Immunogenicity, Reactogenicity & Safety of a Booster Dose of GSK Biologicals' DTPw-HBV/Hib Kft Vaccine Vs GSK Biologicals' DTPw-HBV/Hib Vaccine, in Infants Who Received a 3-Dose Primary Vaccination Course With the Same Vaccines. | http://clinicaltrials.gov/show/NCT00332566 | Excluded for all | Follow up 101223 | NA |
| 109810 | To assess long-term efficacy & safety of subjects approximately 3 years after priming with 2 doses of GlaxoSmithKline (GSK) Biologicals' oral live attenuated human rotavirus (HRV) vaccine (Rotarix) in the primary vaccination study (102247). | http://clinicaltrials.gov/show/NCT00420316 | Excluded for all | Follow up 102247 | NA |
| 111274 | Multi-Center Study to Assess the Efficacy, Safety and Immunogenicity of 2 or 3 Doses of GSK Biologicals' Oral Live Attenuated Human Rotavirus (HRV) Vaccine Given Concomitantly With Routine EPI Vaccinations in Healthy Infants | http://clinicaltrials.gov/show/NCT00241644 | Excluded for all | Follow up 102248 | NA |
| 104690 | Blinded, randomized study to evaluate the immunogenicity and safety of GlaxoSmithKline Biologicals' measles-mumps-rubella-varicella candidate vaccine given to healthy children during the second year of life | http://clinicaltrials.gov/show/NCT00127010 | Excluded for all | Follow up 103388 | NA |
| 111535 | Immunogenicity and reactogenicity study of GlaxoSmithKline Biologicals' Infanrix™/Hib vaccine administered as a booster dose to 18-24 months old children | http://clinicaltrials.gov/show/NCT00696423 | Excluded for all | Follow up 104567 | NA |
| 104730 | A phase III, multicentre booster vaccination study to assess the immunogenicity, safety and reactogenicity of a dose of Mencevax™ ACWY at 24 to 30 months of age in subjects primed with an investigational vaccination regimen in study 100480 and boosted at 15 to 24 months of age in study 104727. | http://clinicaltrials.gov/show/NCT00136604 | Excluded for all | Follow up 104727 | NA |
| 110478 | Immunogenicity and reactogenicity study of a new formulation of GSK Biologicals' DTPa-HBV-IPV/Hib vaccine administered as a booster dose to 18-23 months old children | http://clinicaltrials.gov/show/NCT00611559 | Excluded for all | Follow up 105910 | NA |
| 110031 | Phase II, observer-blind follow-up study to assess reacto-and immunogenicity of GSK Biologicals' pneumococcal conjugate vaccine (GSK1024850A), when given as booster in primed children or as 2-dose catch-up in unprimed children. | http://clinicaltrials.gov/show/NCT00513409 | Excluded for all | Follow up 106208 | NA |
| 111344 | Immunogenicity and reactogenicity of GSK Biologicals' DTPa-HBV-IPV/Hib vaccine when given as a booster dose. | http://clinicaltrials.gov/show/NCT00627458 | Excluded for all | Follow up 106786 | NA |
| 107137 | Prophylactic antipyretic treatment in children receiving booster dose of pneumococcal vaccine GSK1024850A and DTPa-HBV-IPV/Hib vaccine (Infanrix hexa) and assessment of impact of pneumococcal vaccination on nasopharyngeal carriage | http://clinicaltrials.gov/show/NCT00496015 | Excluded for all | Follow up 107017 | NA |
| 111345 | Long-term follow-up study to assess antibody persistence in children previously vaccinated with four doses of pneumococcal conjugate vaccine in primary vaccination study (105553) and booster vaccination study (107046) | http://clinicaltrials.gov/show/NCT00624819 | Excluded for all | Follow up 107046 | NA |
| 111346 | Assessment of long-term antibody persistence and immunological memory in children previously vaccinated with four pneumococcal conjugate vaccine doses and assessment of pneumococcal catch-up vaccination with GSK1024850A at 5 years of age | http://clinicaltrials.gov/show/NCT00624819 | Excluded for all | Follow up 107046 | NA |
| 111347 | Assessment of long-term antibody persistence and immunological memory in children previously vaccinated with four pneumococcal conjugate vaccine doses and assessment of pneumococcal catch-up vaccination with GSK1024850A at 5 years of age. | http://clinicaltrials.gov/show/NCT00624819 | Excluded for all | Follow up 107046 | NA |

| Study ID | Study title | Link | Exclusion | Reason | Inclusion per pathogen |
|---|---|---|---|---|---|
| 217744-076 | An open, multicenter, phase IV clinical trial to assess the immunogenicity and reactogenicity of three doses of GSK Biologicals’ combined DTPa-HBV-IPV/Hib vaccine in healthy infants at 2, 4 and 6 months of age, when co-administered with Wyeth-Lederle’s meningococcal group C conjugate vaccine. | NA | Included | Included | Diphtheria, HBV, HiB, IPV (1, 2, 3), Nmen (C), Pertussis-a (FHA, PRN, PT), Tetanus |
| 711202-001 | Evaluate immunogenicity, reactogenicity, safety of GSK Biologicals’ MenC-TT vaccine (2 formulations) given with Infanrix hexa® + GSK Biologicals’ Hib MenC-TT vaccine (2 formulations) given with Infanrix penta® to infants in mths 3,4,5 of life | http://clinicaltrials.gov/show/NCT00135486 | Included | Included | Diphtheria, HBV, HiB, IPV (1, 2, 3), Nmen (C), Pertussis-a (FHA, PRN, PT), Tetanus |
| 112801 | Vaccination course in children primed and boosted with pneumococcal vaccine GSK 1024850A and in age-matched unprimed children | http://clinicaltrials.gov/show/NCT00950833 | Excluded for all | Follow up 107137 | NA |
| 112909 | Booster vaccination with pneumococcal vaccine GSK1024850A in primed children and catch-up vaccination in unprimed children | http://clinicaltrials.gov/show/NCT01030822 | Excluded for all | Follow up 111188 | NA |
| 113266 | Evaluation of immunological persistence following 3-dose priming with GSK Biologicals’ 10-valent pneumococcal conjugate vaccine in study NCT00808444 and safety and immunogenicity following a booster dose of the same vaccine | http://clinicaltrials.gov/show/NCT01119625 | Excluded for all | Follow up 111654 | NA |
| 113978 | Immunogenicity and safety study of GlaxoSmithKline Biologicals’ GSK2202083A vaccine administered as a booster dose in 12-18 months old healthy children | http://clinicaltrials.gov/show/NCT01171989 | Excluded for all | Follow up 112157 | NA |
| 208136-039 | Follow-up to evaluate the the immunogenicity & safety of GSK Biologicals’ MMRV vaccine given as a two-dose schedule in the second year of life, as compared to separate administration of GSK Biologicals’ Priorix® & Varilrix®. | http://clinicaltrials.gov/show/NCT00406211 | Excluded for all | Follow up 208136-038 | NA |
| 208136-040 | Study to evaluate immunogenicity and safety of three production lots of GSK Biologicals' combined MeMuRu-OKA candidate vaccine given on a two-dose schedule to healthy children, as compared to separate administration of GSK Biologicals' Priorix® and Varilrix® vaccines | NA | Excluded for all | Follow up 208136-038 | NA |
| 208136-041 | Study to evaluate immunogenicity and safety of three production lots of GSK Biologicals' combined MeMuRu-OKA candidate vaccine given on a two-dose schedule to healthy children, as compared to separate administration of GSK Biologicals' Priorix® and Varilrix® vaccines | NA | Excluded for all | Follow up 208136-038 | NA |
| 104420 | A phase II study to evaluate the persistence of measles, mumps and rubella antibodies two years after the single dose primary vaccination in study 209762/151. | NA | Excluded for all | Follow up 209762-151 | NA |
| 217744-083 | Study to assess immunogenicity and reactogenicity of GSK Bio’s combined DTPa-HBV-IPV/Hib vaccine in pre-term infants in comparison with term infants, administered as a booster dose to children who previously were primed with 3 doses of GSK Biologicals combined DTPa-HBV-IPV/Hib vaccine | NA | Excluded for all | Follow up 217744-070 | NA |
| 101518 | A phase IV, open, multicentre study to assess the immunogenicity and reactogenicity of GSK Biologicals’ DTPa-HBV-IPV/Hib vaccine (Infanrix-hexa) given as a booster at 18-24 months of age to children who have received a three-dose primary immunisation course with the same vaccine in a previous study | NA | Excluded for all | Follow up 217744-090 | NA |
| 711202-008 | Evaluate the persistence and immune memory induced by a primary vaccination course with GSK Biologicals’ MenC-TT (1 formulation) & GSK Biologicals’ Hib-MenC-TT (2 formulations) or Meningitec™ in healthy toddlers aged 12-15 mths primed in study 711202/001 | http://clinicaltrials.gov/show/NCT00135564 | Excluded for all | Follow up 711202-001 | NA |
| 711866-005 | Open, 1-year, phase III, immunogenicity follow-up of subjects who previously received GSK Biologicals’ dTpa-IPV vaccine or GSK Biologicals’ dTpa (BoostrixTM) and Pasteur Mérieux’s IPV vaccine (IPV Mérieux() administered separately, at 4 to 8 years of age in study 711866/001 (dTpa-IPV-001) | NA | Excluded for all | Follow up 711866-001 | NA |
| 104756 | Booster Vaccination Study to Assess Safety & Reactogenicity of a Dose of DTPw-HBV/Hib Vaccine and to Assess the Immunogenicity, Safety & Reactogenicity of a Dose of Mencevax™ ACW in Subjects Primed in Study 759346/007 | http://clinicaltrials.gov/show/NCT00317109 | Excluded for all | Follow up 759346-007 | NA |
| 759348-002 | A randomized, controlled, phase II study to evaluate the safety and immunogenicity of 7 different formulations of an investigational vaccination regimen, when administered as a 3-dose primary immunization schedule before 6 months of age, followed by a fourth dose during the second year of life | NA | Excluded for all | Follow up 759348-001 | NA |
| 444563-022 | A phase II, double-blind, randomized, placebo-controlled study to assess the safety, reactogenicity and immunogenicity of three doses of GlaxoSmithKline (GSK) Biologicals’ oral live attenuated human rotavirus (HRV) vaccine | http://clinicaltrials.gov/show/NCT00263666 | Excluded for all | HIV babies | NA |

| Study ID | Study title | Link | Exclusion | Reason | Inclusion per pathogen |
|---|---|---|---|---|---|
| 217744-076 | An open, multicenter, phase IV clinical trial to assess the immunogenicity and reactogenicity of three doses of GSK Biologicals' combined DTPa-HBV-IPV/Hib vaccine in healthy infants at 2, 4 and 6 months of age, when co-administered with Wyeth-Lederle's meningococcal group C conjugate vaccine. | NA | Included | Included | Diphtheria, HBV, HiB, IPV (1, 2, 3), Nmen (C), Pertussis-a (FHA, PRN, PT), Tetanus |
| 711202-001 | Evaluate immunogenicity, reactogenicity, safety of GSK Biologicals' MenC-TT vaccine (2 formulations) given with Infanrix hexa® + GSK Biologicals' Hib MenC-TT vaccine (2 formulations) given with Infanrix penta® to infants in mths 3,4,5 of life | http://clinicaltrials.gov/show/NCT00135486 | Included | Included | Diphtheria, HBV, HiB, IPV (1, 2, 3), Nmen (C), Pertussis-a (FHA, PRN, PT), Tetanus |
| 115884 | Immunogenicity, safety and reactogenicity study of GSK Biologicals' pneumococcal vaccine (Synflorix™) when administered to children who are at an increased risk of pneumococcal infection | http://clinicaltrials.gov/show/NCT01746108 | Excluded for all | High-risk babies and no primary (Sp) | NA |
| 759346-009 | Study to evaluate immunogenicity, reactogenicity and safety of investigational vaccination regimen as compared to GSK Biological's Hiberix vaccine, extemporaneously mixed with GSK Biological's Tritanrix-HepB, when administered intramuscularly in infants at 6, 10 and 14 weeks of age | NA | Excluded for all | No data | NA |
| 102144 | Study to assess immunogenicity and reactogenicity of a booster dose of GSK Biologicals Kft's combined DTPwCSL-HB vaccine as compared to concomitant administration of CSL's DTPw vaccine and GSK Biologicals' hepatitis B vaccine at separate injection sites and to GSK Biologicals' DTPw-HB vaccine | NA | Excluded for all | No data | NA |
| 196131-001 | A phase III, partially double blind, randomized, multicentric study to evaluate the immunogenicity, safety and reactogenicity of GlaxoSmithKline (GSK) Biologicals Kft's combined DTPwCSL-HB vaccine as compared to Commonwealth Serum Laboratory's (CSL's) DTPwCSL (Triple Antigen™) and GSK Biologicals' HBV (Engerix™-B) administered concomitantly at separate injection sites and to GSK Biologicals' DTPw-HB vaccine (Tritanrix™-HepB) when administered to healthy infants at 3, 4 and 5 months of age | NA | Excluded for all | No data | NA |
| 208139-054 | Study in infants vaccinated with Engerix-B to evaluate immunogenicity and reactogenicity of following GSK Bio's vaccines: combined DTP-HB vaccine(10g HBsAg)combined DTP-HB vaccine(5g HBsAg) simultaneous administration of Engerix-B vaccine in right thigh and whole-cell DTP vaccine in left thigh | NA | Excluded for all | No data | NA |
| 209762-146 | Open, multicentre study to assess the safety, reactogenicity and immunogenicity of GlaxoSmithKline Biologicals live attenuated Measles-Mumps-Rubella vaccine (Priorix) given to healthy children at the age of 12 to 15 months or 4- 6 years | NA | Excluded for all | No data | NA |
| 113171 | Safety, reactogenicity and immunogenicity of GlaxoSmithKline (GSK) Biologicals' investigational vaccination regimen in children aged 12-23 months at the time of first vaccination. | http://clinicaltrials.gov/show/NCT00985751 | Excluded for all | No data | NA |
| 114174 | Impact of GSK Biologicals' 2189242A vaccine on nasopharyngeal carriage, safety and immunogenicity when co-administered with routine EPI vaccines in infants following safety assessment in children aged 2-4 years in The Gambia | http://clinicaltrials.gov/show/NCT01262872 | Excluded for all | No data | NA |
| 115373 | Safety and immunogenicity study of GSK Biologicals' pneumococcal vaccine 2830930A when administered as a single dose in healthy toddlers aged 12-23 months | http://clinicaltrials.gov/show/NCT01485406 | Excluded for all | No data | NA |
| 217744-086 | Single-blind, multicentre, phase IV clinical trial to assess and compare the immunogenicity and reactogenicity of GSK Biologicals' DTPa-HBV-IPV/Hib vaccine (Infanrix hexa™) and Aventis Pasteur's DTPa-HBV-IPV-Hib vaccine (Hexavac™) given as a primary vaccination course at 2, 4 and 6 months of age. | NA | Excluded for all | No dates file | NA |
| 217744-075 | Phase III, open, randomised immunogenicity and reactogenicity study to assess the interchangeability between GSK Bios' DTPa-HBV-IPV/Hib and DTPa-IPV/Hib + HBV at 3rd dose of primary vac. course in children who received HBV vac. at birth and one month of age and DTPa-IPV/Hib vac at 3-4 Mth of age | http://clinicaltrials.gov/show/NCT00366366 | Excluded for all | No dates file | NA |
| 217744-099 | A phase II, randomized, partially blinded clinical trial to evaluate the immunogenicity and reactogenicity of an investigational vaccine regimen of GSK Biologicals as compared to GSK Biologicals' DTPa-HBV-IPV/Hib vaccine (Infanrix™ hexa) and to the concomitant administration of GSK Biologicals' DTPa-HBV-IPV (Infanrix™ penta) and Hib (Hiberix™) vaccines, when given as a primary vaccination to healthy infants at 2, 3 and 4 months of age | NA | Excluded for all | No dates file | NA |
| 217744-049 | Immunogenicity and reactogenicity of GSK Biologicals' DTPa-HBV-IPV and Hib vaccines when administered concomitantly to healthy infants administered as a three-dose primary vaccination course at the age of 1.5, 3.5 and 6 months | http://clinicaltrials.gov/show/NCT00879827 | Excluded for all | No dates file | NA |
| 347414-016 | A phase II, single-blind, randomised, controlled, multicentre study to evaluate the safety, reactogenicity and immunogenicity of two experimental formulations versus a licensed Haemophilus influenzae type b (Hib) conjugate vaccine (Hiberix™ or HibTiter™) administered as primary vaccination to infants in their thirth, fourth and fifth months of life, with concomitant administration of SmithKline Beecham Biologicals' DTPa-HBV-IPV vaccine | NA | Excluded for all | No dates file | NA |

| Study ID | Study title | Link | Exclusion | Reason | Inclusion per pathogen |
|---|---|---|---|---|---|
| 217744-076 | An open, multicenter, phase IV clinical trial to assess the immunogenicity and reactogenicity of three doses of GSK Biologicals' combined DTPa-HBV-IPV/Hib vaccine in healthy infants at 2, 4 and 6 months of age, when co-administered with Wyeth-Lederle's meningococcal group C conjugate vaccine. | NA | Included | Included | Diphtheria, HBV, HiB, IPV (1, 2, 3), Nmen (C), Pertussis-a (FHA, PRN, PT), Tetanus |
| 711202-001 | Evaluate immunogenicity, reactogenicity, safety of GSK Biologicals' MenC-TT vaccine (2 formulations) given with Infanrix hexa® + GSK Biologicals' Hib MenC-TT vaccine (2 formulations) given with Infanrix penta® to infants in mths 3,4,5 of life | http://clinicaltrials.gov/show/NCT00135486 | Included | Included | Diphtheria, HBV, HiB, IPV (1, 2, 3), Nmen (C), Pertussis-a (FHA, PRN, PT), Tetanus |
| 217744-060 | Study to assess the immunogenicity and reactogenicity of DTPa-HBV-IPV mixed with Hib vaccine in healthy infants, followed by a dose of the same vaccine administered simultaneously with one dose of oral polio vaccine (OPV) | http://clinicaltrials.gov/show/NCT01457560 | Excluded for all | No dates file | NA |
| 763674-001 | A phase II, open, randomized, controlled study to evaluate an investigational vaccination regimen administered as athree dose primary vaccination course at 2, 3 and 4 months of age. | NA | Excluded for all | No dates file | NA |
| 213501-019 | Study to assess immunogenicity and reactogenicity of GSK Bio's quadrivalent diphtheria, tetanus, whole cell Bordetella pertussis, hepatitis B and Haemophilus influenzae type b conjugate vaccines when mixed extemporaneously and given in a single injection to healthy infants | NA | Excluded for all | No dates file | NA |
| 208108-087 | Phase 2 open randomized primary vaccination study to assess the immunogenicity and reactogenicity of GSK Biologicals' Haemophilus influenzae type b conjugate vaccine administered with commercially available DTPw vaccine as compared to GSK Biologicals' Hib administered mixed with GSK Biologicals' DTPw vaccine in healthy infants | NA | Excluded for all | No dates file | NA |
| 101853 | A randomized, controlled, phase II study to evaluate the safety and immunogenicity of GlaxoSmithKline Biologicals' investigational vaccination regimen, when administered intramuscularly as a 3-dose primary immunization (2-3-4 month schedule) before 6 months of age. | NA | Excluded for all | No dates file | NA |
| 347414-028 | A randomized, controlled, open, phase-II clinical study to evaluate the safety and immunogenicity of an experimental formulation extemporaneously mixed with SmithKline Beecham Biologicals' Haemophilus influenzae type b (Hib) vaccine, administered as a booster dose with DTPa-HBV-IPV to healthy children 12 to 18 months old | NA | Excluded for all | No dates file and no Ab (Nmen) | NA |
| 213503-049 | An open, multicentre, phase IV booster vaccination study to assess the immunogenicity and reactogenicity of GSKBiologicals combined Infanrix-IPV+Hib (DTPa-IPV/Hib) vaccine in healthy children aged 17 to 20 months whocompleted a three-dose primary vaccination course | NA | Excluded for all | No dates file and no primary (av 217744-076, but no PID) | NA |
| 115375 | Antibody persistence in children previously vaccinated with three doses of Infanrix hexa™ or Infanrix-IPV/Hib™ | http://clinicaltrials.gov/show/NCT01358825 | Excluded for all | No primary (av 105539, but no PID) | NA |
| 217744-095 | Study to assess immunogenicity and reactogenicity of GSK Bio's DTPa-HBV-IPV/Hib vaccine when given as a booster dose to children previously primed at 2-4-6 months of age either with Aventis Pasteurs DTPa-HBV-IPV-Hib vaccine or GSK Bio's DTPa-HBV-IPV/Hib vaccine in study DTPa-HBV-IPV-086 | NA | Excluded for all | No primary (av 217744-086 no dates) | NA |
| 763674-002 | A phase II open, randomized, controlled study to evaluate an investigational vaccination regimen administered as athree dose primary vaccination course at 2, 3 and 4 months of age. | NA | Excluded for all | No primary (av 763674-001, but no dates) | NA |
| 100448 | Long-Term Follow Up Study at Years 16-20, to Evaluate the Persistence of Immune Response of GlaxoSmithKline Biologicals' Hepatitis B Vaccine in Newborns of HBeAg+ and HBsAg+ Mothers | http://clinicaltrials.gov/show/NCT00240500 | Excluded for all | No primary (no av 103860-064) | NA |
| 103860-271 | Immunogenicity and protective efficacy of GlaxoSmithKline (GSK) recombinant-DNA hepatitis B vaccine (10 (g) in newborns of HBeAg+ and HBsAg+ mothers in comparison with a historical control group. | NA | Excluded for all | No primary (no av 103860-064) and no serology | NA |
| 106390 | Study to demonstrate non-inferiority of GSK Biologicals' Hib-MenC given with Infanrix™ penta versus NeisVac-C™ given with Infanrix™ hexa at 3, 5 months of age and persistence prior to a Hib-MenC booster at 11 months and immunogenicity of the booster | http://clinicaltrials.gov/show/NCT00327184 | Excluded for all | No primary (no av 106388) | NA |
| 109621 | Safety, reactogenicity and immunogenicity following booster dose of GSK Biologicals´ pneumococcal conjugate vaccine when co-administered with a booster dose of Infanrix-IPV/Hib in preterm born children at 16-18 months of age | http://clinicaltrials.gov/show/NCT00609492 | Excluded for all | No primary (no av 107737) and no serology | NA |
| 114843 | Safety and immunogenicity of a booster dose of new formulations of GlaxoSmithKline Biologicals' DTPa-HBV-IPV/Hib vaccine (GSK217744) | http://clinicaltrials.gov/show/NCT01453998 | Excluded for all | No primary (no av 113948) | NA |

| Study ID | Study title | Link | Exclusion | Reason | Inclusion per pathogen |
|---|---|---|---|---|---|
| 217744-076 | An open, multicenter, phase IV clinical trial to assess the immunogenicity and reactogenicity of three doses of GSK Biologicals' combined DTPa-HBV-IPV/Hib vaccine in healthy infants at 2, 4 and 6 months of age, when co-administered with Wyeth-Lederle's meningococcal group C conjugate vaccine. | NA | Included | Included | Diphtheria, HBV, HiB, IPV (1, 2, 3), Nmen (C), Pertussis-a (FHA, PRN, PT), Tetanus |
| 711202-001 | Evaluate immunogenicity, reactogenicity, safety of GSK Biologicals' MenC-TT vaccine (2 formulations) given with Infanrix hexa® + GSK Biologicals' Hib MenC-TT vaccine (2 formulations) given with Infanrix penta® to infants in mths 3,4,5 of life | http://clinicaltrials.gov/show/NCT00135486 | Included | Included | Diphtheria, HBV, HiB, IPV (1, 2, 3), Nmen (C), Pertussis-a (FHA, PRN, PT), Tetanus |
| 100565 | An open study to evaluate the immunogenicity, safety and reactogenicity of GlaxoSmithKline Biologicals' commercially available combined hepatitis A / hepatitis B vaccine (TWINRIX ADULT) containing 720 ELISA units of hepatitis A antigen and 20 µg of hepatitis B surface antigen, administered following a two-dose (0, 6 months) schedule in healthy children between the ages of 1 and 11 years | NA | Excluded for all | No primary (no av 208127-076) | NA |
| 100386 | Evaluate the persistence of immune response of GSK Biologicals' TWINRIX™ ADULT, administered according to 0,6 month schedule and 0,12 month schedule, in volunteers aged 12-15 years inclusive at the time of first vaccine dose | http://clinicaltrials.gov/show/NCT00197171 | Excluded for all | No primary (no av 208127-082) | NA |
| 100387 | Evaluate the persistence of immune response of GSK Biologicals' TWINRIX™ ADULT, administered according to 0,6 month schedule and 0,12 month schedule, in volunteers aged 12-15 years inclusive at the time of first vaccine dose | http://clinicaltrials.gov/show/NCT00197171 | Excluded for all | No primary (no av 208127-082) | NA |
| 100566 | Evaluate Persistence of Immune Response of GSK Biologicals' TWINRIX™ Vaccine Administered According to 0,6 Month Schedule Versus TWINRIX™ JUNIOR Administered According to 0,1,6 Month Schedule, in Subjects Aged 12-15 years at Time of First Vaccine Dose | http://clinicaltrials.gov/show/NCT00197119 | Excluded for all | No primary (no av 208127-084) | NA |
| 100567 | Evaluate Persistence of Immune Response of GSK Biologicals' TWINRIX™ Vaccine Administered According to 0,6 Month Schedule Versus TWINRIX™ JUNIOR Administered According to 0,1,6 Month Schedule, in Subjects Aged 12-15 years at Time of First Vaccine Dose | http://clinicaltrials.gov/show/NCT00197119 | Excluded for all | No primary (no av 208127-084) | NA |
| 100568 | Evaluate Persistence of Immune Response of GSK Biologicals' TWINRIX™ Vaccine Administered According to 0,6 Month Schedule Versus TWINRIX™ JUNIOR Administered According to 0,1,6 Month Schedule, in Subjects Aged 12-15 years at Time of First Vaccine Dose | http://clinicaltrials.gov/show/NCT00197119 | Excluded for all | No primary (no av 208127-084) | NA |
| 100569 | Evaluate Persistence of Immune Response of GSK Biologicals' TWINRIX™ Vaccine Administered According to 0,6 Month Schedule Versus TWINRIX™ JUNIOR Administered According to 0,1,6 Month Schedule, in Subjects Aged 12-15 years at Time of First Vaccine Dose | http://clinicaltrials.gov/show/NCT00197119 | Excluded for all | No primary (no av 208127-084) | NA |
| 100570 | Evaluate Persistence of Immune Response of GSK Biologicals' TWINRIX™ Vaccine Administered According to 0,6 Month Schedule Versus TWINRIX™ JUNIOR Administered According to 0,1,6 Month Schedule, in Subjects Aged 12-15 years at Time of First Vaccine Dose | http://clinicaltrials.gov/show/NCT00197119 | Excluded for all | No primary (no av 208127-084) | NA |
| 208355-124 | An open, 3.5 year, immunogenicity follow-up of subjects who previously received GSK Biologicals' dTpa vaccine or GSK Biologicals' DTPa vaccine or Chiron Behring's Td vaccine + either Pasteur Merieux's Pa vaccine or GSK Biologicals' pa vaccine, administered as a booster dose at age 4-6 years in study 208355/118 (APV-118) | NA | Excluded for all | No primary (no av 208355-118, but no dates) | NA |
| 208355-123 | A phase 3, open, multicenter study of the safety and immunogenicity of a booster dose of SB Bios' Diphtheria and Tetanus toxoids and acellular Pertussis vaccine and Pasteur Mérieux's Haemophilus influenzae type b conjugate vaccine when admin intramuscularly as separate injections(12-18 mths of age) | NA | Excluded for all | No primary (no av 217744-027) | NA |
| 217744-066 | Open, randomized, multicentre, phase III clinical trial to assess the reactogenicity and immunogenicity of a booster dose of GlaxoSmithKline (GSK) Biologicals' DTPa-HBV-IPV/Hib vaccine, co-administered during the same visit with GSK Biologicals' HAV vaccine (Havrix(), in children in their second ... | NA | Excluded for all | No primary (no av 217744-039, 041, 043 and 057) | NA |
| 217744-061 | Phase II, single-blinded, randomized comparative study of the safety and immunogenicity of a booster dose of SB Bios' DTPa-HBV-IPV vaccine compared with Infanrix when both vaccines are co-administered with OmniHIB between 15 and 18 months of age after primary immunization at the age of 2, 4 and 6 | NA | Excluded for all | No primary (no av 217744-044) | NA |
| 106744 | Multicentre study to assess persistence of antibodies against hepatitis B & immune response to a hepatitis B challenge dose in healthy children 7 to 9 yrs old previously vaccinated with 4 doses of GSK Biologicals' DTPa-HBV-IPV/Hib vaccine | http://clinicaltrials.gov/show/NCT00356564 | Excluded for all | No primary (no av 217744-048, 039, 058 and 059) | NA |
| 208127-122 | An open study to evaluate the immunogenicity and reactogenicity of GlaxoSmithKline Biologicals' combined hepatitis A / hepatitis B vaccine (schedule 0, 1, 6 months) in healthy children aged 1 to 6 years. | NA | Excluded for all | No primary (no av 3208127-042) | NA |
| 371594-006 | A phase II, open, randomized study to assess the immune memory induced by a primary vaccination course of an investigational vaccination regimen and the immunogenicity and reactogenicity of a fourth dose of an investigational vaccination regimen, to healthy toddlers primed in study 371594/004. | NA | Excluded for all | No primary (no av 371594-004) | NA |

| Study ID | Study title | Link | Exclusion | Reason | Inclusion per pathogen |
|---|---|---|---|---|---|
| 217744-076 | An open, multicenter, phase IV clinical trial to assess the immunogenicity and reactogenicity of three doses of GSK Biologicals' combined DTPa-HBV-IPV/Hib vaccine in healthy infants at 2, 4 and 6 months of age, when co-administered with Wyeth-Lederle's meningococcal group C conjugate vaccine. | NA | Included | Included | Diphtheria, HBV, HiB, IPV (1, 2, 3), Nmen (C), Pertussis-a (FHA, PRN, PT), Tetanus |
| 711202-001 | Evaluate immunogenicity, reactogenicity, safety of GSK Biologicals' MenC-TT vaccine (2 formulations) given with Infanrix hexa® + GSK Biologicals' Hib MenC-TT vaccine (2 formulations) given with Infanrix penta® to infants in mths 3,4,5 of life | http://clinicaltrials.gov/show/NCT00135486 | Included | Included | Diphtheria, HBV, HiB, IPV (1, 2, 3), Nmen (C), Pertussis-a (FHA, PRN, PT), Tetanus |
| 263855-035 | Phase IIIb study to evaluate immunogenicity, antibody persistency and reactogenicity of DTPa - INFANRIX and dTpa - BOOSTRIX vaccines administered to healthy children previously primed with 3 doses of DTPa vaccine compared to placebo (HAVRIX®JUNIOR) | http://clinicaltrials.gov/show/NCT00544271 | Excluded for all | No primary (Diphtheria, Tetanus, Pertussis), No Ab (HAV) | NA |
| 213503-046 | Open, randomised phase IIIb, clinical trial to compare the immunogenicity and reactogenicity of GSK Biologicals' DTaP-IPV vaccine, with GSK Biologicals' DTaP and Aventis Pasteur MSD's IPV vaccines administered to healthy children, previously vaccinated with 4 doses of DTaP and polio vaccines, and co-administered with GSK Biologicals' MMR vaccine | NA | Excluded for all | No primary (Measles, Mumps, Rubella, Diphtheria, Tetanus, Pertussis, Polio) | NA |
| 213503-047 | Open, randomized, phase II, clinical trial to compare the immunogenicity and safety of a booster dose of GSK Biologicals' DTaP-IPV vaccine (Infanrix®-IPV) co-administered with a booster dose of Merck and Company's M-M-R®II, to that of separate injections of GSK Biologicals' DTaP vaccine (Infanrix®), Aventis Pasteur's IPV (IPOL®) and M-M-R®II administered as booster doses to healthy children 4 to 6 years of age. | http://clinicaltrials.gov/show/NCT00263692 | Excluded for all | No primary (Measles, Mumps, Rubella, Diphtheria, Tetanus, Pertussis, Polio) | NA |
| 213503-048 | Safety, immunogenicity&consistency of 3 manufacturing lots of DTaP-IPV vaccine vs separate injections of GSK Biologicals' DTaP + Aventis Pasteur's IPV admd as booster doses to healthy children 4-6 yrs, each co-admd with Merck's MMR vaccine | http://clinicaltrials.gov/show/NCT00148941 | Excluded for all | No primary (Measles, Mumps, Rubella, Diphtheria, Tetanus, Pertussis, Polio) | NA |
| 111763 | Immunogenicity and safety of GSK Biologicals' dTpa-IPV vaccine (Boostrix Polio) as a booster dose in 3 and 4-year-old children | http://clinicaltrials.gov/show/NCT01245049 | Excluded for all | No primary (Measles, Mumps, Rubella, Diphtheria, Tetanus, Pertussis, Polio) | NA |
| 105909 | Comparative study evaluating the immunogenicity & safety of MeMuRu-OKA vaccine & measles-mumps-rubella vaccine (Priorix™) co-administered with varicella vaccine (Varilrix™) in children primed with both measles-mumps-rubella & varicella vaccines | http://clinicaltrials.gov/show/NCT00352898 | Excluded for all | No primary (Measles, Mumps, Rubella, VZV) | NA |
| 115158 | Immunogenicity and safety study of GSK Biologicals' combined measles-mumps-rubella vaccine in subjects four to six years of age (209762) | http://clinicaltrials.gov/show/NCT01621802 | Excluded for all | No primary (Measles, Mumps, Rubella, VZV, Diphtheria, Tetanus, Pertussis, Polio) | NA |
| 113264 | Immunogenicity and reactogenicity of a booster dose of GlaxoSmithKline Biologicals' GSK2036874A vaccine in healthy toddlers | http://clinicaltrials.gov/show/NCT01106092 | Excluded for all | No primary (routine) | NA |
| 110474 | Long-term persistence of hepatitis B antibodies & immune response to a hepatitis B vaccine challenge in 7-8 year old children, previously vaccinated in infancy with GlaxoSmithKline (GSK) Biologicals' HBV vaccine. | http://clinicaltrials.gov/show/NCT00519649 | Excluded for all | No primary (routine) | NA |

| Study ID | Study title | Link | Exclusion | Reason | Inclusion per pathogen |
|---|---|---|---|---|---|
| 217744-076 | An open, multicenter, phase IV clinical trial to assess the immunogenicity and reactogenicity of three doses of GSK Biologicals' combined DTPa-HBV-IPV/Hib vaccine in healthy infants at 2, 4 and 6 months of age, when co-administered with Wyeth-Lederle's meningococcal group C conjugate vaccine. | NA | Included | Included | Diphtheria, HBV, HiB, IPV (1, 2, 3), Nmen (C), Pertussis-a (FHA, PRN, PT), Tetanus |
| 711202-001 | Evaluate immunogenicity, reactogenicity, safety of GSK Biologicals' MenC-TT vaccine (2 formulations) given with Infanrix hexa® + GSK Biologicals' Hib MenC-TT vaccine (2 formulations) given with Infanrix penta® to infants in mths 3,4,5 of life | http://clinicaltrials.gov/show/NCT00135486 | Included | Included | Diphtheria, HBV, HiB, IPV (1, 2, 3), Nmen (C), Pertussis-a (FHA, PRN, PT), Tetanus |
| 213503-050 | Study to evaluate safety and immunogenicity of a booster dose of DTPa-IPV/Hib at 18 months of age with GSK Biologicals DTPa-IPV/Hib compared to Aventis Pasteurs Pentacel, after an initial primary vaccination series administered at 2, 4, and6 months of age with Aventis Pasteurs Pentacel | NA | Excluded for all | No primary (routine) | NA |
| 213503-045 | Study to compare immunogenicity and reactogenicity of GSK Bio's DTPa-IPV vaccine, with GSK Bio's DTPa (Infanrix) and Aventis MSDs IPV vaccine (Imovax Polio) administered separately to healthy children 4 to 6 years of age | NA | Excluded for all | No primary (routine) | NA |
| 106789 | Assess long-term persistence of hepatitis B antibodies & immune response to a hepatitis B vaccine (Engerix-B Kinder) challenge in children aged 4-5 yrs (previously primed & boosted in the 1st 2 yrs of life with DTPa-HBV-IPV/Hib vaccine) | http://clinicaltrials.gov/show/NCT00411697 | Excluded for all | No primary (routine) | NA |
| 711866-006 | A phase III, open study to evaluate the immunogenicity, safety and reactogenicity of GSK Bios' dTpa-IPV vaccine when administered as a booster vaccination to healthy children (6-8 Y) and previously vaccinated with 4 doses of DTPw and at least 3 doses of OPV/ IPV vaccines | NA | Excluded for all | No primary (routine) | NA |
| 711866-001 | A study to assess the lot-to-lot consistency of GSK Biologicals dTpa-IPV vaccine administered to healthy children 4 to 8 years of age, previously vaccinated with four doses of DTPa or DTPa-based combination vaccines and at least 3 doses of OPV or IPV | NA | Excluded for all | No primary (routine) | NA |
| 201532 | Evaluation of immunogenicity, safety and reactogenicity of GSK Biologicals' dTpa booster vaccine (263855) (Boostrix) administered as a booster dose in healthy Russian subjects | http://clinicaltrials.gov/show/NCT03311659 | Excluded for all | No primary (routine) | NA |
| 104005 | Phase IIIb, open, randomized, multicenter study to assess the immunogenicity & safety of GSK Biologicals' combined DTPa-HBV-IPV/Hib vaccine in Indian infants when given at 6-10-14 weeks of age or at 2-4-6 months of age | http://clinicaltrials.gov/show/NCT00316147 | Excluded for all | No sero file | NA |
| 105987 | A single-blind, randomized, controlled, multinational study for the evaluation of safety of GlaxoSmithKline (GSK) Biologicals' investigational vaccination regimen compared to monovalent Haemophilus influenzae type b (Hib) control vaccine in healthy infants at 2, 4, 6, and 12 to 15 months of age. | http://clinicaltrials.gov/show/NCT00345579 | Excluded for all | No sero file | NA |
| 103812 | Assess reactogenicity & safety of a booster of either Tritanrix™-HepB/Hib-MenAC or Tritanrix™-HepB/Hiberix™ given (single-blind) at 15-18 (Philippines)/15-24 mths (Thailand) & a dose of Mencevax™ ACWY at 24-30 mths (open label) | http://clinicaltrials.gov/show/NCT00228917 | Excluded for all | No sero file | NA |
| 104171 | Assess reactogenicity & safety of a booster of either Tritanrix™-HepB/Hib-MenAC or Tritanrix™-HepB/Hiberix™ given (single-blind) at 15-18 (Philippines)/15-24 mths (Thailand) & a dose of Mencevax™ ACWY at 24-30 mths (open label) | http://clinicaltrials.gov/show/NCT00228917 | Excluded for all | No sero file | NA |
| 100791 | Study to assess immunogenicity and non-inferiority of investigational vaccination regimen as compared to Tritanrix-HepB/Hiberix and as compared to Meningitec when administered to healthy infants | http://clinicaltrials.gov/show/NCT00317187 | Excluded for all | No sero file | NA |
| 200147 | Safety and immunogenicity study of 2 formulations of GSK Biologicals' varicella vaccines given as a 2-dose course in the second year of life. | http://clinicaltrials.gov/show/NCT02570126 | Excluded for all | No sero file<br>No primary (Measles, Mumps, Rubella) | NA |
| 205249 | A Phase 3, Open Label, Randomized, Controlled, Multi-Center Study to Evaluate the Safety and Immunogenicity of GSK Biologicals' Meningococcal B Recombinant Vaccine When Administered concomitantly with Routine Vaccines to Healthy Infants in Taiwan. | http://clinicaltrials.gov/show/NCT02173704 | Excluded for all | Novartis study | NA |
| 205240 | A Phase 3b, Open-Label, Randomized, Multicenter Study to Assess the Safety and Immunogenicity of GlaxoSmithKline Biologicals Meningococcal group B Vaccine When Administered Concomitantly with GlaxoSmithKline Biologicals MenACWY Conjugate Vaccine to Healthy Infants | http://clinicaltrials.gov/show/NCT02106390 | Excluded for all | Novartis study | NA |
| 205450 | A Phase 3, Partially Blinded, Randomized, Multi-Center, Controlled Study to Evaluate Immunogenicity, Safety and Lot to Lot Consistency of Novartis Meningococcal B Recombinant Vaccine When Administered With Routine Infant Vaccinations to Healthy Infants | http://clinicaltrials.gov/show/NCT00657709 | Excluded for all | Novartis study. No sero file. | NA |

| Study ID | Study title | Link | Exclusion | Reason | Inclusion per pathogen |
| --- | --- | --- | --- | --- | --- |
| 217744-076 | An open, multicenter, phase IV clinical trial to assess the immunogenicity and reactogenicity of three doses of GSK Biologicals' combined DTPa-HBV-IPV/Hib vaccine in healthy infants at 2, 4 and 6 months of age, when co-administered with Wyeth-Lederle's meningococcal group C conjugate vaccine. | NA | Included | Included | Diphtheria, HBV, HiB, IPV (1, 2, 3), Nmen (C), Pertussis-a (FHA, PRN, PT), Tetanus |
| 711202-001 | Evaluate immunogenicity, reactogenicity, safety of GSK Biologicals' MenC-TT vaccine (2 formulations) given with Infanrix hexa® + GSK Biologicals' Hib MenC-TT vaccine (2 formulations) given with Infanrix penta® to infants in mths 3,4,5 of life | http://clinicaltrials.gov/show/NCT00135486 | Included | Included | Diphtheria, HBV, HiB, IPV (1, 2, 3), Nmen (C), Pertussis-a (FHA, PRN, PT), Tetanus |
| 205453 | A Phase 3, Open Label, Multi-Center, Extension Study to Evaluate the Safety, Tolerability and Immunogenicity of Novartis Meningococcal B Recombinant Vaccine When Administered as a Booster at 12 Months of Age or as a Two-dose Catch-up to Healthy Toddlers Who Participated in Study V72P13 | http://clinicaltrials.gov/show/NCT01139021 | Excluded for all | Novartis study. No sero file. | NA |
| 205399 | A Phase 2b, Open Label, Randomized, Parallel-Group, Multi-Center Study to Evaluate the Safety, Tolerability and Immunogenicity of Novartis Meningococcal B Recombinant Vaccine When Administered With or Without Routine Infant Vaccinations to Healthy Infants According to Different Immunization Schedules. | http://clinicaltrials.gov/show/NCT00721396 | Excluded for all | Novartis study. No sero file. | NA |
| 106481 | Phase IIIb, Double Blind, Randomised, Placebo-Controlled, Multi-Country/Centre, Study to Assess Safety, Reactogenicity & Immunogenicity of 2 Doses of GSK Biologicals' Oral Live Attenuated Human Rotavirus (HRV) Vaccine in Pre-Term Infants | http://clinicaltrials.gov/show/NCT00420745 | Excluded for all | Pre-term babies | NA |
| 217744-090 | A phase IV, open, multicentre study to assess the immunogenicity and reactogenicity of GlaxoSmithKline Biologicals' DTPa-HBV-IPV/Hib vaccine (Infanrix hexa() given as a primary vaccination course at 2, 4 and 6 months of age to pre-term infants. | NA | Excluded for all | Pre-term babies | NA |
| 103860-280 | Long-term study of immune response persistence of GSK Biologicals' 2-dose thiomersal-free Engerix™-B and 3-dose preservative-free Engerix™-B vaccines in subjects aged 11-15 yrs | http://clinicaltrials.gov/show/NCT00343915 | Excluded for all | Teenagers | NA |
| 101695 | Long-term study of immune response persistence of GSK Biologicals' 2-dose thiomersal-free Engerix™-B and 3-dose preservative-free Engerix™-B vaccines in subjects aged 11-15 yrs | http://clinicaltrials.gov/show/NCT00343915 | Excluded for all | Teenagers and no primary (av 103860-280) | NA |
| 101696 | Long-term study of immune response persistence of GSK Biologicals' 2-dose thiomersal-free Engerix™-B and 3-dose preservative-free Engerix™-B vaccines in subjects aged 11-15 yrs | http://clinicaltrials.gov/show/NCT00343915 | Excluded for all | Teenagers and no primary (av 103860-280) | NA |
| 101697 | Long-term study of immune response persistence of GSK Biologicals' 2-dose thiomersal-free Engerix™-B and 3-dose preservative-free Engerix™-B vaccines in subjects aged 11-15 yrs | http://clinicaltrials.gov/show/NCT00343915 | Excluded for all | Teenagers and no primary (av 103860-280) | NA |
| 101698 | Long-term study of immune response persistence of GSK Biologicals' 2-dose thiomersal-free Engerix™-B and 3-dose preservative-free Engerix™-B vaccines in subjects aged 11-15 yrs | http://clinicaltrials.gov/show/NCT00343915 | Excluded for all | Teenagers and no primary (av 103860-280) | NA |
| 107625 | Efficacy, safety, reactogenicity and immunogenicity study of the lyophilised formulation of Rotarix vaccine in healthy Japanese infants | http://clinicaltrials.gov/show/NCT00480324 | Excluded for all | No Ab (Diphtheria, Tetanus, Pertussis, HBV) | NA |
| 111188 | Primary vaccination course in healthy children receiving the pneumococcal vaccine GSK 1024850A co-administered with Tritanrix™-HepB/Hib at 6, 10 and 14 weeks of age | http://clinicaltrials.gov/show/NCT00814710 | Excluded for all | No primary (HBV), No Ab baseline (Diphtheria, Tetanus, Pertussis, HiB) | NA |
| 113615 | Feasibility study of GlaxoSmithKline Biologicals' GSK2202083A vaccine in healthy infants at 2, 4 and 12 months of age | http://clinicaltrials.gov/show/NCT01090453 | Excluded for all | No Ab (Rotavirus), No Ab baseline (Diphtheria, Tetanus, Pertussis, HBV, HiB, Nmen, Polio, Sp) | NA |
| 107005 | To assess safety, reactogenicity and immunogenicity of GSK Biologicals' 10-valent pneumococcal conjugate vaccine, when co-administered with DTPa-combined vaccines and MenC or Hib-MenC vaccines during the first 6 months of age. | http://clinicaltrials.gov/show/NCT00334334 | Excluded for all | No primary (HBV), No Ab baseline (Diphtheria, Tetanus, Pertussis, | NA |

| Study ID | Study title | Link | Exclusion | Reason | Inclusion per pathogen |
|---|---|---|---|---|---|
| 217744-076 | An open, multicenter, phase IV clinical trial to assess the immunogenicity and reactogenicity of three doses of GSK Biologicals' combined DTPa-HBV-IPV/Hib vaccine in healthy infants at 2, 4 and 6 months of age, when co-administered with Wyeth-Lederle's meningococcal group C conjugate vaccine. | NA | Included | Included | Diphtheria, HBV, HiB, IPV (1, 2, 3), Nmen (C), Pertussis-a (FHA, PRN, PT), Tetanus |
| 711202-001 | Evaluate immunogenicity, reactogenicity, safety of GSK Biologicals' MenC-TT vaccine (2 formulations) given with Infanrix hexa® + GSK Biologicals' Hib MenC-TT vaccine (2 formulations) given with Infanrix penta® to infants in mths 3,4,5 of life | http://clinicaltrials.gov/show/NCT00135486 | Included | Included | Diphtheria, HBV, HiB, IPV (1, 2, 3), Nmen (C), Pertussis-a (FHA, PRN, PT), Tetanus |
| | | | | HiB, Nmen, Polio, Sp) | |
| 105539 | An open, randomized, phase IIIa study to evaluate the safety and immunogenicity of GSK Biologicals' 10-valent pneumococcal conjugate vaccine, when administered intramuscularly according to a 2-4-11 months vaccination schedule | http://clinicaltrials.gov/show/NCT00307034 | Excluded for all | No Ab baseline (Diphtheria, Tetanus, Pertussis, HBV, HiB, Polio) | NA |
| 759348-003 | A randomized, controlled, phase II study to evaluate the safety and immunogenicity of four different formulations of GlaxoSmithKline (GSK) Biologicals' investigational vaccination regimen, when administered as a 3-dose primary immunization schedule beginning before 6 months of age. | NA | Excluded for all | No Ab baseline (Diphtheria, Tetanus, Pertussis, HBV, HiB, Polio) | NA |
| 111654 | Non-inferiority of a commercial lot of the pneumococcal vaccine GSK1024850A compared to a clinical lot. | http://clinicaltrials.gov/show/NCT00808444 | Excluded for all | No Ab baseline (Diphtheria, Tetanus, Pertussis, HiB, Polio, Rotavirus, Sp), No primary (HBV) | NA |
| 347414-008 | Study to assess immunogenicity and safety of GSK Biologicals' investigational vaccination regimen administered concomitantly in separate injections with GSK Biologicals' DTPa-IPV/Hib vaccine as a primary vaccination course to healthy infants at 2-4-6 months of age and as a booster dose at 12 to 18 months of age | NA | Excluded for all | No Ab (Diphtheria, Tetanus, Pertussis, HBV, Polio) | NA |
| 106445 | Study to demonstrate non-inferiority of GSK Biologicals' Hib-MenC with Priorix™, versus MenC-CRM197 vaccine with Hiberix™ & Priorix™ in toddlers primed with Hib but not MenC & to evaluate persistence up to 5 years after vaccination. | http://clinicaltrials.gov/show/NCT00326118 | Excluded for all | No Ab (Measles, Mumps, Rubella), No primary (HiB, Diphtheria, Tetanus, Pertussis) | NA |
| 107058 | Evaluate immunogenicity, safety & reactogenicity of GSK Biologicals' 10-valent pneumococcal conjugate vaccine given as catch-up immunization in children older than 7 mo of age or as 3-dose primary immunization in children before 6 mo of age | http://clinicaltrials.gov/show/NCT00345358 | Excluded for all | No Ab baseline (Diphtheria, Tetanus, Pertussis, HiB, Polio) | NA |
| 347414-020 | A Phase III, randomized, single blind study to assess the immune response induced by Prevenar™(Wyeth Lederle) when administered to healthy infants with DTPa-HBV-IPV/Hib, compared to an investigational vaccination regimen as a 3-dose primary vaccination course at a monthly interval starting at 8–16 Weeks of age | NA | Excluded for all | No primary (HBV) | NA |
| 112640 | Immunogenicity, safety and reactogenicity of GlaxoSmithKline Biologicals' pneumococcal vaccine GSK1024850A following primary and booster vaccination of healthy Japanese children | http://clinicaltrials.gov/show/NCT01027845 | Excluded for all | No primary (HBV), No vaccine concentration (HiB, Pertussis) | NA |
| 217744-069 | Immunogenicity and safety of GSK Biological's DTPa-HBV-IPV/Hib vaccine or DTPa-IPV/Hib co-administered with HBV vaccine as primary and booster vaccination in healthy infants born to hepatitis B surface antigen negative mothers | http://clinicaltrials.gov/show/NCT00880477 | Excluded for all | No Ab baseline (HBV, Polio) | NA |
| 217744-077 | A phase III, double-blind, randomized, multicenter primary vaccination study to bridge the DTPa-HBV-IPV vaccine manufactured according to the large scale manufacturing process with the DTPa-HBV-IPV vaccine manufactured by the small scale manufacturing process when administered intramuscularly to infants at 2, 4 and 6 months of age, co-administered with Merck's Hib conjugate vaccine (Liquid PedvaxHIB®) in a separate injection at 2 and 4 months of age | NA | Excluded for all | No Ab (HiB) | NA |
| 208136-022 | Study to evaluate immunogenicity and safety of one dose of GSK Bios' combined measles-mumps-rubella-varicella vaccine compared to concomitant administrations of GSK Bios' measles-mumps-rubella vaccine and varicella vaccine | NA | Excluded for all | No primary routine (Measles) | NA |

| Study ID | Study title | Link | Exclusion | Reason | Inclusion per pathogen |
|---|---|---|---|---|---|
| 217744-076 | An open, multicenter, phase IV clinical trial to assess the immunogenicity and reactogenicity of three doses of GSK Biologicals' combined DTPa-HBV-IPV/Hib vaccine in healthy infants at 2, 4 and 6 months of age, when co-administered with Wyeth-Lederle's meningococcal group C conjugate vaccine. | NA | Included | Included | Diphtheria, HBV, HiB, IPV (1, 2, 3), Nmen (C), Pertussis-a (FHA, PRN, PT), Tetanus |
| 711202-001 | Evaluate immunogenicity, reactogenicity, safety of GSK Biologicals' MenC-TT vaccine (2 formulations) given with Infanrix hexa® + GSK Biologicals' Hib MenC-TT vaccine (2 formulations) given with Infanrix penta® to infants in mths 3,4,5 of life | http://clinicaltrials.gov/show/NCT00135486 | Included | Included | Diphtheria, HBV, HiB, IPV (1, 2, 3), Nmen (C), Pertussis-a (FHA, PRN, PT), Tetanus |
| 110058 | Immunogenicity of GlaxoSmithKline Biologicals' MMRV vaccine (208136) vs. ProQuad®, when coadministered with hepatitis A and pneumococcal conjugate vaccines to children 12-14 months of age. | http://clinicaltrials.gov/show/NCT00578175 | Excluded for all | No Ab method (VZV, Mumps), No primary (Sp) | NA |
| 111870 | A phase II, randomized, observer blind, controlled, multicenter study to assess immunogenicity and antibody persistence following vaccination with GSK's candidate combined measles, mumps, and rubella vaccine (MMR) versus M-M-R® II as a first dose, both administered subcutaneously at 12-15 months of age, concomitantly with hepatitis A vaccine (HAV), varicella vaccine (VV) and pneumococcal conjugate vaccine (PCV) but at separate sites. | http://clinicaltrials.gov/show/NCT00861744 | Excluded for all | No Ab method (VZV, Mumps), No primary (Sp) | NA |
| 103860-277 | Phase III study of immunogenicity and safety of 3 doses of GSK Biologicals' thimerosal-free hepatitis B vaccine compared to the US-licensed GSK Biologicals' preservative-free hepatitis B vaccine when administered intramuscularly on a 0, 1, 6-month schedule to healthy infants in their first two weeks of life | NA | Excluded for all | Outside age range (HBV) | NA |
| 402764-004 | A phase III, open, randomized study to evaluate the immunogenicity and reactogenicity of investigational vaccination regimens | NA | Excluded for all | Outside age range (Diphtheria, Tetanus, Pertussis, Nmen, HBV, HiB) | NA |
| 213501-018 | Study to evaluate immunogenicity, safety and reactogenicity of two different immunization regimens against hepatitis B, diphtheria, tetanus, pertussis and Haemophilus influenzae type b (Hib)diseases in healthy infants primed with a birth dose of GSK Biologicals hepatitis B | NA | Excluded for all | Outside age range (Diphtheria, Tetanus, Pertussis, HBV, HiB) | NA |
| 104977 | Non-inferiority of one formulation of GSK Biologicals' DTPw-HBV/Hib to 2 formulations of GSK Biologicals' DTPw-HBV/Hib with respect to the immune response to the PRP antigen, when administered to healthy infants at 6, 10, 14 weeks of age | http://clinicaltrials.gov/show/NCT00473668 | Excluded for all | Latitude range <80% (Diphtheria, Tetanus, Pertussis, HBV, HiB) | NA |
| 104567 | Phase IIIb, multicentre study to assess safety & immunogenicity of GSK Biologicals' combined DTPa/Hib (Infanrix/Hib) vaccine vs separate administration of DTPa (Infanrix) & Hib (Hiberix) vaccines in healthy infants 3,4,&5 months of age as compared with the separate administration of DTPa and Hib vaccines at different injection sites. | http://clinicaltrials.gov/show/NCT00412854 | Excluded for all | Latitude range <80% (Diphtheria, Tetanus, Pertussis, HiB) | NA |
| 792014-001 | A phase II, open (partially double-blind), randomised, controlled, multicentre, primary vaccination study to evaluate the immunogenicity (including immune memory), reactogenicity and safety of three different formulations of the GSK Biologicals' combined Haemophilus influenzae type b-meningococcal serogroups CY conjugate vaccine given concomitantly with Infanrix® penta and Prevenar®, versus ActHIB® and Meningitec® given concomitantly with Infanrix® penta and versus ActHIB® given concomitantly with Infanrix® penta and Prevenar® in infants according to a 2-4-6 month schedule. | http://clinicaltrials.gov/show/NCT00127855 | Excluded for all | Latitude range <80% (Diphtheria, Tetanus, Pertussis, Nmen, HBV, HiB, Polio, Sp) | NA |
| 103792 | A multicenter study of the immunogenicity & safety of 2 doses of GSK Biologicals' oral live attenuated human rotavirus vaccine (RIX4414) as primary dosing of healthy infants in India aged approximately 8 wks at the time of the first dose | http://clinicaltrials.gov/show/NCT00289172 | Excluded for all | Latitude range <80% (Rotavirus) | NA |
| 115992 | Two-dose primary vaccination with either GSK Biologicals' 10-valent pneumococcal vaccine (Synflorix™) or Pfizer's Prevenar 13™ or both vaccines followed by a booster dose of Synflorix™ | http://clinicaltrials.gov/show/NCT01641133 | Excluded for all | Latitude range <80% (Sp) | NA |

RCT = Randomized controlled trial, Ab = Antibody immunogenicity, HAV = Hepatitis A virus, HBV = Hepatitis B virus, HiB = *Haemophilus influenzae Type B*, VZV = Varicella Zoster virus, Sp = *Streptococcus pneumoniae*, Nmen = *Neisseria meningitidis*, FHA = filamentous hemagglutinin, PRN = pertactin; PT = pertussis toxoid, NA = not applicable.

**Stable 2. Summary of analytical methods, target antigens, and serological thresholds for immunogenicity assessment.** Overview of the laboratory assays and predefined cut-off values used to evaluate the immune response for each pathogen included in the studies.

| Pathogen | Method | Antigen | Cut-off | Unit |
|---|---|---|---|---|
| Diphtheria | ELISA | Anti-diphtheria | 0.10 | IU/mL |
| | PRNT | Anti-diphtheria | 0.016 | IU/mL |
| HAV | ELISA | Anti-hav | 15.00 | mIU/mL |
| HBV | ELISA | Anti-hbv | 10.00 | mIU/mL |
| *H. influenzae type B* | ELISA | Anti-hib | 0.15 | µg/mL |
| Polio (Inactive) | PRNT | Anti-polio 1 | 8.00 | ED50 |
| | PRNT | Anti-polio 2 | 8.00 | ED50 |
| | PRNT | Anti-polio 3 | 8.00 | ED50 |
| Measles | ELISA | Anti-measles | 150.00 | mIU/mL |
| *N. meningitidis* | ELISA | Anti-men A | 0.30 | µg/mL |
| | ELISA | Anti-men C | 0.30 | µg/mL |
| | BAC | Anti-men A | 8.00 | Dilution |
| | BAC | Anti-men C | 8.00 | Dilution |
| Mumps | ELISA | Anti-mumps | 231.00 | U/mL |
| | PRNT | Anti-mumps | 24.00 | ED50 |
| Pertussis (acellular) | ELISA | Anti-pertussis PT | 15.00 | EU/mL |
| | ELISA | Anti-pertussis FHA | 5.00 | EU/mL |
| | ELISA | Anti-pertussis PRN | 5.00 | EU/mL |
| Pertussis (whole) | ELISA | Anti-pertussis PT | 15.00 | EU/mL |
| Rotavirus | ELISA | Anti-rota IgA | 20.00 | U/mL |
| Rubella | ELISA | Anti-rubella | 4.00 | IU/mL |
| *S. pneumoniae* | ELISA | Anti-strep 01 | 0.05 | µg/mL |
| | ELISA | Anti-strep 03 | 0.05 | µg/mL |
| | ELISA | Anti-strep 04 | 0.05 | µg/mL |
| | ELISA | Anti-strep 05 | 0.05 | µg/mL |
| | ELISA | Anti-strep 06a | 0.05 | µg/mL |
| | ELISA | Anti-strep 06b | 0.05 | µg/mL |
| | ELISA | Anti-strep 07f | 0.05 | µg/mL |
| | ELISA | Anti-strep 09v | 0.05 | µg/mL |
| | ELISA | Anti-strep 14 | 0.05 | µg/mL |
| | ELISA | Anti-strep 18c | 0.05 | µg/mL |
| | ELISA | Anti-strep 19a | 0.05 | µg/mL |
| | ELISA | Anti-strep 19f | 0.05 | µg/mL |
| | ELISA | Anti-strep 23f | 0.05 | µg/mL |
| | OPA | Anti-strep 01 | 8.00 | Dilution |
| | OPA | Anti-strep 03 | 8.00 | Dilution |
| | OPA | Anti-strep 04 | 8.00 | Dilution |
| | OPA | Anti-strep 05 | 8.00 | Dilution |
| | OPA | Anti-strep 06a | 8.00 | Dilution |
| | OPA | Anti-strep 06b | 8.00 | Dilution |
| | OPA | Anti-strep 07f | 8.00 | Dilution |
| | OPA | Anti-strep 09v | 8.00 | Dilution |
| | OPA | Anti-strep 14 | 8.00 | Dilution |
| | OPA | Anti-strep 18c | 8.00 | Dilution |
| | OPA | Anti-strep 19a | 8.00 | Dilution |
| | OPA | Anti-strep 19f | 8.00 | Dilution |
| | OPA | Anti-strep 23f | 8.00 | Dilution |
| Tetanus | ELISA | Anti-tetanus | 0.10 | IU/mL |
| VZV | IFA | Anti-vzv | 4.00 | mIU/mL |

HAV = Hepatitis A virus, HBV = Hepatitis B virus, VZV = Varicella Zoster virus, FHA = filamentous hemagglutinin, PRN = pertactin, PT = pertussis toxoid, ELISA = Enzyme-linked immunosorbent assay, OPA = Opsonophagocytosis assay, PRNT = Plaque reduction neutralization test, IFA = Immunofluorescence assay, BAC = Serum bactericidal assay.

**Stable 3. Randomized clinical trials and the number of participants included in each pathogen-antigen subset.**

| Pathogen | Antigen | Method | Doses | Studies | Participants |
|---|---|---|---|---|---|
| **Main analyses** | | | | | |
| Diphtheria | Anti-diphtheria | ELISA | 3 | 28 | 6267 |
| HAV | Anti-hav | ELISA | 1 | 1 | 1022 |
| HBV | Anti-hbv | ELISA | 3 | 8 | 2043 |
| *H. influenzae type B* | Anti-hib | ELISA | 3 | 27 | 5516 |
| Polio (Inactive) | Anti-polio 1 | PRNT | 3 | 15 | 3343 |
| Measles | Anti-measles | ELISA | 1 | 24 | 20845 |
| *N. meningitidis* | Anti-men C | ELISA | 3 | 15 | 4903 |
| Mumps | Anti-mumps | ELISA | 1 | 21 | 11694 |
| Pertussis (whole-cell) | Anti-pertussis PT | ELISA | 3 | 13 | 4102 |
| Rotavirus | Anti-rota IgA | ELISA | 2–3 | 29 | 7191 |
| Rubella | Anti-rubella | ELISA | 1 | 24 | 20847 |
| *S. pneumoniae* | Anti-strep 04 | ELISA | 3 | 19 | 7358 |
| Tetanus | Anti-tetanus | ELISA | 3 | 29 | 2133 |
| VZV | Anti-vzv | IFA | 1 | 18 | 8786 |
| **Additional antigens** | | | | | |
| *N. meningitidis* | Anti-men A | ELISA | 3 | 5 | 2056 |
| Polio (Inactive) | Anti-polio 2 | PRNT | 3 | 15 | 3270 |
| Polio (Inactive) | Anti-polio 3 | PRNT | 3 | 15 | 3179 |
| Pertussis (acellular) | Anti-pertussis PT | ELISA | 3 | 28 | 9302 |
| Pertussis (acellular) | Anti-pertussis FHA | ELISA | 3 | 28 | 9305 |
| Pertussis (acellular) | Anti-pertussis PRN | ELISA | 3 | 28 | 9335 |
| *S. pneumoniae* | Anti-strep 01 | ELISA | 3 | 16 | 6667 |
| *S. pneumoniae* | Anti-strep 03 | ELISA | 3 | 6 | 2800 |
| *S. pneumoniae* | Anti-strep 05 | ELISA | 3 | 16 | 6670 |
| *S. pneumoniae* | Anti-strep 06a | ELISA | 3 | 12 | 3826 |
| *S. pneumoniae* | Anti-strep 06b | ELISA | 3 | 19 | 7316 |
| *S. pneumoniae* | Anti-strep 07f | ELISA | 3 | 16 | 6702 |
| *S. pneumoniae* | Anti-strep 09v | ELISA | 3 | 19 | 7344 |
| *S. pneumoniae* | Anti-strep 14 | ELISA | 3 | 19 | 7335 |
| *S. pneumoniae* | Anti-strep 18c | ELISA | 3 | 19 | 7345 |
| *S. pneumoniae* | Anti-strep 19a | ELISA | 3 | 12 | 3910 |
| *S. pneumoniae* | Anti-strep 19f | ELISA | 3 | 19 | 7333 |
| *S. pneumoniae* | Anti-strep 23f | ELISA | 3 | 19 | 7349 |
| **Additional immunogenicity measurements** | | | | | |
| Diphtheria | Anti-diphtheria | PRNT | 3 | 4 | 197 |
| Mumps | Anti-mumps | PRNT | 1 | 7 | 5051 |
| *N. meningitidis* | Anti-men A | BAC | 3 | 5 | 1784 |
| *N. meningitidis* | Anti-men C | BAC | 3 | 15 | 4889 |
| *S. pneumoniae* | Anti-strep 01 | OPA | 3 | 15 | 4016 |
| *S. pneumoniae* | Anti-strep 03 | OPA | 3 | 5 | 2266 |
| *S. pneumoniae* | Anti-strep 05 | OPA | 3 | 15 | 3967 |
| *S. pneumoniae* | Anti-strep 04 | OPA | 3 | 15 | 3999 |
| *S. pneumoniae* | Anti-strep 06a | OPA | 3 | 11 | 1978 |
| *S. pneumoniae* | Anti-strep 06b | OPA | 3 | 15 | 3183 |
| *S. pneumoniae* | Anti-strep 07f | OPA | 3 | 15 | 3853 |
| *S. pneumoniae* | Anti-strep 09v | OPA | 3 | 15 | 3921 |
| *S. pneumoniae* | Anti-strep 14 | OPA | 3 | 15 | 3996 |
| *S. pneumoniae* | Anti-strep 18c | OPA | 3 | 15 | 3919 |
| *S. pneumoniae* | Anti-strep 19a | OPA | 3 | 10 | 1791 |
| *S. pneumoniae* | Anti-strep 19f | OPA | 3 | 15 | 3878 |
| *S. pneumoniae* | Anti-strep 23f | OPA | 3 | 15 | 3919 |
| **Multivariate analyses** | | | | | |
| MMRV | Anti-measles | ELISA | 1 | 14 | 6944 |
| | Anti-mumps | ELISA | | | |
| | Anti-rubella | ELISA | | | |
| | Anti-vzv | IFA | | | |
| MMR | Anti-measles | ELISA | 1 | 21 | 11592 |
| | Anti-mumps | ELISA | | | |
| | Anti-rubella | ELISA | | | |
| Hib-Men | Anti-hib | ELISA | 3 | 14 | 2087 |
| | Anti-men C | ELISA | | | |

| | | | | | |
|---|---|---|---|---|---|
| DTP (acellular) | Anti-diphtheria | ELISA | 3 | 20 | 1684 |
| | Anti-tetanus | ELISA | | | |
| | Anti-pertussis PT | ELISA | | | |
| DTP (whole-cell) | Anti-diphtheria | ELISA | 3 | 6 | 221 |
| | Anti-tetanus | ELISA | | | |
| | Anti-pertussis PT | ELISA | | | |
| *N. meningitidis* | Anti-men A | ELISA | 3 | 5 | 2051 |
| | Anti-men C | ELISA | | | |
| *S. pneumoniae 7-serotypes* | Anti-strep 04 | ELISA | 3 | 9 | 951 |
| | Anti-strep 06b | ELISA | | | |
| | Anti-strep 09v | ELISA | | | |
| | Anti-strep 14 | ELISA | | | |
| | Anti-strep 18c | ELISA | | | |
| | Anti-strep 19f | ELISA | | | |
| | Anti-strep 23f | ELISA | | | |
| *S. pneumoniae 10-serotypes* | Anti-strep 01 | ELISA | 3 | 10 | 2773 |
| | Anti-strep 04 | ELISA | | | |
| | Anti-strep 05 | ELISA | | | |
| | Anti-strep 06b | ELISA | | | |
| | Anti-strep 07f | ELISA | | | |
| | Anti-strep 09v | ELISA | | | |
| | Anti-strep 14 | ELISA | | | |
| | Anti-strep 18c | ELISA | | | |
| | Anti-strep 19f | ELISA | | | |
| | Anti-strep 23f | ELISA | | | |
| Pertussis (acellular) | Anti-pertussis PT | ELISA | 3 | 28 | 9273 |
| | Anti-pertussis FHA | ELISA | | | |
| | Anti-pertussis PRN | ELISA | | | |
| Polio (Inactive) | Anti-polio 1 | PRNT | 3 | 15 | 3061 |
| | Anti-polio 2 | PRNT | | | |
| | Anti-polio 3 | PRNT | | | |

HAV = Hepatitis A virus, HBV = Hepatitis B virus, VZV = Varicella Zoster virus, FHA = filamentous hemagglutinin, PRN = pertactin, PT = pertussis toxoid, ELISA = Enzyme-linked immunosorbent assay, OPA = Opsonophagocytosis assay, PRNT = Plaque reduction neutralization test, IFA = Immunofluorescence assay, BAC = Serum bactericidal assay.
